# Characterization of shared and ancestry-specific signals driving complex traits using multi-ancestry fine-mapping

**DOI:** 10.1101/2025.11.13.25339614

**Authors:** Tara Mirmira, Nichole Ma, Jonathan Margoliash, Wilfredo G. Gonzalez Rivera, Tiffany Amariuta, Kelly A. Frazer, Alon Goren, Melissa Gymrek

## Abstract

While most signals identified by genome-wide association studies (GWAS) are shared across populations, the growing size and diversity of GWAS datasets provides evidence that a subset of signals are ancestry-specific. Yet, characterizing these signals remains challenging, since the underlying causal variants are often unknown. Statistical fine-mapping aims to identify candidate causal variants, but struggles to distinguish between variants in high linkage disequilibrium (LD). Multi-study fine-mapping methods can improve resolution by leveraging population-specific LD patterns, but typically assume causal variants are shared and/or polymorphic across studies, making it challenging to study ancestry-specific contributions. To overcome these limitations, we introduce PIPSORT, a multi-study fine-mapping method which simultaneously detects both shared and ancestry-specific signals and quantifies evidence of signal sharing across studies. We applied PIPSORT to fine-map platelet count and LDL cholesterol (LDL-C) in individuals of primarily African vs. European ancestry in the UK Biobank (UKB) and *All of Us* (AoU) datasets. Due to the bias of these datasets toward Europeans (94% in UKB, 49% in AoU), most trait-associated regions identified have strong signals in Europeans. We estimate 89%-99% of these regions are shared with Africans, but detect dozens of examples of ancestry-specific signals. Of these, 10, including known African-specific missense variants in *MPL* (platelet count) and *PCSK9* (LDL-C), could only be confidently detected in the more diverse AoU cohort. We additionally applied PIPSORT to fine-map schizophrenia signals in East Asians vs. Europeans, which identified multiple ancestry-specific signals including a known missense variant in *SLC39A8*. Finally, we leveraged the high degree of admixture within AoU to identify specific signals for platelet count and LDL-C that are driven by interaction with local vs. global ancestry. Overall, our finding that multiple strong ancestry-specific signals could be identified for all traits studied provides novel insights into the genetic architecture of complex traits in different populations and has important implications for development of future multi-ancestry methods for complex trait analysis.

## Introduction

The growing size of genome-wide association study (GWAS) datasets and availability of more diverse cohorts has the potential to improve identification of causal variants across human populations. Although prior work indicates that most causal signals are likely to be shared^1^, increasing evidence suggests ancestry-specific signals may make important contributions to complex traits: multiple variants that are common only in some ancestries are well known to influence complex traits^2–5^. Additionally, the low portability of PRSs based on European datasets^6^ suggests they may be missing important sources of genetic variation specific to other ancestries. Further, although causal variants tend to have similar effects regardless of the ancestral haplotype on which they fall^7^, recent methods that model the impact of local ancestry on effect sizes (e.g. TRACTOR^8^) imply that the effects of some variants may differ across ancestries even when the variants are present in all ancestries.

A major challenge in identifying and characterizing ancestry-specific contributions is that for the majority of GWAS signals the underlying causal variants are unknown. Identifying the causal variants driving these signals is critical for the key goal of translating GWAS to biological insight and may help improve performance, interpretability, and portability of predictive models such as polygenic risk scores (PRS) across ancestries^9^. Yet, pinpointing causal variants has proven to be a major challenge, since most trait-associated variants are not actually causal but simply in linkage disequilibrium (LD) with one or more causal variants (as reviewed in Schaid *et al*.^10^).

Statistical fine-mapping methods aim to pinpoint causal variants, often using a Bayesian approach (such as that introduced by CAVIAR^11^) to compute a per-variant posterior inclusion probability (PIP) based on GWAS summary statistics while modeling correlation between variants due to LD. Some fine-mapping methods additionally take advantage of information beyond GWAS summary statistics and incorporate functional information^12,13^ to inform prior probabilities of causality for each variant. Although these methods identify individual high confidence variants in some cases, it is not always possible to distinguish between variants that are in perfect or nearly perfect LD.

Multi-study fine-mapping methods can overcome this challenge to improve the resolution of causal variant identification by leveraging differences in LD patterns across studies performed in cohorts with different ancestries. Existing methods typically make the assumption that causal variants are shared and polymorphic across studies and in most cases output a single PIP quantifying the overall evidence of causality for each variant^14–16^. Yet, this assumption prevents consideration of variants that only have causal effects in a subset of ancestries. One way such ancestry-specific variants could arise is if a variant is polymorphic in certain populations but monomorphic in others, and thus could not be considered as a causal variant in all groups. We refer to these as *ancestry-specific variants*, or AS-Vs, below. Well-known examples of AS-Vs include protein-coding mutations in *PCSK9* specific to individuals of African ancestry influencing LDL cholesterol^2,3^ and in *APOL1* influencing kidney disease risk (as reviewed in Daneshpajouhnejad *et al*.^4^). Ancestry-specific causal variants could also represent variants whose effects depend either on the local ancestry background on which they fall (as modeled in TRACTOR^8^) or on the global ancestry of an individual (due to either genetic or non-genetic factors associated with ancestry). We refer to these as *ancestry-specific effects*, or AS-Es, below.

Although recently developed multi-study fine-mapping approaches allow for heterogeneity of effect sizes of causal variants, each of these methods only handles a subset of scenarios by which ancestry-specific signals can arise (see **Fig. 1** and **Supplementary Table 1**). In particular, AS-Vs are not accounted for in MsCAVIAR^16^ or MESuSiE^17^, which require the set of variants considered to be present and polymorphic in all studies. MultiSuSiE^15^ and SuSiEx^14^ can handle variants that are polymorphic only in one study but still model variants as shared and do not specifically consider AS-Vs. None except MESuSiE^17^ explicitly consider AS-E cases where the effect is exactly 0 in some studies and non-zero in others. Finally, the results of all of these methods can be highly sensitive to sample size imbalances across ancestries, magnitudes of effect sizes, and prior probabilities that signals are shared, but the effects of these factors on results in simulation or real datasets have not been thoroughly evaluated.

**Figure 1:**
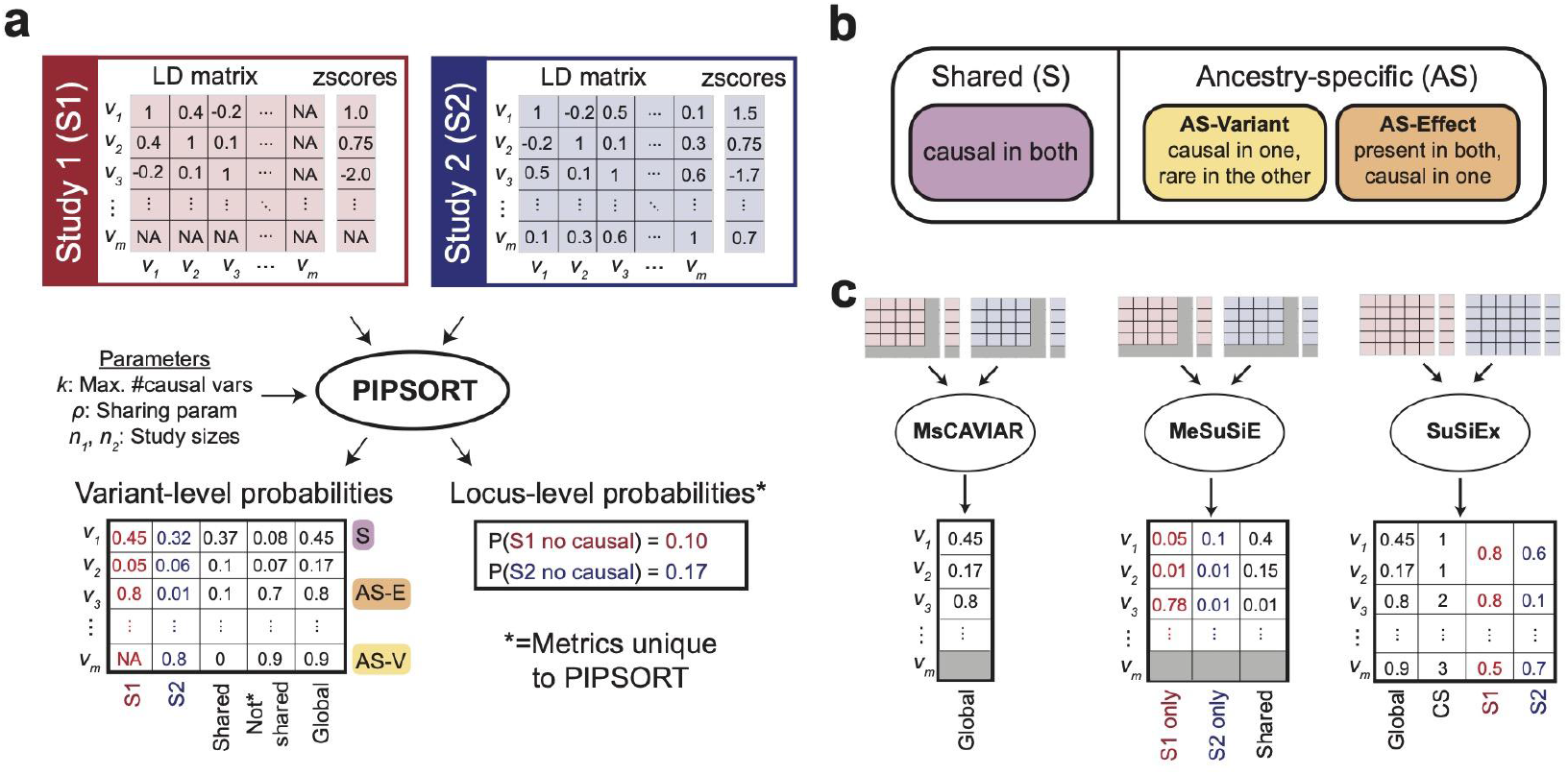
Overview of the PIPSORT method, types of causal signals, and related multi-ancestry fine-mapping methods. (a) Schematic diagram of **PIPSORT**. Provided association summary statistics (Z-scores) and LD information for two studies at a single trait-region, PIPSORT computes multiple causal probabilities for each variant, including separate causal probabilities for each variant in each study and an overall causal probability across all studies. Each reported probability encapsulates different causal information that can then be used to determine if causal variants are shared or study (ancestry)-specific. **(b) Different types of causal signals**. For two ancestries, shared (S) signals are causal across both. Ancestry-specific (AS) signals can be divided into *AS-Variant signals*, which are causal in one ancestry and monomorphic or very low MAF and therefore not detected as causal in the other, and *AS-Effect signals*, which are polymorphic and common in both ancestries but causal in only one. The different probabilities output by PIPSORT can be used to categorize signals as S, AS-Variant or AS-Effect. **(c) Overview of related multi-ancestry fine-mapping methods**. Extending the example from **(a)**, we illustrate the different inputs and outputs for three related fine-mapping methods. Similar to **(a)**, red and blue denote summary statistics for studies 1 and 2, respectively. Grey indicates values for variants that are not considered due to missing data in one of the studies. MsCAVIAR assumes all signals are shared. AS-Vs are not considered by MsCAVIAR or MeSuSiE. SuSiEx models variants as shared but performs a post-hoc computation of ancestry-specific probabilities for each credible set (CS). In that case the credible set to which the AS-Vs belongs receives a PIP of approximately 0.5 in the population in which it is monomorphic. Additional details in **Supplementary Table 1**.

Here, we leverage multiple diverse biobanks and multi-ancestry fine-mapping methods to identify and characterize ancestry-specific genetic contributions to complex traits. To overcome limitations of existing methods, we introduce PIPSORT, a multi-study fine-mapping method which can simultaneously identify shared and study-specific causal variants, including both AS-Vs and AS-Es. Similar to most existing methods, PIPSORT requires only GWAS summary statistics and LD matrices for each study as input. PIPSORT outputs study-specific PIPs for each variant plus additional probabilities of interest, including the probability that each variant is a shared causal signal as well as the probability that a study contains zero causal variants in a region. Our simulation studies demonstrate that PIPSORT outperforms MsCAVIAR, MeSuSiE, and SuSiEx in identifying and distinguishing ancestry-specific vs. shared signals while achieving similar performance on shared signals. We additionally perform detailed benchmarking of all methods, which highlights challenges arising from low sample sizes in existing non-European datasets. Finally, we apply PIPSORT to identify and characterize AS-V and AS-E signals for LDL cholesterol (LDL-C) and platelet count separately in cohorts of primarily African (AFR) vs. European (EUR) ancestry from the UK Biobank (UKB) and *All of Us* (AoU) datasets as well as for schizophrenia (SCZ) in European vs. East Asian (EAS) ancestry cohorts from the Psychiatric Genetics Consortium (PGC). Overall, our results demonstrate that while the majority of signals are likely shared, we identify multiple intriguing examples of ancestry-specific signals, a finding with important implications for complex trait analysis in diverse populations.

## Results

### Overview of the PIPSORT method

PIPSORT, which is an extension of the MsCAVIAR^16^ model, takes as input LD matrices and GWAS summary statistics for two studies for a trait-associated genomic region of interest (termed a *trait-region* or locus; typically ∼1Mb) and computes PIPs for each variant (**Fig. 1a**). Beyond overall PIPs output by MsCAVIAR, PIPSORT computes multiple additional posterior probabilities, including study-specific PIPs at both the variant and locus level, which allows the causal probability of a variant to differ across studies and enables the exploration of ancestry-specific variants as causal signals. We define a *shared* causal signal as a signal that is present and causal in all studies, possibly with different effect sizes. A *study-specific* (or *ancestry-specific*) signal is one that is either (1) present and causal in some ancestries but is monomorphic (and therefore not causal) in the others (AS-V); or (2) present in all ancestries but not causal in all (AS-E) (**Fig. 1b**). Note the terms *ancestry-specific* and *study-specific* are used interchangeably throughout this manuscript as it is common for studies to correspond to different ancestries.

To compute PIPs and related quantities, PIPSORT evaluates a set of configurations, represented by binary vectors that denote which variants are causal in each study. For each configuration (we provide an example in **Supplementary Fig. 1**), PIPSORT computes a posterior probability based on observed LD, summary statistics, and the prior probability of the configuration. A key component of the prior is the *sharing parameter* (ρ), which indicates the prior probability that a causal variant is shared across studies. Because we do not know the true percentage of shared causal variants, we set the default of this parameter to 0.75, based on previous observations across a range of traits that most, but not all, causal effects are shared across ancestries^18–20^. As expected when using a Bayesian approach, results are sensitive to this prior when limited data is available, but become less sensitive as GWAS sample sizes increase. The effect of this parameter, and of analogous parameters for related tools, is evaluated in more detail below. PIPs and related probabilities for each variant are computed by summing posterior probabilities across all configurations in which that variant is set to be causal. A full description of the PIPSORT model is given in **Methods**.

### Benchmarking PIPSORT on simulated data

We first benchmarked PIPSORT using simulated phenotypes based on real genotype data for samples of European and African ancestry from the UK Biobank^21^ (UKB). For each simulation setting, we evaluated cases in which the study sizes were unbalanced (European n=343,760; African n=7,146; “unbal”) as well as balanced (Europeans down-sampled to 8000, roughly the African sample size; “bal”). Full simulation parameters for each setting are provided in **Methods**. We compared PIPSORT’s performance to MESuSiE^17^, which directly outputs ancestry-specific PIPs, as well as SuSiEx^14^, which outputs post-hoc ancestry-specific probabilities per credible set. As a baseline we additionally compared to MsCAVIAR^16^, whose model is most similar to that of PIPSORT but does not allow for computing ancestry-specific PIPs and so could not be directly compared to for some benchmarks below (**Supplementary Figs. 2-3**).

In the first setting, we simulated a single causal variant with varying effect size and minor allele frequency (MAF) across the two ancestries, and set the variant to be causal in both (shared). As expected, the three methods showed similar global PIPs, and showed lower confidence (manifested as lower or more variable PIPs) as the effect size decreased (**Supplementary Fig. 2**). In the case of shared causal variants, ancestry-specific PIPs mirror the global PIPs for each method (**Supplementary Fig. 2**). Notably, in some settings (429/1500 for balanced, 185/1500 for unbalanced) with low effect and/or sample sizes, SuSiEx failed to converge.

We next simulated variants that are polymorphic in both studies but only causal in one (AS-E). For this setting we first evaluated the sensitivity of each tool’s results to the prior probability that a signal is shared, which we set using the sharing parameter for PIPSORT and the ancestry weights for MESuSiE. We observed that the PIPSORT PIPs for the study in which a variant is not causal are closely tied to the value of the sharing parameter at low sample sizes, but this effect can be overcome by increasing the sample size (**Fig. 2a; Supplementary Fig. 3, 4**). It follows that in the unbalanced population setting in UKB it is far easier to confidently detect causal variants that are African-specific, whereas additional data would be required to confidently conclude a causal variant is European-specific. A similar trend was observed when modulating MESuSiE ancestry weights to favor shared vs. ancestry-specific signals (**Fig. 2b, Supplementary Fig. 3**), but in the case of MESuSiE increasing sample size did not overcome the impact of this parameter. Notably, the SuSiEx prior assumes all configurations are equally likely and thus does not enable users to adjust the prior that signals are shared (**Fig. 2c**).

**Figure 2:**
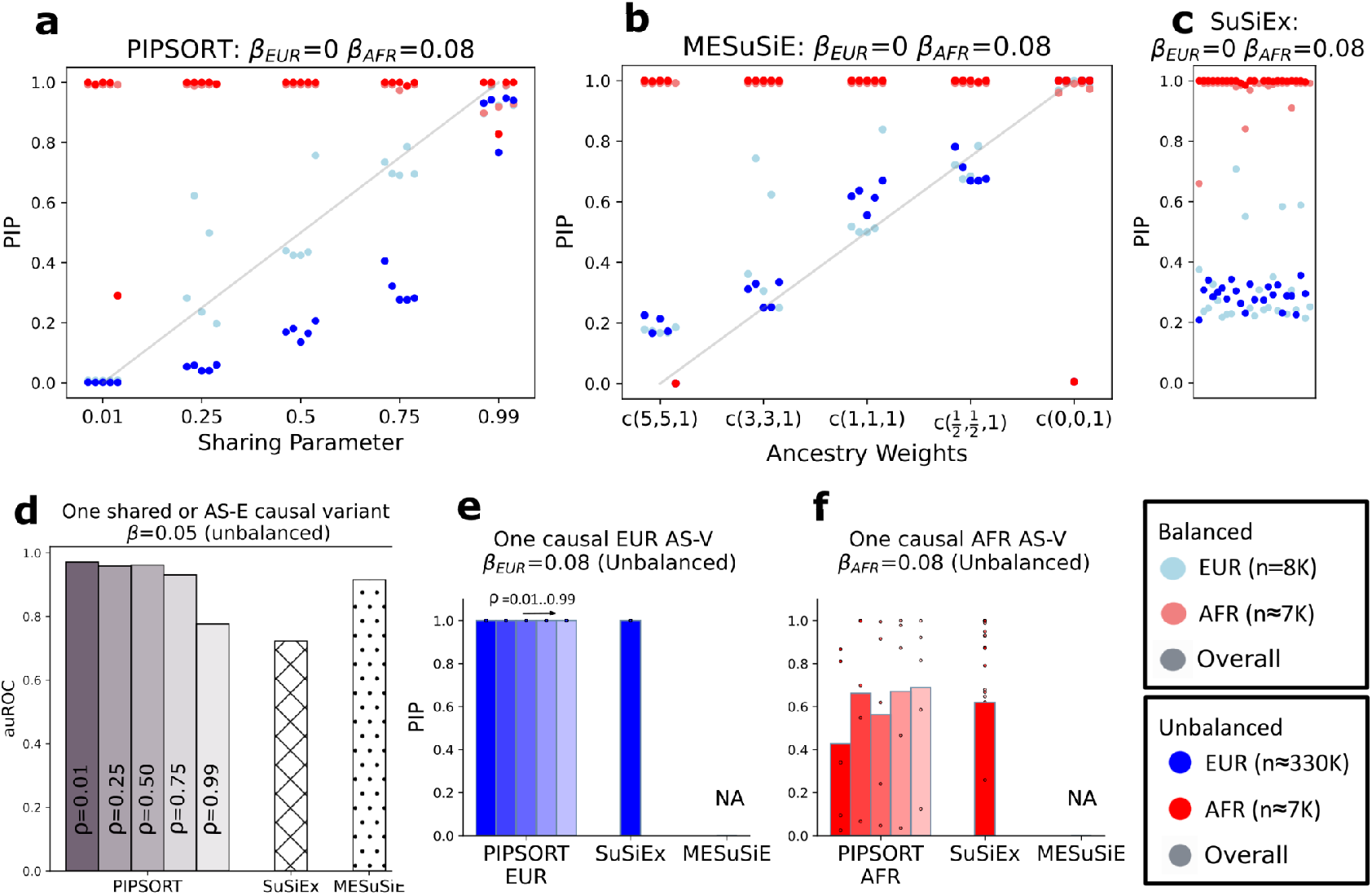
Benchmarking PIPSORT, MeSuSiE, and SuSiEx on simulated data. **(a-c) Evaluating the impact of the prior probability of signal sharing.** We simulated a single African-specific causal variant with β_*AFR*_ = 0. 08. For each prior setting (varying the sharing parameter in PIPSORT and the ancestry weights in MESuSiE), we ran 5 trials and plot the ancestry-specific PIPs. Panels show results for PIPSORT **(a)**, MESuSiE **(b)**, and SuSiEx **(c)**. Notably, SuSiEx does not have an equivalent user-set parameter analogous to the sharing parameter in PIPSORT and thus only one setting with 25 trials is shown. For **a-b**, the x=y line is shown in gray. **(d) auROC for predicting an AS-E variant**. Each bar depicts the area under the receiver operator curve (auROC) for using the difference in ancestry-specific PIPs to predict if a causal variant is an AS-E vs. shared signal. Each bar is computed from 300 simulations: 100 that simulate a EUR AS-E, 100 that simulate an AFR AS-E, and 100 that simulate a shared AS-E, all with an effect size of 0.05. For PIPSORT, bar shading from left to right indicates simulations with different values of the sharing parameter (0.01, 0.25, 0.5, 0.75, 0.99). For MESuSiE, we used ancestry weights w=(3,3,1), which indicates a 3:3:1 ratio of signals that are present in EUR-only, AFR-only, or both and performs similarly to setting the PIPSORT sharing parameter to 0.25. The corresponding results for the balanced setting are provided in **Supplementary Fig. 6. (e-f) Simulation results with a single causal variant polymorphic in one study (AS-V)**. Bars shown are for the unbalanced setting. Bar shading from left to right indicates the sharing parameter (0.01, 0.25, 0.5, 0.75, 0.99). PIPs denoted by each bar are averaged across 5 simulations. The underlying values for each simulation are shown as dots. Note for **e-f**, PIPs are undefined (not 0) for MeSuSiE. SuSiEx bars are averaged across 21 simulations for the EUR AS-V (4 of the 25 trials returned NA for ancestry-specific PIPs) and across 25 simulations for the AFR AS-V. For **a-c** and **e-f**, light blue=European cohort downsampled to 8000 samples; dark blue=full European cohort; light and dark red=full African cohort.

We then evaluated the overall ability of each method to correctly identify ancestry-specific signals. We first looked at the ability to distinguish shared vs. ancestry-specific effect (AS-E) variants based on the difference of their ancestry-specific PIPs. We found that PIPSORT achieved the highest auROC except in the extreme case of setting the sharing parameter to 0.99 (**Fig. 2d**) which essentially forces all signals to be shared. We then considered ancestry-specific variants (AS-Vs). As expected, if the causal variant is only polymorphic in one study, PIPSORT outputs a high PIP and SuSiEx outputs a high post-hoc probability for the credible set for the study in which the variant is causal, whereas MESuSiE and MsCAVIAR (not shown) both exclude the variant from analysis (**Fig. 2b-c**).

Finally, we evaluated PIPSORT’s performance in the setting of two causal variants allowing each variant to have either shared or study-specific effects (**Supplementary Fig. 5**). Similar to the simulations above, PIPSORT tends to report a PIP of 1 for shared causal variants. On the other hand, in cases where the causal variants are not shared, the resulting PIPs depend on the effect sizes, sharing parameter, and relative sample sizes between groups. As in the single variant unbalanced setting, it is easier to detect population specific events in the population with smaller sample size: when both simulated variants are African-specific, they can be detected by PIPSORT (**Supplementary Fig. 5b**). When one of the causal variants is shared and the other is African-specific, the African-specific one is detected only in the balanced simulations (**Supplementary Fig. 5a**). On the other hand, when one of the causal variants is European-specific and the other is African-specific, the European variant is detected but the African variant is detected only in the balanced simulations (**Supplementary Fig. 5c**).

Overall, our simulation results demonstrate that all tested methods show similar performance on shared variants, whereas PIPSORT is best able to distinguish ancestry-specific vs. shared effects and shows the most comprehensive coverage of all signal categories considered (shared, AS-E, and AS-V; **Fig. 1b**). Further, our results highlight general challenges in performing multi-ancestry fine-mapping including the impact of study size imbalance, in which increased statistical power from the larger population might result in missed ancestry-specific signals in the smaller population. For PIPSORT, this limitation can be overcome by increasing the sample size, whereas we did not observe substantial improvements for the other methods when the sample size of the larger cohort was increased (**Fig. 2b-c**). Additional simulations for a range of effect sizes and variant frequencies are shown in **Supplementary Figs. 2-3**.

### Multi-ancestry fine-mapping supports extensive signal sharing across populations

We applied PIPSORT to perform two-study fine mapping in three separate datasets: UK Biobank^21^ (UKB), *All of Us*^22^ (AoU), and schizophrenia cohorts from the Psychiatric Genetics Consortium^23^ (PGC) (**Fig. 3a**). For UKB, we considered studies based on cohorts of self-reported African (AFR, n=7,562) and European (EUR, n=343,760) ancestry. Given the lower overall sample sizes, high phenotype missingness rates, and greater genetic diversity in AoU, we set out to identify two comparable cohorts in that dataset that would have the most power for association testing. For the first cohort, we included individuals with predicted African ancestry (AFR, n=44,384). For the second study, we considered cohorts of either individuals predicted to be of European ancestry (n=119,178) or all individuals of non-African ancestry (n=174,828), which includes many admixed individuals. Collectively these samples have an average of 74% European ancestry (vs. 89% for the European-only cohort) based on global ancestry analysis using Rye^24^ (**Supplementary Fig. 7; Methods**). Overall, we found that the increased sample size of the non-African cohort resulted in improved power for association testing (**Supplementary Fig. 8**), and therefore focused on fine-mapping using the African vs. non-African cohorts in AoU below. For AoU and UKB, we analyzed two complex traits: platelet count (n=334,324 and 6,771, n=98,663 and 19,959 for UKB EUR and AFR, AoU NOT AFR and AFR, respectively, after preprocessing; **Methods**) and LDL cholesterol (LDL-C; n=327,917 and 6,684, 56,057 and 10,699, respectively). For schizophrenia (SCZ), we used publicly available summary statistics for case-control GWAS from PGC based on European (n=127,906) and East Asian (EAS; n=27,363) cohorts.

**Figure 3:**
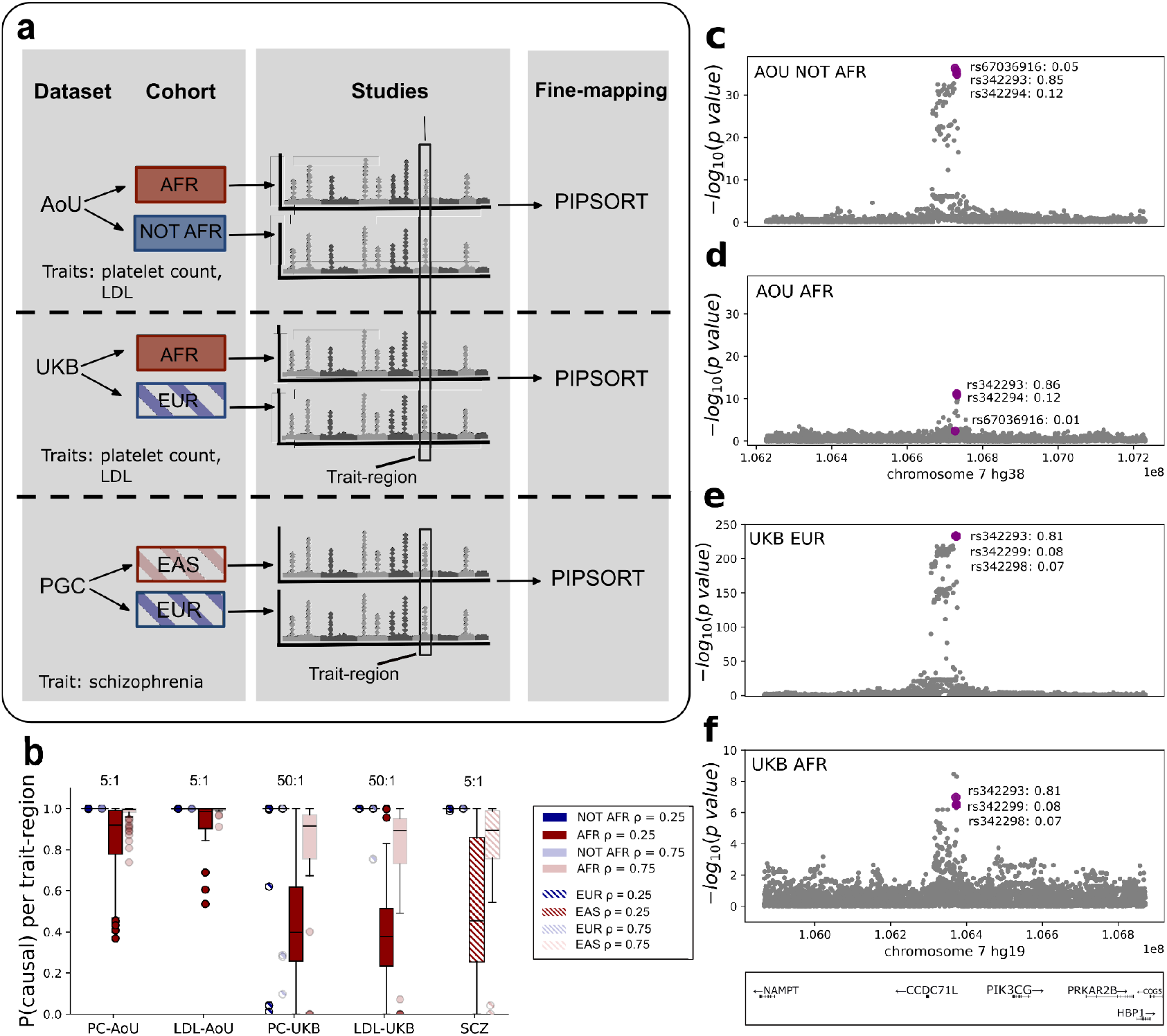
Evidence of extensive signal sharing across African and non-African cohorts in AoU. **(a) Schematic overview of how multi-ancestry fine-mapping was applied to the AoU, UKB and SCZ datasets**. For AoU and UKB, trait-regions were identified jointly across the two datasets and PIPSORT was then applied to each trait-region separately in AoU and UKB. For AoU and UKB, we used in-sample LD matrices. For SCZ, we used the 1000Genomes dataset to compute LD. We performed two-study fine-mapping in each dataset on each trait. **(b) PIPSORT probability of at least one causal variant at each trait-region in each cohort**. Boxplots show the distribution of P(causal), computed as 1-P(no causal variant), for each cohort. The trait is labeled along the x-axis (PC=platelet count, SC=schizophrenia). Results are shown separately for each cohort (blue=NOT AFR, red=AFR, blue hatched=EUR, red hatched=EAS) and for each value of the sharing parameter tested (dark=0.25, light=0.75). The approximate ratio of the larger cohort size to the smaller cohort size for each trait in each dataset is annotated above the boxplots. Results shown are from PIPSORT only as MESuSiE and SuSiEx do not compute an equivalent probability. Note blue boxes are not visible since most values are very close to 1.0. Horizontal lines show median values, boxes span from the 25th percentile (Q1) to the 75th percentile (Q3), and whiskers extend to Q1–1.5*IQR (bottom) and Q3+1.5*IQR (top), where IQR gives the interquartile range (Q3-Q1). Outlier values are shown as individual data points. **c-f. An example trait-region with a high-confidence shared variant. (c)** shows per-variant association statistics (-log10 P-values) for a Chromosome 7 trait-region for platelet count in the AoU NOT AFR cohort, **(d)** for the AoU AFR cohort, **(e)** for the UKB EUR cohort, and **(f)** for the UKB AFR cohort. The fine-mapped variant and corresponding study-specific PIP are labeled in each plot. Purple dots are variants that were fine-mapped in both cohorts across AoU and UKB.

For platelet count and LDL-C, we first performed standard GWAS for each trait in each cohort in AoU and UKB (**Supplementary Figs. 9-10**). As expected based on available sample sizes, the UKB EUR cohort has the highest power to detect associations (manifested by strongest association p-values), followed by the AoU NOT AFR cohort. For both datasets, the non-African cohorts are substantially better powered than the African cohorts. However, this imbalance is less dramatic in AoU. To enable direct comparison of results between AoU and UKB, we used the results from all four GWASs for each trait to construct a unified set of distinct trait-associated regions (termed trait-regions below), each with a minimum length of 1Mb centered at lead variants meeting genome-wide significance (association P<5×10^-8^ in EUR/NOT AFR, P<1×10^-6^ in AFR) in at least one of the four cohorts (**Fig. 3a; Methods**). We obtained a total of 279 and 143 trait-regions for platelet count and LDL-C respectively. We used a conditional regression procedure to set the maximum number of causal variants considered per region, up to a maximum possible of 3 (**Methods**). For SCZ (**Supplementary Fig. 11**), we applied a similar procedure to construct 139 trait-regions and used the number of peak variants added to a trait-region as the maximum number of causal variants for each (**Methods**). For each locus, we ran PIPSORT on each trait-region in each dataset with a sharing parameter (ρ) of either 0.75 or 0.25. Only trait-regions with at least one variant reaching genome-wide significance and at least one variant achieving a study-specific PIP of 0.1 are included in the results below. The full results of each PIPSORT fine-mapping run are reported in **Supplementary Tables 2-5**. For comparison, we also ran MESuSiE and SuSiEx on platelet count and LDL-C in AoU and for SCZ (**Supplementary Data 1; Methods**).

We first examined evidence for sharing of each *trait-region* across cohorts in each dataset, which we assessed using the probability that there exists at least one causal signal in each trait-region, measured separately in each cohort (**Fig. 3b**). Across both traits analyzed in AoU, the median probability of at least one causal variant in each trait-region is >99% in both cohorts with ρ=0.75. As expected, evidence of sharing decreases under the setting of ρ=0.25, but remains high with median >88% (range 89%-99% across both quantitative traits/cohorts). Similar trends were observed for the two traits analyzed in UKB as well as SCZ (**Fig. 3b**) but with lower overall sharing probabilities, which we hypothesize is attributable to the more severe sample size imbalances in those datasets compared to AoU (**Fig. 3b, Supplementary Fig. 12a-b**). We additionally assessed evidence that individual *causal variants* driving signals at each trait-region are shared. Overall, we estimated that the median probability that each independent signal is shared across loci ranges from 0.48-0.52 across both traits in AoU under ρ=0.75 (ranging from 0.28-0.38 for ρ=0.25), suggesting widespread sharing of causal variants. Stratifying both sharing metrics by the best Z-score at each trait-region in the African cohorts shows that evidence of sharing is highest at regions with the strongest association signals (**Supplementary Fig. 13**). Similar trends were observed in UKB and PGC (**Supplementary Figs. 12**,**14**) but with larger gaps between the two cohorts, suggesting trait-regions for which we fail to confidently identify shared signals may be due to insufficient power, particularly in the cohorts with smaller sample size (AFR and EAS), which we discuss in more detail below.

Similarly to the simulation studies, we observed that the sharing parameter has a larger impact on fine-mapping results when the power to detect associations is lower. Overall, per-variant study-specific PIPs are strongly correlated across both cohorts for all traits (**Supplementary Fig. 15**). There is a higher correlation under ρ=0.75 (average r=0.99, P<1e-85) than ρ=0.25 (average r=0.83, P<1e-37), primarily driven by weaker overall PIPs in the non-European cohorts in AoU and SCZ. We observed that PIPs for each variant remain largely consistent across ρ=0.75 vs. 0.25 for the larger cohorts but show substantial variation in the smaller (African/East Asian) cohorts for each dataset (**Supplementary Fig. 16**), with the most dramatic effects at the weakest association signals (**Supplementary Fig. 17**). Our simulations suggest the overall trend toward weaker PIPs in non-European cohorts is primarily driven by power differences due to sample size imbalances, although in some cases, weaker PIPs in one population can indicate ancestry-specific signals, which are discussed further in the next sections.

Finally, we examined shared signals for which fine-mapping results replicated across both AoU and UKB. Overall, we identified 30 and 92 variants with shared PIP≥0.8 in AoU and UKB with ρ=0.75, respectively (19 and 29 with ρ=0.25). Of these, 7 variants with ρ=0.75 (4 remain with ρ=0.25) were supported by both datasets (**Supplementary Table 6**). One example signal for platelet count on Chromosome 7 is shown in **Fig. 3d-e** (AoU) and **Fig. 3f-g** (UKB). In this case, PIPSORT identified variant rs342293 (hg38:7:106731773:C:G) as the likely causal variant with PIP≥0.8 in both cohorts in both datasets (for both ρ=0.25 and ρ=0.75). This variant was also identified by SuSiEx (overall PIP=0.9) and MESuSiE (shared PIP=0.9). This variant overlaps an intergenic candidate *cis* regulatory region identified by ENCODE^25^ at the known platelet count-associated *CCDC71L-PIKC3G* locus^26,27^. Additional examples of shared signals that replicated in both biobanks include a 3’UTR variant in *THPO* associated with platelet count and a 3’UTR variant in *CELSR2* associated with LDL-C (**Supplementary Fig. 18**). Notably, the low overall overlap between shared causal variants identified in both datasets is likely explained by power differences across cohorts and ancestries as well as differences in the sets of variants included in each dataset (see **Discussion**).

### Identification of ancestry-specific causal variants

We next sought to leverage the ancestry-specific PIPs output by PIPSORT to characterize ancestry-specific signals for these traits. For this, we primarily focused on the AoU cohort in which it is easier to detect cohort-specific effects due to the higher African cohort sample size. We first identified candidate trait-regions containing AS-Vs as those for which at least one variant is present in one but not the other cohort and fine-mapped with a PIP≥0.1. Candidate AS-Vs tend to occur at trait-regions with relatively stronger association signals in the target cohort compared to the other (**Fig. 4a-f**). We also observed as expected that the number of detected trait-regions with AS-Vs decreases when increasing the sharing parameter to 0.75 (**Fig. 4a-f**). Similar trends are observed in UKB (**Supplementary Fig. 19**), but with almost no AFR AS-Vs detected (only two at ρ=0.25), likely due to the lower power in the African cohort and more severe population size imbalance in that dataset. Subsequently, to identify a higher confidence set of AS-Vs, we further restricted to trait-regions for which (1) the sum of cohort-specific PIPs for such variants exceeds 0.50 indicating the majority of at least one signal is ancestry-specific, and (2) the minimum Z-score achieved by any such variant is >5 in the African cohort or >6 in the non-African cohort, allowing a less strict threshold in the African cohort to account for power differences. We identified 9 trait-regions with at least one AS-V passing these criteria for platelet count and 3 for LDL-C in AoU. Applying similar criteria identified 4 EUR-specific AS-Vs for SCZ trait-regions using a sharing parameter of 0.25 (**Table 1**), including a known missense variant in *SLC39A8*^28^ (rs13107325) that is common in Europeans but rare (MAF<0.1%) in East Asians. All but 1 candidate AS-V identified by PIPSORT (at the *HP* locus) were also identified by SuSiEx (**Methods**).

**Figure 4:**
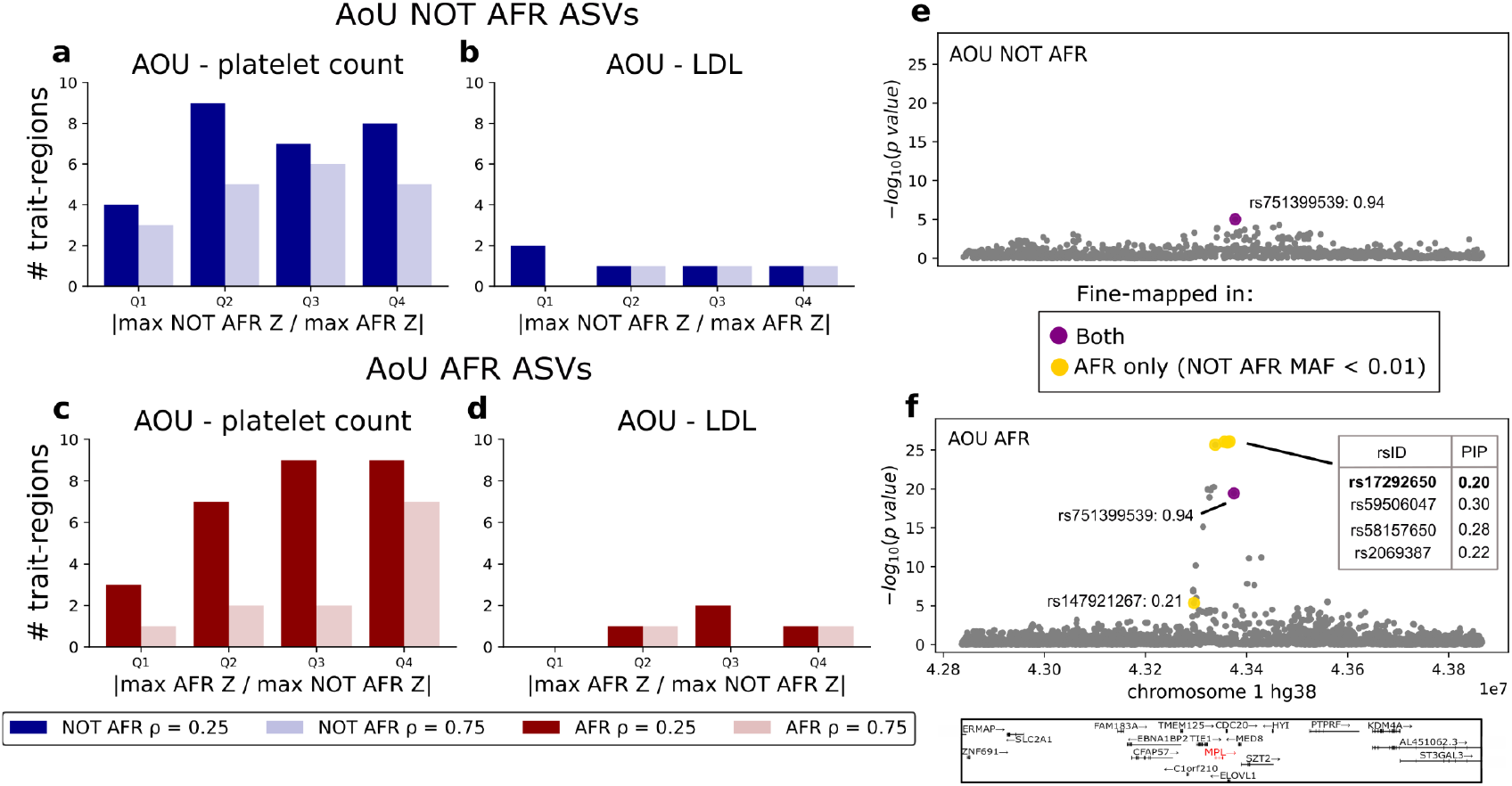
PIPSORT identifies candidate high impact cohort-specific causal variants. **a-b. The number of trait-regions with AS-Vs in the NOT AFR cohort for different values of ρ stratified by the relative significance of NOT AFR to AFR association statistics.** The height of the bar (y-axis) corresponds to the number of trait-regions with NOT AFR AS-Vs. The x-axis stratifies the bars by quartiles for the absolute value of the ratio of the strongest NOT AFR Z-score to the strongest AFR Z-score for a variant in the trait-region. For each quartile, the left bar with darker shading corresponds to ρ=0.25 and the right bar with lighter shading to ρ =0.75. **(a)** shows the results for platelet count and **(b)** for LDL. **c-d. The number of trait-regions with AS-Vs in the AFR cohort for different values of** ρ **stratified by the relative significance of AFR to NOT AFR association statistics**. These plots are the same as those in **(a-b)** but counting the trait-regions with AFR AS-Vs vs. the relative significance of AFR to not AFR association statistics. **(c)** shows the results for platelet count and **(d)** for LDL. Panels **a-d** show results from AoU. Similar plots for UKB and PGC are shown in **Supplementary Fig. 19. e-f. An example trait-region with a high-confidence AFR AS-V corresponding to the *MPL* gene. (e)** shows the Manhattan plot for a Chromosome 1 trait-region for platelet count in the NOT AFR cohort and **(f)** for the AFR cohort. Variants in purple are fine-mapped in both cohorts. variants in yellow are rare in NOT AFR and fine-mapped only in AFR. The PIPs for the cluster of 4 variants at the top are listed in the inset table. The candidate causal missense variant is bolded. The corresponding UKB Manhattan plots are provided in **Supplementary Fig. 21**.

**Table 1:**
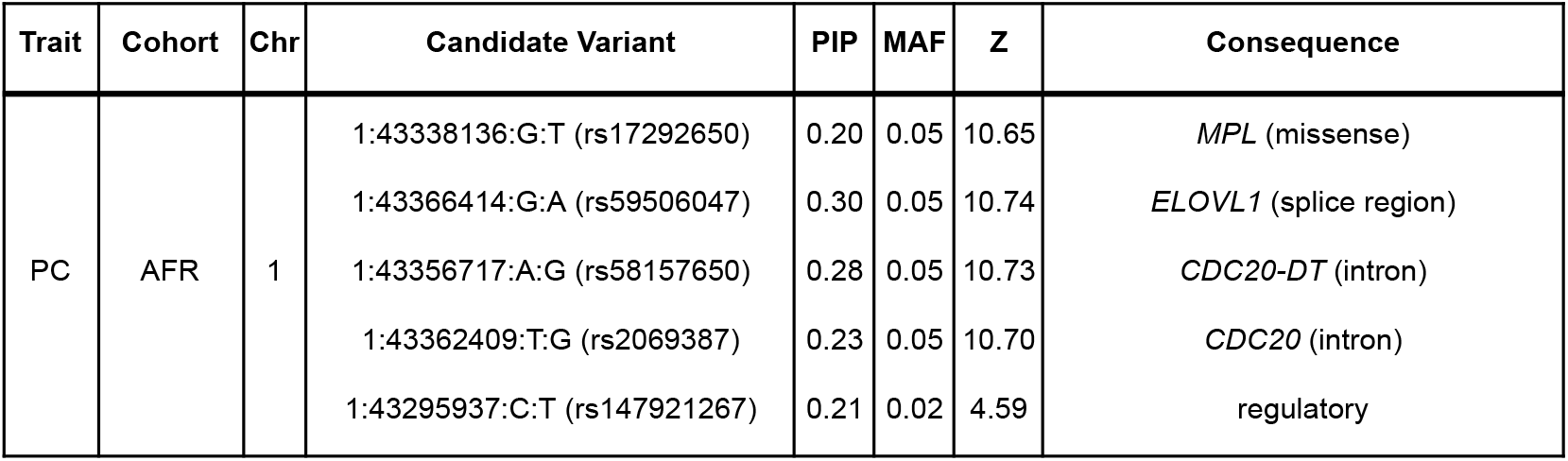
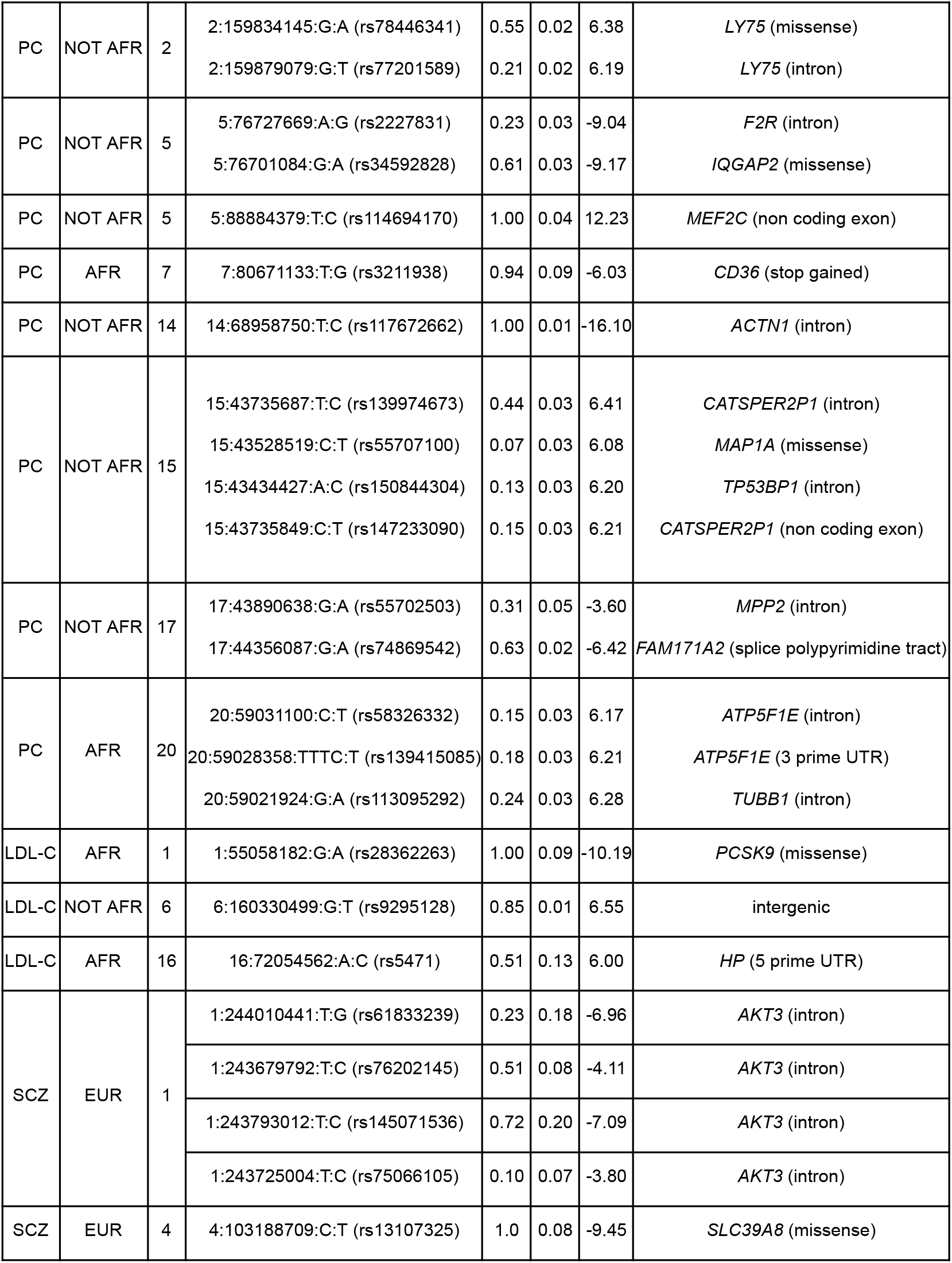
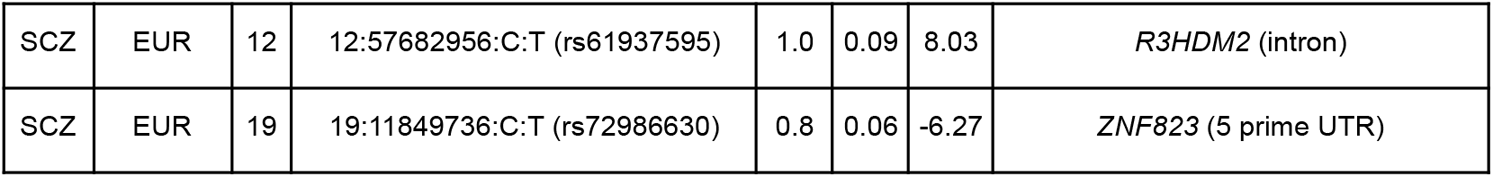
Trait-regions with high confidence AS-Vs identified in AoU and PGC. For each trait-region, we list all candidate AS-Vs alongside their variant effect predictor (VEP) annotation and respective PIP (results shown for ρ=0.25), MAF, and Z-score in the cohort in which they were fine-mapped. We report variants with PIP≥ 0.1 with the exception of 15:43528519:C:T (rs55707100). Variant coordinates are in hg38 for platelet count and LDL-C and hg19 for SCZ. (PC=platelet count). The corresponding table for UKB is provided in **Supplementary Table 7**.

| Trait | Cohort | Chr | Candidate Variant | PIP | MAF | Z | Consequence |
| --- | --- | --- | --- | --- | --- | --- | --- |
| PC | AFR | 1 | 1:43338136:G:T (rs17292650) | 0.20 | 0.05 | 10.65 | *MPL* (missense) |
|  |  |  | 1:43366414:G:A (rs59506047) | 0.30 | 0.05 | 10.74 | *ELOVL1* (splice region) |
|  |  |  | 1:43356717:A:G (rs58157650) | 0.28 | 0.05 | 10.73 | *CDC20-DT* (intron) |
|  |  |  | 1:43362409:T:G (rs2069387) | 0.23 | 0.05 | 10.70 | *CDC20* (intron) |
|  |  |  | 1:43295937:C:T (rs147921267) | 0.21 | 0.02 | 4.59 | regulatory |
| PC | NOT AFR | 2 | 2:159834145:G:A (rs78446341) | 0.55 | 0.02 | 6.38 | *LY75* (missense) |
|  |  |  | 2:159879079:G:T (rs77201589) | 0.21 | 0.02 | 6.19 | *LY75* (intron) |
| PC | NOT AFR | 5 | 5:76727669:A:G (rs2227831) | 0.23 | 0.03 | -9.04 | *F2R* (intron) |
|  |  |  | 5:76701084:G:A (rs34592828) | 0.61 | 0.03 | -9.17 | *IQGAP2* (missense) |
| PC | NOT AFR | 5 | 5:88884379:T:C (rs114694170) | 1.00 | 0.04 | 12.23 | *MEF2C* (non coding exon) |
| PC | AFR | 7 | 7:80671133:T:G (rs3211938) | 0.94 | 0.09 | -6.03 | *CD36* (stop gained) |
| PC | NOT AFR | 14 | 14:68958750:T:C (rs117672662) | 1.00 | 0.01 | -16.10 | *ACTN1* (intron) |
| PC | NOT AFR | 15 | 15:43735687:T:C (rs139974673) | 0.44 | 0.03 | 6.41 | *CATSPER2P1* (intron) |
|  |  |  | 15:43528519:C:T (rs55707100) | 0.07 | 0.03 | 6.08 | *MAP1A* (missense) |
|  |  |  | 15:43434427:A:C (rs150844304) | 0.13 | 0.03 | 6.20 | *TP53BP1* (intron) |
|  |  |  | 15:43735849:C:T (rs147233090) | 0.15 | 0.03 | 6.21 | *CATSPER2P1* (non coding exon) |
| PC | NOT AFR | 17 | 17:43890638:G:A (rs55702503) | 0.31 | 0.05 | -3.60 | *MPP2* (intron) |
|  |  |  | 17:44356087:G:A (rs74869542) | 0.63 | 0.02 | -6.42 | *FAM171A2* (splice polypyrimidine tract) |
| PC | AFR | 20 | 20:59031100:C:T (rs58326332) | 0.15 | 0.03 | 6.17 | *ATP5F1E* (intron) |
|  |  |  | 20:59028358:TTTC:T (rs139415085) | 0.18 | 0.03 | 6.21 | *ATP5F1E* (3 prime UTR) |
|  |  |  | 20:59021924:G:A (rs113095292) | 0.24 | 0.03 | 6.28 | *TUBB1* (intron) |
| LDL-C | AFR | 1 | 1:55058182:G:A (rs28362263) | 1.00 | 0.09 | -10.19 | *PCSK9* (missense) |
| LDL-C | NOT AFR | 6 | 6:160330499:G:T (rs9295128) | 0.85 | 0.01 | 6.55 | intergenic |
| LDL-C | AFR | 16 | 16:72054562:A:C (rs5471) | 0.51 | 0.13 | 6.00 | *HP* (5 prime UTR) |
| SCZ | EUR | 1 | 1:244010441:T:G (rs61833239) | 0.23 | 0.18 | -6.96 | *AKT3* (intron) |
|  |  |  | 1:243679792:T:C (rs76202145) | 0.51 | 0.08 | -4.11 | *AKT3* (intron) |
|  |  |  | 1:243793012:T:C (rs145071536) | 0.72 | 0.20 | -7.09 | *AKT3* (intron) |
|  |  |  | 1:243725004:T:C (rs75066105) | 0.10 | 0.07 | -3.80 | *AKT3* (intron) |
| SCZ | EUR | 4 | 4:103188709:C:T (rs13107325) | 1.0 | 0.08 | -9.45 | *SLC39A8* (missense) |
| SCZ | EUR | 12 | 12:57682956:C:T (rs61937595) | 1.0 | 0.09 | 8.03 | *R3HDM2* (intron) |
| SCZ | EUR | 19 | 19:11849736:C:T (rs72986630) | 0.8 | 0.06 | -6.27 | *ZNF823* (5 prime UTR) |

Trait-regions with candidate AS-Vs passing these criteria in AoU included multiple known African-specific associations. One example is the *PCSK9* locus, which is well-known to harbor variants with strong effects on LDL that are more common in African individuals^2^. PIPSORT identified rs28362263 (hg38:1:55058182:G:A), a missense variant (A443T) known to result in loss of function^3^ of *PCSK9* that is common in Africans (MAF=0.10 in AoU) but not included due to low frequency in non-Africans (MAF<0.0093 in AoU), as an AS-V (AFR PIP=1.0; **Supplementary Fig. 20**). Another known example captured by PIPSORT is a trait-region on Chromosome 1 associated with platelet count at which a cluster of 5 variants specific to the African cohort are assigned PIPs≥0.2 (**Fig. 4e-f**). One variant in this cluster, rs17292650 (hg38:1:43338136:G:T) is a missense variant in *MPL* (K39N), with high frequency in Africans only (MAF=0.05 in AoU AFR). This variant, which is associated with reduced function of the MPL protein, a thrombopoietin receptor, was previously shown to result in a clinical phenotype of thrombocytosis specifically in African Americans^29^. A third example is a region on Chromosome 7 associated with platelet count, which includes rs3211938 (hg38:7:80671133:T:G; AFR PIP=0.94), an African-specific nonsense mutation in *CD36*, which has previously been shown to be under selection and associated with malaria susceptibility^30^ (**Supplementary Fig. 22**). Although these high impact African-specific protein-coding variants were previously known, they serve as positive controls and also highlight known high-confidence variants that are missed by alternative methods that only consider variants polymorphic across all populations studied.

Beyond these known examples, PIPSORT identified an African-specific AS-V in a trait-region on Chromosome 20 associated with platelet count (**Supplementary Fig. 23**). In addition to at least one shared signal, this region harbors three tightly linked (r=0.99) variants, each with AFR MAF=0.03 and PIPs≥0.2 and strong associations in the African cohort (P=3.36e-10, 5.50e-10) that are very rare in non-Africans. One of these, rs113095292 (hg38:20:59021924:G:A), is an intronic variant in *TUBB1*, a gene encoding the beta-1 tubulin protein which has been shown to play a key role in platelet formation^31^. Another, rs139415085 (hg38:20:59028358:TTTC:T), results in the deletion of a single repeat unit at a trinucleotide TTC repeat in the 3’ UTR of *ATP51FE* which is approximately 2Kb downstream of *TUBB1*. None of these variants were identified by MESuSiE (which does not consider AS-Vs). SuSiEx placed all three candidate variants identified by PIPSORT in an African-specific credible set with an overall PIP of 0.30 and non-African and African post-hoc PIPs of 0.53 and 0.86 respectively. Notably, none of the African-specific AS-Vs described above were detected in UKB, either because they were not included in the variant sets used or due to lack of power (**Fig. 4e-f; Supplementary Figs. 20-23**). Overall, these examples further highlight the benefit of increasing sample sizes for more diverse cohorts to detect additional AS-Vs.

PIPSORT additionally identified multiple candidate AS-Vs in the non-African cohort in AoU. One example is a region on Chromosome 5 associated with platelet count. The variant rs34592828 (hg38:5:76701084:G:A), a missense variant in *IQGAP2* that has previously been associated with platelet count and mean platelet volume^32^, is very rare (MAF=0.007) in the African cohort, but has a MAF=0.03 in the non-African cohort in which it was fine-mapped with a PIP of 0.61 (**Supplementary Fig. 24**). Other variants identified as non-African AS-Vs include a missense variant in *LY75* and a variant in a non-coding exon of *MEF2C* associated with platelet count (**Supplementary Figs. 25-26**). While these signals have been identified in previous studies^32,33^, here we identify them as candidate causal variants specific to non-African individuals. Additional high-confidence AS-Vs can be found in **Table 1**.

### Identification of variants with ancestry-specific effects driven by local vs. global ancestry

We next examined fine-mapped trait-regions for evidence of ancestry-specific effects (AS-Es). In contrast to the AS-Vs described above, in which a causal variant is cohort-specific because it is monomorphic or extremely rare in the other cohort, we define AS-Es as variants that are polymorphic in both cohorts but only have a causal effect in one. We identified candidate AS-Es as variants fine-mapped with a PIP difference of at least 0.1 between the two cohorts and for which the probability that the variant was not a shared signal was at least 0.1. Overall, we identified a total of 74 trait regions with ρ=0.25 (21 with ρ=0.75) harboring a candidate AS-E in AoU. Similarly to AS-Vs, we found that the number of candidate AS-Es identified is strongly influenced by the power to detect associations in a trait region: the majority of AS-Es identified are specific to the AoU NOT AFR, UKB EUR, and PGC EUR cohorts. Further, we observed fewer trait-regions with AS-Es in one cohort as the association statistics for the other cohort become more significant (**Supplementary Fig. 27**).

In practice, we found that a shared variant may be incorrectly identified as an AS-E due to low power in one population, a limitation that has been discussed in previous work^15^ and that is amplified by sample size imbalances across ancestries in current datasets. To restrict to a higher confidence set of AS-Es, we applied a second round of stricter filtering (**Methods**) including restricting to trait regions in which one population has a high probability (≥0.3) of not having any causal variants. This locus-level probability is uniquely computed by PIPSORT (**Fig. 1a**) and is important for confidently characterizing AS-Es given the limited power in the non-European cohorts and sample size imbalances. Notably, although these criteria likely remove false positives, they also remove true AS-Es in trait-regions that harbor any signal in the other population. A side effect is that all AFR-specific AS-Es were filtered, since nearly all trait-regions analyzed have at least one signal in non-Africans. After filtering, a total of 6 candidate trait-regions with high confidence AS-Es were identified in AoU (**Table 2;** UKB and SCZ results shown in **Supplementary Tables 8-9**). Example AS-Es supported by both AoU and UKB are shown in **Fig. 5** and **Supplementary Figs. 28-29**.

**Figure 5:**
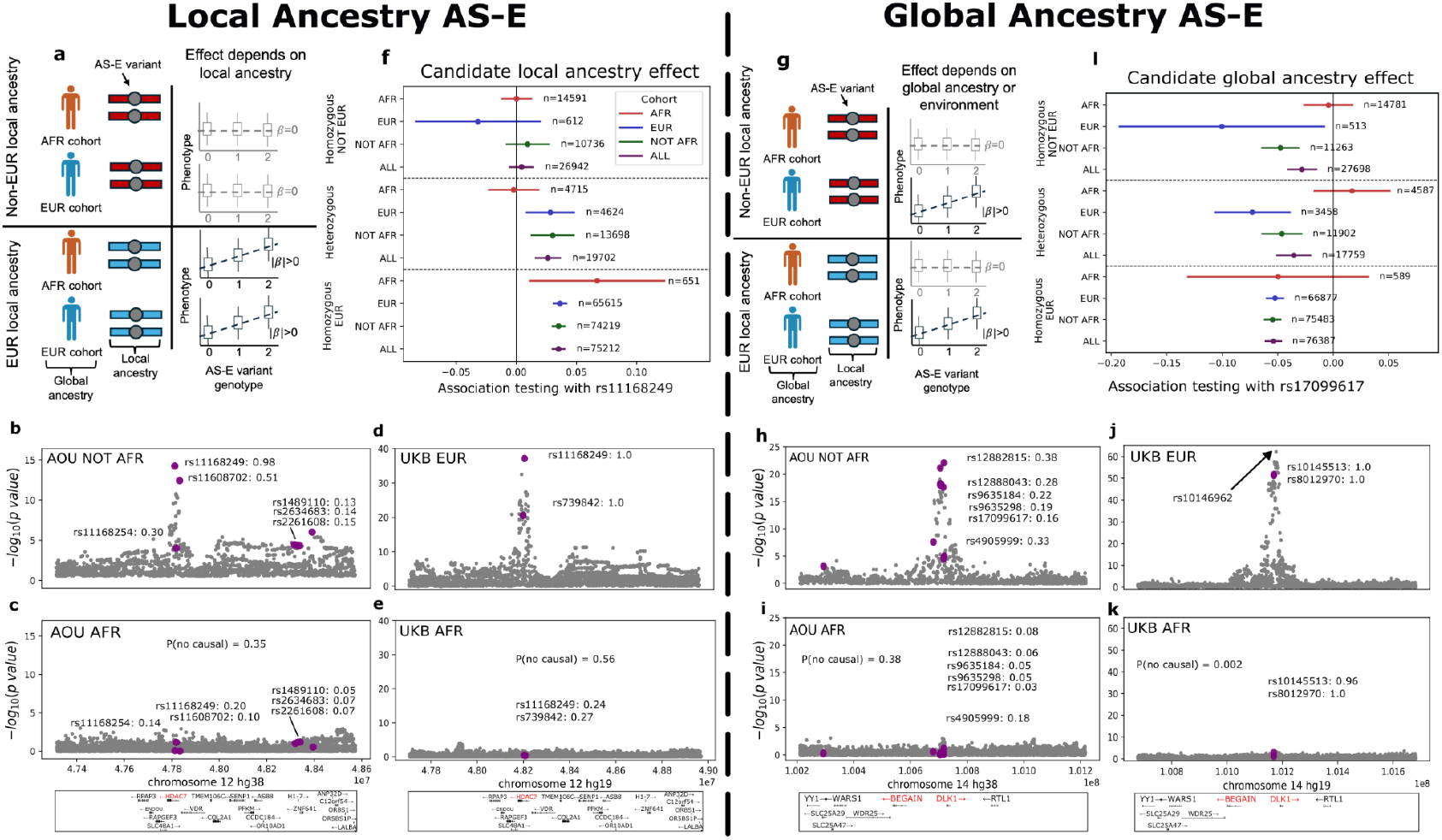
Example candidate ancestry specific effects. **a. Strategy for identifying AS-Es driven by local effects.** For an AS-E whose effect depends on local ancestry, we expect it to have a significant association in samples homozygous for local European ancestry regardless of the global ancestry label. **b-e. Example AS-E dependent on local ancestry**. For a Chromosome 12 trait-region overlapping *HDAC7* associated with platelet count, Manhattan plots are shown for AoU NOT AFR **(b)**, AoU AFR **(c)**, UKB EUR **(d)**, and UKB AFR **(e). f. Association of the Chromosome 12 candidate AS-E stratified by local ancestry in 4 cohorts**. Colors denote the cohort (red=African, blue=European, green=not African, purple=all samples). **h. Strategy for identifying AS-Es driven by global ancestry effects**. For AS-Es whose effect depends on global ancestry or an environmental factor correlated with global ancestry, we expect the AS-E to have a significant association only in samples with global European ancestry regardless of local ancestry. **h-l. Example AS-E dependent on global ancestry**. Similarly, for a Chromosome 14 trait-region associated with platelet count, Manhattan plots are shown for AoU NOT AFR **(h)**, AoU AFR **(i)**, UKB EUR **(j)**, and UKB AFR **(k)**. In **(h)** and **(i)**, fine-mapped variants are approximately matched with their corresponding purple dot, but due to space constraints, are also listed in order of decreasing association significance in the AoU NOT AFR cohort. **(l)** is similar to (**f**) but for the Chromosome 14 trait-region. For **b-e** and **h-k**, variants fine-mapped in both cohorts are shown in purple. For **(f)** and **(l)**, we plot the effect size +/-1 s.e. from association testing of the candidate AS-E. Results from association testing with samples homozygous for non-European ancestry are shown in the top 4 rows of **(f)** and **(l)**, with samples heterozygous for European ancestry in the middle 4 rows, and with samples homozygous for European ancestry in the bottom 4 rows.

**Table 2:**
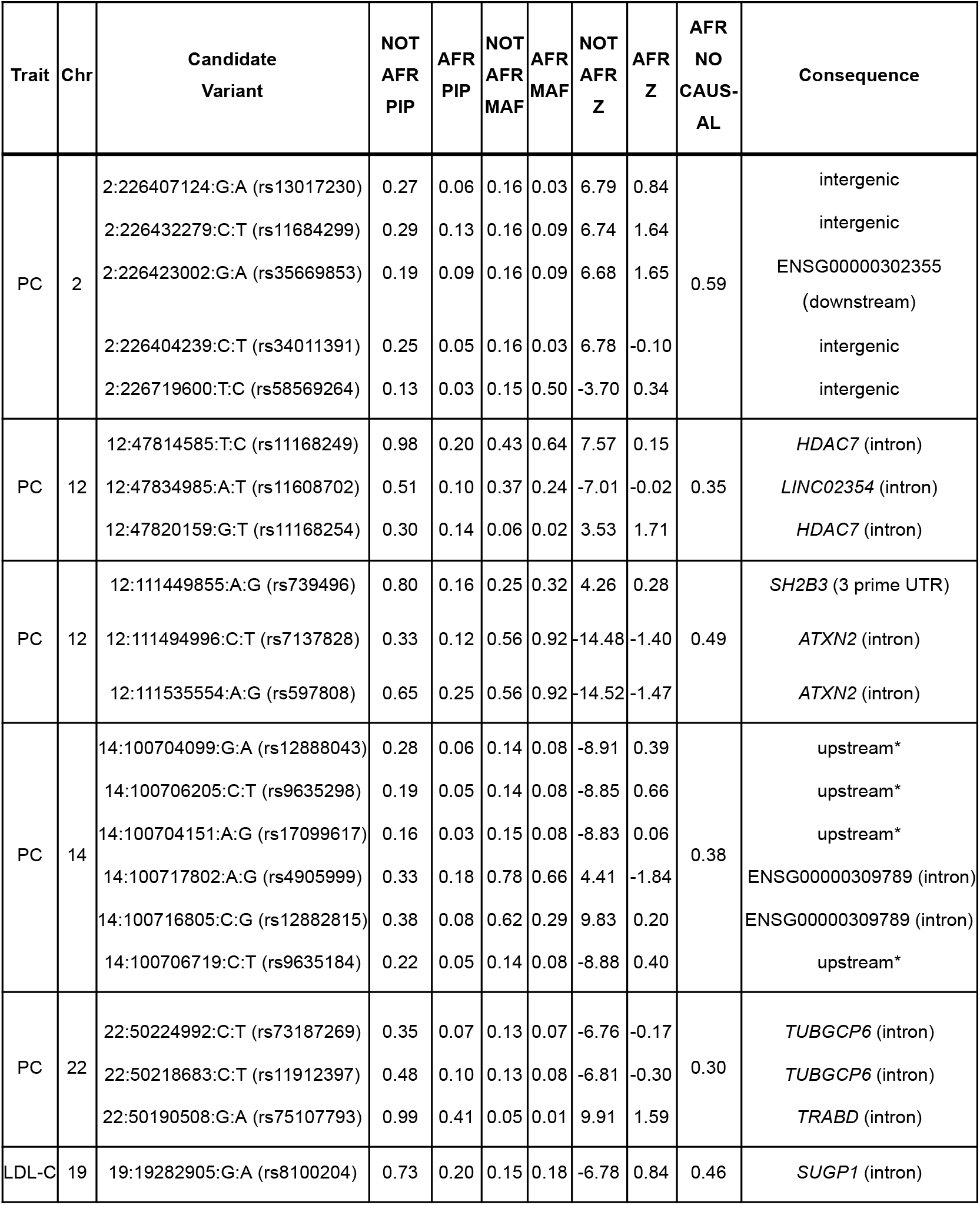
Trait-regions with high confidence AS-Es identified in AoU. For each trait-region, we list all candidate AS-Es alongside their VEP annotation and respective PIPs (with ρ=0.25), MAFs, and Z-scores in both the NOT AFR and AFR cohorts. All trait-regions listed contain candidate NOT AFR AS-Es and we note the corresponding probability, P(AFR no causal), that the AFR cohort does not have a causal signal in this region. Platelet count is abbreviated by PC. The corresponding table for UKB is provided in **Supplementary Table 8** and for SCZ in **Supplementary Table 9**. *Variants whose consequence is labeled with an asterisk for the Chromosome 14 trait-region associated with platelet count are upstream of multiple non-coding RNAs of which this region is well known to have a high density^35^.

We next considered why a variant might be present in both cohorts but only causal in one. Potential scenarios that could give rise to AS-Es include interaction with other variants in *cis* or *trans* or environmental factors that are more prevalent in one cohort (**Supplementary Fig. 30a**). Alternatively, a variant may appear to have an ancestry-specific effect if it tags an underlying but ungenotyped causal variant in only one cohort (**Supplementary Fig. 30b**). We reasoned that effects driven by either interaction with, or tagging of, ancestry specific variants in *cis* would depend primarily on the local ancestry of an individual in that region, rather than that individual’s global ancestry assignment. In this case, AS-E variants identified to be specific to Europeans would be expected to have an effect in admixed individuals even if they are primarily of African ancestry, as long as they inherited a genomic segment of European origin in this particular region (**Fig. 5a**). Notably, many individuals in the AoU AFR cohort are admixed, with an average of 15% European ancestry (**Supplementary Fig. 7**), enabling us to identify individuals from that group with both African and European ancestry in particular regions.

To quantify evidence that AS-Es could be driven by local ancestry, we tested for an interaction between the variant effect and local ancestry in the region harboring each AS-E (**Methods**). Of 21 candidate AS-Es tested across 6 trait-regions, 5 candidate AS-Es across 4 unique trait-regions showed nominal evidence (two-tailed P<0.05) of an interaction with local ancestry and 2 remained significant after multiple hypothesis correction. We obtained similar results using a Welch t-test to compare results from stratified regression with individuals homozygous for European vs. non-European local ancestry at each region (**Supplementary Fig. 31**). We note that some AS-Es that did not pass this test still showed evidence suggestive of local ancestry effects when stratifying individuals based on local ancestry (**Supplementary Fig. 32**) and could just be underpowered to detect an interaction.

One example candidate AS-E with evidence of a local ancestry effect (interaction effect two-sided P=0.03 for rs11168249/hg38:12:47184585:T:C, 0.08 for rs11608702/hg38:12:47834985:A:T, 0.80 for rs11168254/hg38:12:47820159:G:T) was identified at a Chromosome 12 trait-region associated with platelet count. In this region, we detected three variants (rs11168249 and rs11168254, in the first intron of *HDAC7* and rs11608702, in an intergenic region between *HDAC7* and *VDR*) fine-mapped with high PIPs in the AoU NOT AFR cohort, but with low PIPs in AoU AFR (**Fig. 5b-c**). The variant rs11168249, which is common in both Europeans (MAF=0.47) and Africans (MAF=0.65), was also fine-mapped in UKB and was similarly assigned a high PIP in the EUR cohort and low PIP in the AFR cohort (**Fig. 5d-e**). In both datasets, PIPSORT assigned this trait-region a relatively high probability of having no causal variant in the AFR cohorts (56% in UKB and 35% in the AoU dataset). Although rs11168249 did not have a significant association when testing the AFR cohort overall, we found that rs11168249 has a significant association with platelet count when restricting to individuals homozygous for European ancestry at this region (effect size 0.035 +/-0.005; P=9.73e-13; n=75,212) but not when restricting to individuals homozygous for non-European ancestry (effect size 0.004 +/-0.01; P=0.64; n=26,942; **Fig. 5f**). Individuals heterozygous for European ancestry showed an intermediate effect with suggestive significance (effect size 0.026 +/-0.01; P=0.009; n=19,702). Overall, these results suggest that the effect of rs11168249 depends on having European *local ancestry*, and does not depend on the ancestry of an individual elsewhere in the genome.

Inspection of stratified regression results identified a subset of AS-Es that remained strongly significant across non-African samples regardless of the local ancestry label at the AS-E. This suggests that these AS-Es are driven by either global ancestry or other factors (**Fig. 5g**, right column; example in **Fig. 5l**). Of the 6 trait-regions harboring at least one AS-E specific to non-Africans, 4 had at least one AS-E with a nominally significant association even when restricting to individuals homozygous for non-European ancestry and 2 had at least one AS-E that remained significant after multiple hypothesis correction.

One example AS-E that could not be explained by European local ancestry was identified at a Chromosome 14 trait-region associated with platelet count, in which six candidate NON AFR variants were detected in AoU (**Fig. 5h-i**). This region also shows evidence of an ancestry-specific effect in UKB (**Fig. 5j-k**). The region overlaps the *DLK1-DIO3* imprinted locus^34^, which also contains one of the largest clusters of microRNAs in the human genome^35^. A previous study showed that differences in *PAR4*-mediated platelet activation between Black vs. White study participants may be partially driven by increased expression of microRNAs in this region in White individuals^36^. These results are consistent with our findings of an ancestry-specific role for this locus in individuals of European vs. African ancestry.

Stratified association testing with one of the candidate AS-E variants in this region (rs17099617; hg38:14:100704151:A:G) revealed significant associations with individuals homozygous for non-European ancestry (Effect size -0.03 +/-0.01; P=0.03; n=27,698), heterozygous for European ancestry (Effect size -0.035 +/-0.015; P=0.02; n=17,759), and homozygous for European ancestry (Effect size -0.054 +/-0.007; P=1.7e-14; n=76,387; **Fig. 5l**). This shows that while the effect of rs17099617 is specific to non-Africans, it is not dependent on having European local ancestry. Similar results were obtained for stratified regressions using the other five candidate AS-Es in this region (**Supplementary Fig. 33**). Overall, these results suggest the ancestry-specificity of effects in this region are due to global, rather than local European ancestry.

## Discussion

Here we applied multi-ancestry fine-mapping to characterize the contribution of ancestry-specific signals to multiple complex traits. Because existing multi-ancestry fine-mappers only consider a subset of scenarios by which ancestry-specific signals arise, we developed PIPSORT, which enables simultaneous detection of shared causal variants, as well as ancestry-specific variants (AS-Vs) and ancestry-specific effects (AS-Es). Simulation studies demonstrate that PIPSORT is better able to distinguish ancestry-specific effects while maintaining similar performance on shared signals compared to existing tools. We applied PIPSORT to perform two-study fine-mapping for two quantitative traits (platelet count and LDL cholesterol) in two separate datasets (UKB and AoU) and one for a case-control trait (schizophrenia). Finally, we characterized several example signals identified for these traits and evaluated the concordance of fine-mapping results across UKB and AoU.

PIPSORT identified many candidate signals driven by variants that are polymorphic in only a single ancestry group considered (AS-Vs). These include several known signals, including high-impact African-specific missense variants in *PCSK9* and *MPL*, as well as new examples of candidate AS-Vs in both African and non-Africans (**Table 1**) such as missense variants in *LY75* and *IQGAP2* associated with platelet count in non-Africans and variants in or near *TUBB1* associated with platelet count in Africans. Importantly, these examples are missed by multi-ancestry fine-mapping tools that require variants to be polymorphic in all populations considered, and the African-specific examples could not be found by fine-mapping on European datasets alone.

Although our results are broadly consistent with previous studies showing most causal variants have similar effects regardless of ancestry^7^, we also identified multiple intriguing examples of AS-Es, in which a variant is polymorphic in both populations but identified as a candidate causal variant only in one (**Table 2; Fig. 5; Supplementary Figs. 28-29**). Our analysis only identified example AS-Es in primarily European cohorts. However, we suspect that examples also exist in the non-European cohorts but identification of these is challenging due to sample size imbalances, which biases both association testing and fine-mapping towards identification of signals in the larger cohort. Further study of these signals suggests that a subset of AS-Es are driven by local ancestry (**Fig. 5b-f; Supplementary Fig. 31**) indicating a potential interaction with other variants in *cis*. We hypothesize that a substantial subset of AS-Es that appear to be driven by local-ancestry specific effects may actually be driven by un-genotyped complex variants with ancestry-specific allele distributions and LD patterns (**Supplementary Fig. 30b**). One example of this is at the well-known *LPA* locus associated with lipoprotein(a) concentration [Lp(a)]. Previous work found that local ancestry is strongly associated with Lp(a) and that common SNPs in the region show associations with Lp(a) in European but not African populations^37^. However, this is likely driven by the fact that the main causal variant in the region is a highly polymorphic tandem repeat which is differentially tagged by bi-allelic SNPs in Africans vs. Europeans^37^. On the other hand, some AS-Es appear to depend on global ancestry. These could potentially be driven by interaction in *trans* with ancestry-specific variants or by interaction with ancestry-specific non-genetic factors. Additional work is needed to definitively determine why certain variants show ancestry-specific associations, and future studies of these regions have the potential to reveal novel insights into the mechanisms driving population-level differences in these traits.

Regardless of the underlying biological explanation of how AS-Es arise, the presence of such variants has important implications for the development of polygenic risk scores (PRSs) constructed from common SNPs. In the examples identified in **Table 2**, a PRS trained on European cohorts might assign strong non-zero weights to one or more variants in those regions, even though those variants have no predictive power in African individuals and should likely be excluded. Further, there likely exist African-specific AS-Es that, if known, could boost the predictive power of PRSs for these traits in African cohorts. Ultimately, identification and genotyping of the truly causal variants in these regions, rather than LD proxies, is likely to lead to the best predictive performance.

While we identify many intriguing ancestry-specific examples, our results suggest that the majority of GWAS signals for these traits are shared across the ancestries we studied, a finding that is consistent with modeling assumptions often made by trans-ancestry analysis tools^14–16^. Yet, precisely quantifying the number of signals that are shared vs. ancestry-specific remains challenging, in large part due to the limited power to detect associations in non-European cohorts with the current small sample sizes. Both our simulation experiments as well as results from real data demonstrate that study-specific PIPs as well as the estimated probability that a causal variant is shared can be sensitive to the prior probability defined by the sharing parameter, particularly in cases where association tests are underpowered. Further, in cases where the two studies have highly unbalanced sample sizes, PIPSORT tends, as expected, to assign higher PIPs to variants that are causal in the more well-powered study. Our simulation studies demonstrate that power has a similarly large impact even in the single-ancestry fine-mapping setting, with low effect sizes or low sample sizes having a dramatic influence on the stability of the resulting per-variant PIPs (**Supplementary Fig. 2**). Further, our results demonstrate similar sensitivities of existing multi-ancestry fine-mappers to this prior probability of sharing (**Fig. 2a-c**).

These power and sample size imbalance challenges have important implications in real data applications: first, determining whether a variant truly has an ancestry-specific effect in the larger non-African cohorts is challenging, since there may just be insufficient power to detect the association in the African cohorts. Second, we are likely missing ancestry-specific variants and effects in the African cohorts which did not reach required significance thresholds to include in fine-mapping. On the other hand, we have higher confidence in variants that are identified to be causal only in the African cohorts since the large sample sizes in non-Africans make it unlikely that those variants were missed in those cohorts due to low power. Overall, our findings suggest that as sample sizes for non-European individuals continue to increase, this challenge will be overcome and likely uncover additional ancestry-specific causal variants and effects. Indeed, whereas very few African-specific signals could be identified in UKB, we found multiple high confidence examples in AoU, in which the African sample size is around 3 times larger for most traits.

Application of PIPSORT separately in UKB and AoU resulted in surprisingly little overlap between variants confidently identified as causal in each dataset (e.g., only 7 shared variants with strong PIPs in both cohorts in both datasets). We suspect that there are multiple factors contributing to this finding. First, as discussed above, the African cohort is far less powered in UKB, making it challenging to confidently fine-map variants in that cohort. Relatedly, the severe sample size imbalance biased UKB results toward reporting European-specific effects. Second, although we considered African vs. largely European cohorts in both datasets, there are substantial differences in ancestry across these cohorts. For example, both the African and non-African cohorts in the AoU analysis show high rates of admixture, with the African cohort having on average 15% European ancestry and the non-African cohort having on average 26% non-European ancestry. Third, the datasets each underwent different data preprocessing procedures resulting in different final sets of variants included in analysis (**Methods**). For example, a European-specific missense variant identified in *PCSK9* was only identified as an AS-V in UKB since it was filtered due to being tri-allelic in AoU (**Supplementary Fig. 20**). Finally, due to computational constraints we limited PIPSORT to considering a maximum of 3 causal variants at each trait-region. Many trait-regions harbor additional causal variants, and it is possible that PIPSORT identified true, but different, causal variants in AoU vs. UKB.

Our study faced several limitations that could be improved in future efforts. One challenge is the computational cost of exploring large numbers of causal configurations, which grows exponentially with the number of causal variants considered. As noted above, we restricted PIPSORT to explore configurations with up to 3 causal variants per locus in order to limit run time, but many signals likely have additional causal variants. We have implemented an optional feature to explore a user-defined subset of configurations as a partial solution, but run time could be further improved with a stochastic search process such as that implemented by FINEMAP^38^. For trait regions with a large number of causal variants, SuSiEx-style methods are still most suitable. An additional limitation of PIPSORT, as well as nearly all other fine-mappers, is that it assumes the true causal variant is included in the input. Here we considered only bi-allelic SNPs and indels, although the PIPSORT software supports fine-mapping with any type of variant whose genotypes can be represented as dosages and therefore should be capable of including e.g. linear effects of tandem repeat copy number on a trait^39,40^. Further, we only performed fine-mapping across two ancestries. The PIPSORT statistical framework can easily be extended to multiple studies, but this will come with increased computational cost.

In conclusion, we found that while the majority of GWAS signals are shared across ancestries, a substantial subset of signals are driven by ancestry specific variants and effects which are not directly modeled by most existing fine-mapping workflows. Compared to existing multi-ancestry fine-mapping methods, PIPSORT can detect the broadest range of scenarios by which ancestry-specific signals can arise and can best distinguish between shared and ancestry-specific signals. Finally, we highlight the need to continue to collect data from diverse cohorts, which is likely to continue to identify high-impact ancestry-specific causal variants. Ultimately, identification of these variants will lead to improved understanding of complex traits and will likely contribute to improved predictive performance of PRSs and other models.

## Methods

### PIPSORT method

PIPSORT performs fine-mapping jointly across two or more studies to take advantage of the improved resolution that comes primarily from differences in LD patterns across cohorts. As outlined in **Fig. 1**, PIPSORT takes as input variant-level association summary statistics (Z-scores) for a trait of interest and an LD reference panel for each study. It computes multiple posterior probabilities, including separate causal probabilities for each variant in each study and a global causal probability that the variant is causal in at least one study (see below).

We describe here the PIPSORT model when considering two studies, but this framework could be extended to consider an arbitrary number of studies. Assume we have *m*_*1*_ variants in study 1 and *m*_*2*_ in study 2. Let *m* be the number of variants in the union of the studies. The variants in both studies need not be the same, as some variants may be analyzed in both studies whereas others may be considered in only one. Let Σ_*i*_ with dimensions *m*_*i*_ × *m*_*i*_ be the LD matrix for the *i*^th^ study such that Σ_*i*_ [*j, k*] = *r*_*ijk*_ where *r*_*ijk*_ is the correlation between variants *j* and *k* in study *i*. Let *C*_*1*_ and *C*_*2*_ be binary vectors of length *m*_*1*_ and *m*_*2*_, respectively, consisting of causal configurations for each study, where *C*_*i*_[*j*] is set to 1 to model the *j*^th^ variant measured in study *i* as causal in that study. Unlike previous methods, this allows modeling a particular variant as causal in only one study, while still enabling PIPSORT to borrow information across studies. We consider an overall configuration vector *C* of length *m*_*1*_*+m*_*2*_ formed by concatenating *C*_*1*_ and *C*_*2*_. The construction of example configuration vectors is shown in **Supplementary Fig. 1**. The goal of PIPSORT is to compute the posterior probability of each potential causal configuration, and use the resulting values to compute variant-level posterior inclusion probabilities (PIPs).

Importantly, the number of potential causal configurations *C* is too large to exhaustively explore (*2*^|*C*|^). Thus, we only consider causal configurations with up to *k* unique causal variants across the two studies. In the case where the same *n* variants are considered in both studies and *k=2*, PIPSORT will evaluate 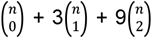 models. The 3 comes from the 3 possible models with one causal variant: (1) causal only in study 1, (2) causal only in study 2, or (3) causal in both. The 9 is similar but since there are two causal variants, we have these 3 settings for each. The number of configurations explored by PIPSORT is larger than the 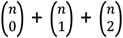 models that will be considered by MsCAVIAR^16^ and thus increases model evaluation time.

#### Computing the probability of a configuration

We can compute the posterior probability of a particular configuration *C* given the association summary statistics (Z-scores) available across all studies, denoted *D*, using Bayes’ Theorem:

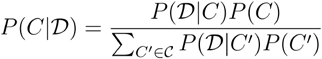

where *P(C)* represents the prior (see below). We follow MsCAVIAR in assuming Fisher’s polygenic model (linear and independent effects of multiple variants) and in modeling *D*|*C* by a multivariate normal distribution: 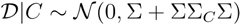 where Σ is a block LD matrix defined as follows:

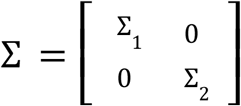

Σ_*C*_ is a matrix that depends on *C* (*C*=*C*_*1*_ |*C*_*2*_ as described above) and models the distribution of causal effect sizes and the heterogeneity of causal effect sizes across studies. The random effects model enables the heterogeneity of effect sizes by assuming that the effect size of variant *j* in study *i* is drawn from 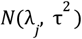 where *λ*_*j*_ is the overall non-centrality parameter for variant *j* and *τ* is the heterogeneity parameter. According to Fisher’s polygenic model, *λ*_*j*_ is drawn from *N*(0, σ)^*2*^ where σ^*2*^ is the global genetic variance. Given a *C*, we have all the information needed to construct Σ_*C*_. Following the example in **Supplementary Fig. 1**, if we have two studies, with two variants in the first and three in the second, and *C* = 11101 where variant 1 is a shared causal variant, variant 2 is ancestry-specific but present in both, and variant 3 is ancestry-specific but present only in study 2, then:

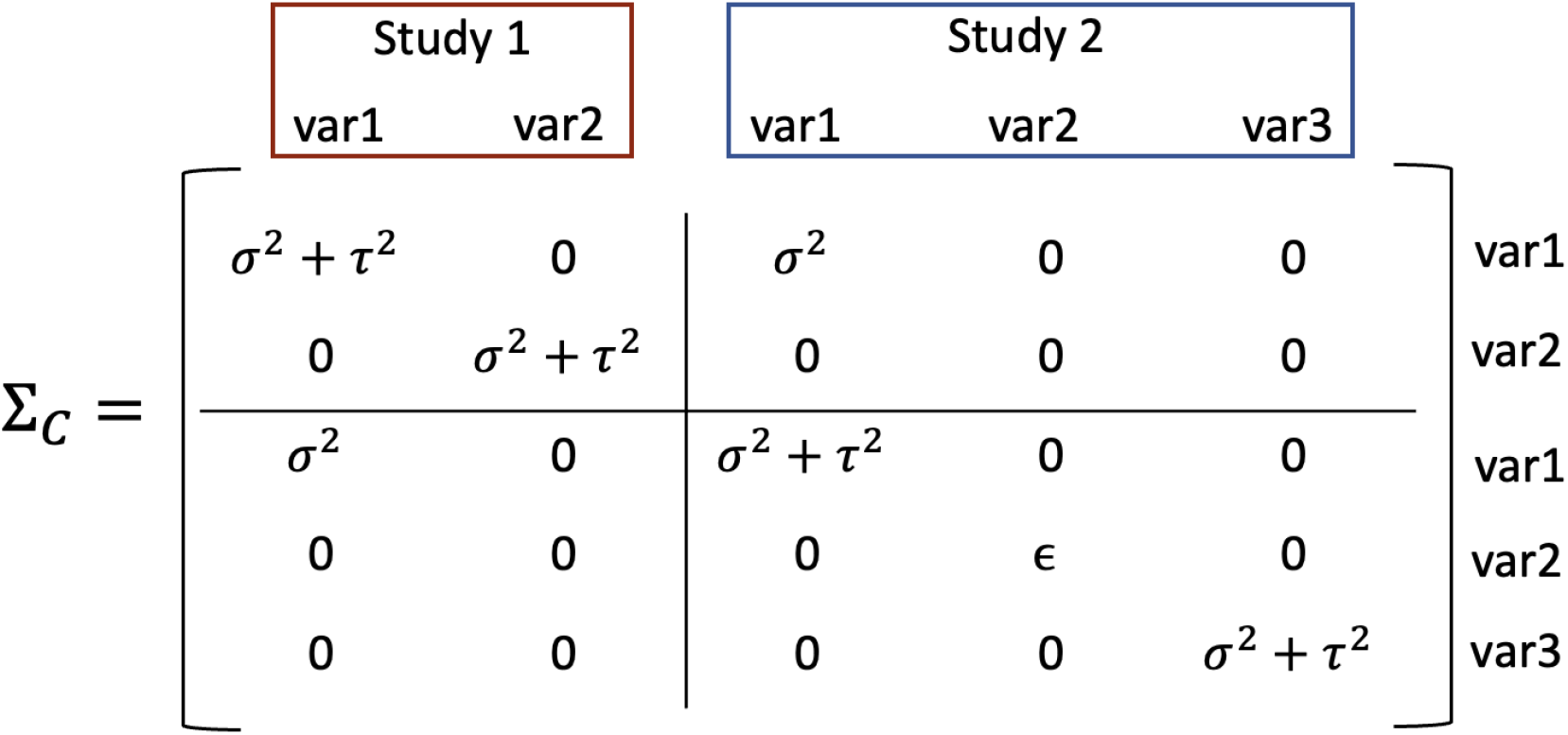

The diagonal term for non-causal (independent of causality in the other study) variants, is ϵ, a small constant to ensure full rank. The diagonal variance terms for causal variants include both Σ^*2*^ and *τ*^*2*^. As the study-specific effect sizes are modeled as independently drawn given the overall non-centrality parameter, the off-diagonal covariance terms are non-zero only for shared causal variants and include only σ^2^. Note, in this toy example, study 1 and study 2 have only 2 and 3 variants each whereas in real data, both studies would typically have hundreds of variants.

#### Constructing the prior

In MsCAVIAR and related methods, it is typical to model the prior probability of each configuration under the assumption that each variant is equally likely to be causal. Here we extend that prior to account for the sharing of signals across studies. We introduce a sharing parameter ρ to quantify the prior probability that a causal signal is shared across studies. A sharing parameter close to 1 favors configurations where the signals are shared whereas a value closer to 0 favors configurations where causal variants are study-specific. A sharing parameter of 0.5 does not favor either shared or study-specific signals.

Assume we have the same setup with two studies described above. Let 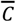 be a vector of length *m* such that 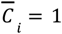 if variant *i* is set to 1 in either *C*_*1*_ or *C*_*2*_. Let *k* be the number of causal variants in the configuration, i.e. the number of positions set to 1 in 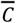. Then we can construct a vector *S* of length *k* where the j^th^ position of *S* is set to 1 if the *j*^th^ causal variant is causal in both studies and 0 if it is causal in only one study. That is, if the j^th^ causal variant corresponds to the i^th^ variant in the study, then if 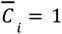 and *C*_*1i*_ = *C*_*1i*_ = *1* we set *S*_*j*_ = *1*. If 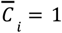 and *C*_*1i*_ ≠ *C*_*2i*_ then *S*_*j*_ = 0. We assume that every variant has a prior probability *γ* of being causal. Then the prior can be computed as follows:

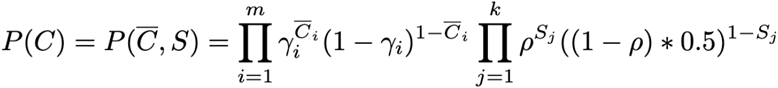

The first product incorporates the prior probability that each variant is causal, similar to existing fine-mapping methods, but accounting here for differing sets of variants and causal configurations across studies. The second product incorporates the amount of sharing expected between studies. For the *j*^th^ causal variant, the prior is multiplied by ρ when the variant is shared and by *1* − ρ otherwise. When a signal is not shared, it can be causal in study 1 and not in study 2, or it can be causal in study 2 and not in study 1. The 0.5 term in the second product accounts for this and ensures the priors for all configurations sum to 1. The prior is assumed to be uniform across all variants but the model could easily be extended to use a variant-specific prior, for example to incorporate functional annotations.

##### Relationship of sharing parameter to prior probabilities in related tools

MESuSiE and SuSiEx similarly incorporate prior probabilities that individual causal variants (MESuSiE) or credible sets (SuSiEx) are shared vs. ancestry-specific. For MESuSiE, users can set priors using a tuple of weights indicating the expected ratio of variants that are specific to study 1, specific to study 2, or shared. Our results on simulated and real data show that the default MESuSiE setting of these weights to (3,3,1) is similar to setting the PIPSORT sharing parameter to 0.25. SuSiEx assumes all configurations are equally likely and so does not enable users to adjust the prior that signals are shared. We similarly found that SuSiEx settings are similar to setting the PIPSORT sharing parameter to 0.25. In contrast, PIPSORT’s default sharing parameter is set to 0.75 to reflect the high degree of causal variant sharing^7^ across populations.

#### Posterior inclusion probabilities (PIPs)

Because PIPSORT considers configurations that allow the causal signals to differ across studies, we can compute study-specific PIPs. For variant *j* in study *i*, the study-specific PIP is:

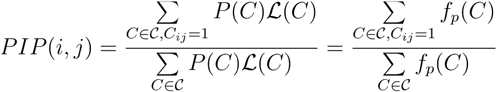

where ℒ(*C*) is *P*(𝒟|*c*) as defined above and the posterior probability *f*_*p*_(*C*) = *P*(*C*)ℒ(*C*). We can also compute other posterior probabilities that provide different causal information, summarized below. For ease of reading, we provide only the numerator, as the denominator is always the same as in the formula above. Some of the formulas as well as our implementations are specific to two studies, but could be generalized to more than two studies.

PIPSORT computes the following PIPs and related probabilities:

- *Study-specific PIP*: The probability that variant *j* is causal in study *i* is computed as 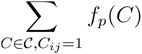
- *Global PIP*: The probability of variant *j* being causal in any study is computed as 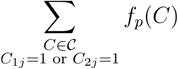
- *Shared PIP*: The probability that variant *j* is causal in both studies is computed as 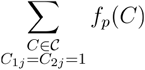
- *Not-shared PIP*: The probability that variant *j* is causal in only one study is computed as 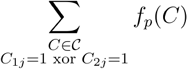
- *No causal*: The probability that study *i* has no causal signals is computed as 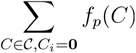

#### Specifying a set of configurations to test

PIPSORT explores an exhaustive search space, which comes with runtime limitations and can become intractable at loci with large numbers of variants or with many causal signals. Other fine-mapping methods such as FINEMAP^38^ have evaluated techniques including stochastic search for reducing the search space. While we do not currently incorporate this strategy, PIPSORT does offer an optional mode in which the user can limit the search space by providing as input a set of causal configurations to explore. As a starting point, we provide a script in the PIPSORT repository for computing a smaller set of causal configurations by using a conditional regression procedure to narrow down the search space.

#### Accounting for different sample sizes across studies

In the same manner as MsCAVIAR, PIPSORT models the effect size to account for different sample sizes across studies. This is done by setting Σ^*2*^ in Σ_*C*_ to 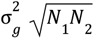 where *N*_*1*_ and *N*_*2*_ are the sample sizes for the studies 1 and 2 respectively and effect sizes are modeled as 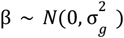. As done in MsCAVIAR^16^, we set the default of 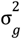 such that 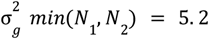 for all variants where 5.2 is the absolute Z-score corresponding to the traditional genome-wide significance level of 5e-8.

### Simulation experiments setup

We used haptools^41^ v0.4.0 to simulate a phenotype given the desired effect size(s) and causal variant(s) based on real genotypes from individuals in the UK Biobank^21^ corresponding to the UKB EUR and AFR cohorts described below (see ***Sample and cohort preprocessing***). Dosages in pgen were first converted to hard call genotypes in bgen using the plink flag `--hard-call-threshold 0.4999`. The bgen was then converted to a vcf with the plink flag `--recode vcf` which was passed as input to haptools for phenotype simulation. We used PLINK 2.0^42,43^ to extract loci, compute LD matrices, and compute GWAS summary statistics. There is a significant size imbalance between these two cohorts. To understand the impact of sample size, we conduct experiments in which we randomly downsampled the EUR cohort to match the size of the AFR cohort. Experiments conducted with the full and downsampled cohorts are labeled as *unbal* (unbalanced) and *bal* (balanced), respectively.

For the first simulation experiment with a single causal variant, we chose a SNP with a specified minor allele frequency (MAF) at random to simulate as causal and used haptools to obtain the simulated quantitative phenotype for a range of effect sizes. MAFs ranged from 0.001-0.25, effect sizes ranged across the set {0.01, 0.05, 0.08, 0.1, 0.2}, and the sharing parameter ranged across {0.01, 0.25, 0.5, 0.75, 0.99}. We approximated corresponding sharing parameters (the ancestry weight parameter) for MESuSiE as {c(5,5,1), c(3,3,1), c(1,1,1), c(0.5,0.5,1), c(0,0,1)}. Separately for the EUR and AFR cohorts, we computed association summary statistics with plink flags `--rm-dup force-first --maf 0.001 --linear allow-no-covars` and LD matrices with plink flags `--r-unphased square ref-based` (additionally adding `bin4` for SuSiEx) for all variants in a 200Kb window centered at the simulated causal SNP, which were used as input to PIPSORT, SuSiEx v1.1.2, and MESuSiE. For simulation experiments, we did not use covariates. We performed five trials of each simulation setting, in each case varying the seed passed to haptools to simulate the phenotype. We used the same setup for simulation with a single causal AS-V, but downsampled the EUR cohort further to 100k samples for the unbalanced setting and focused on a smaller set of parameters, using a sharing parameter of 0.25 and an effect size of 0.08. The variant rs11591147 was simulated as a causal AS-V in EUR and rs28362263 in AFR as these have the necessary MAFs to be AS-Vs in their respective cohorts.

For the second simulation experiment with two causal SNPs, performed in PIPSORT only, we evaluated multiple settings with: (1) both SNPs as shared, (2) one as shared, one as ancestry-specific, and (3) both as ancestry-specific. For simplicity, the effect size was set to 0.2 for both SNPs and we looked at sharing parameters 0.25 and 0.75. The region for fine-mapping starts 100Kb before the minimum causal SNP position and ends 100Kb after the maximum causal SNP position. We again performed five trials for each simulation setting. The two causal SNPs were chosen mostly at random but were also chosen to model the LD structure setting in which the two causal SNPs are neither in strong LD with each other nor commonly in strong LD with any other variant in the region. Both variants are common (0.01≤MAF≤0.07) and in low LD (r^2^<0.001) with each other in the UKB EUR and AFR cohorts.

### Preprocessing UK Biobank and All of Us data

#### Variant data

##### UK Biobank

We used 93,095,623 autosomal SNPs and indels imputed by the UKB team^44^. Briefly, they genotyped 670,739 autosomal SNPs and indels using a microarray, filtered variants not passing quality control measures and then phased those variants into haplotypes. Into those haplotypes they imputed SNPs and indels from the Haplotype Reference Consortium, UK10K and 1000 genomes phase 3 reference panels, resulting in 93,095,623 total genotyped variants. For QC, we applied a minor allele frequency threshold to these variants at a later step (see below). We followed preprocessing steps described in Margoliash et al.^39^ to encode imputed genotypes as non-reference allele dosages in plink pgen format.

##### All of Us

We used the short-read whole genome sequences (srWGS) v7.1 ACAF dataset consisting of 99,250,816 SNPs and indels (at 48,314,438 sites) with allele frequency greater than 1% or allele count greater than 100 in any computed ancestry subpopulation^45^.

#### Sample and cohort preprocessing

##### UK Biobank

We use the sample quality control (QC) process from Margoliash et al.^39^ to obtain a maximal set of White British participants and a maximal set of Black (see below) participants that have been previously filtered^39^ for excessive genetic relatedness, sex discrepancies, withdrawn participants, and sex chromosome aneuploidies. The ancestry of the White British participants was determined by the UKB team and their results can be found in UKB Data Showcase Resource ID 531. Black participants consist of samples with African and Caribbean ancestry as denoted by the codings 4, 4001, 4002, and 4003. Additional participants are filtered due to phenotype preprocessing steps, resulting in 334,324 EUR and 6,771 AFR participants for platelet count and 327,817 EUR and 6,684 AFR participants for LDL.

##### All of Us

The AoU research program provides a list of participants to remove to obtain a maximal set of unrelated individuals, which we used to filter for genetic relatedness. All available samples passed the sex concordance check performed by the AoU research program. We used samples that self-report “male” or “female” for sex assigned at birth. We performed additional QC filters specific to WGS as similarly done by prior work on AoU^46^: for each sample, we compute the transition/transversion ratio, heterozygous/homozygous ratio, insertion/deletion ratio, and number of singletons and filtered samples whose value for any of these metrics falls outside 8 standard deviations of the mean across all samples. We use two cohorts, a NOT AFR cohort and an AFR cohort. The NOT AFR cohort consists of samples whose genetically inferred ancestry is not “afr” and self-reported race is not “Black or African American.” The AFR cohort consists of samples whose genetically inferred ancestry is “afr” and self-reported race is “Black or African American.” The NOT AFR cohort contains 174,829 samples and the AFR cohort contains 44,384. Additional samples are filtered due to phenotype preprocessing steps, resulting in 98,663 NOT AFR and 19,959 AFR samples for platelet count and 56,057 EUR and 10,699 AFR samples for LDL. Some analyses used two additional AOU cohorts: AoU EUR and AoU ALL. AoU EUR (n=119,178) consists of samples that have self-identified race as White and genetically inferred ancestry as EUR and AoU ALL (n=225,116) consisting of all samples that pass QC.

#### Global ancestry analysis in *All of Us*

We first constructed a reference panel for global ancestry inference using a total of 2,418 samples from the 1000 Genomes Project^47^ and Human Genome Diversity Project^48^ (HGDP). Genotype files for 1000 Genomes samples and for HGDP were obtained from the plink2 resources webpage^42^. The reference samples were labeled according to six ancestry groups: European (GBR, FIN, IBS, CEU, TSI), African (ACB, GWD, ESN, MSL, LWK, ASW, YRI), South Asian (PJL, BEB, STU, ITU, GIH), and East Asian (CHS, CDX, KHV, CHB, JPT) in 1000 Genomes and West Asian (Druze, Bedouin, Palestinian), and Admixed American (Surui, Maya, Karitiana, Pima) in HGDP. We then used plink2^42^ to subset the srWGS ACAF dataset described above to filter variants with call rates below 5%, minor allele frequencies below 5%, and those deviating from Hardy-Weinberg equilibrium (p<1×10^-6^). We then used the plink2 indep-pairwise function to select independent SNPs using a window size of 50Kb, step size of 2 variants, and an LD r^2^ threshold of 0.2 and merged pruned variants from AoU and the reference panel files, retaining 83,922 shared variants for downstream analysis. We used plink2 to perform PCA on the reference samples and AoU samples to retrieve the first 10 principal components. The resulting principal components were used as input to Rye^24^ v0.1 to infer global ancestry proportions with non-default options pcs = 10, round = 100, and inter = 100.

#### Phenotype preprocessing in UKB and AoU

We performed GWAS followed by fine-mapping using PIPSORT in UKB and AoU on two quantitative phenotypes: LDL-C,and platelet count.

##### UK Biobank

We obtained data from fields 30780 and 30080 for LDL and platelet count, respectively. For each trait, we dropped samples that were missing measurements across all patient visits. We added a binary covariate(s) to denote from which visit the measurement was taken, preferring the measurement from the earliest visit when measurements from multiple visits are available. For LDL, we one-hot encoded aliquot information (field 30782 for LDL) as binary covariates. For platelet count, we one-hot encoded device information (field 30083) as binary covariates. For LDL, we additionally included two binary covariates to encode statin use at either of the two measurement collections. Field 20003 contains medication information, which we used to construct the statin use covariates, encoding use of any of seven statins (simvastatin, fluvastatin, pravastatin, eptastatin, velastatin, atorvastatin, rosuvastatin) at either visit. If any categorical covariate value was not obtained by at least 0.1% of samples, we dropped all samples with this covariate value.

##### All of Us

For platelet count, we obtained data corresponding to concept ID 3024929 (“Platelets [#/volume] in Blood by Automated count”). Data points with a unit other than “thousand per microliter”, “thousand per cubic millimeter”, or “billion per liter” were removed. We additionally kept data points with “no value” listed as the unit since these followed a similar distribution to the retained measurements. For LDL cholesterol, we obtained data corresponding to concept ID 3028288 (“Cholesterol in LDL [Mass/volume] in Serum or Plasma by calculation”). Data points with a unit other than “milligram per deciliter” were removed. For all traits, after restricting to data points from individuals passing sample preprocessing above, we filtered values more than 5 standard deviations away from the mean. We then obtained the median phenotype value per year that measurements were available, then took the median of medians across all years. We recorded the year corresponding to the final value used as the age covariate for downstream association testing. For LDL-C, we additionally obtained data from concept ID 21601855 (exposure to HMG-CoA reductase inhibitors) and encoded a binary covariate indicating if the phenotype value used overlapped statin usage.

### Association testing in AoU and UKB

*UK Biobank*: We used PLINK 2.0^42,43^ to compute association statistics using the flags `--rm-dup exclude-all --maf 0.01 --linear hide-covar --covar-variance-standardize`. All traits used age, sex, and 10 ancestry principal coordinates (PCs) as covariates. Phenotype values were rank inverse normalized. The PCs were provided by the UKB team (field 22009).

*All of Us*: We used Hail^49^ 0.2 to compute association statistics. All traits used age, sex, and 10 ancestry principal coordinates (PCs) as covariates. Phenotype values were rank inverse normalized. The PCs were provided by the AoU research program. Prior to association testing, we filtered the ACAF dataset described above to exclude genotypes for which the filter field was not set to “PASS” or the genotype quality was less than 20, samples with call rate <0.9, and variants with (1) more than two alleles, (2) call rate <0.9, (3) overall minor allele frequency (MAF) ≤0.01, or (4) dramatic departures from Hardy-Weinberg equilibrium (p≤1e-100). The Hail function “linear_regression_rows” was used to perform association testing.

### Identifying trait-regions for fine-mapping in AoU and UKB

To enable comparison across the UK Biobank and *All of Us* datasets, we jointly constructed loci for fine-mapping. For each trait, we have 4 GWASs: UKB with the EUR cohort, UKB with the AFR cohort, AoU with the NOT AFR cohort, and AoU with the AFR cohort. For each GWAS, we first identify peaks of width 250Kb centered on lead variants using the peak detection method described in “Defining significant peaks” in Margoliash *et al*.^39^ For the EUR and NOT AFR cohorts, we required lead variants to pass the standard p<5e-8 genome-wide significance threshold. For the AFR cohorts, we relaxed this threshold to 1e-6 due to the substantial loss of power resulting from lower cohort sizes. UK Biobank data is provided in hg19 reference coordinates whereas AoU uses the hg38 reference assembly. To account for this, UKB peaks were converted to hg38 using the UCSC Table Browser^50^. While the majority of UKB peaks were successfully converted to hg38 coordinates, some (platelet count: 1/25 for AFR, 41/418 for EUR, LDL: 1/12 for AFR, 28/272 for EUR) could not be and were excluded from downstream analyses.

Contiguous regions for fine-mapping were constructed by applying the following steps to the union of peaks across all 4 GWASs: starting with any peak variant not already assigned to a region, we repeatedly add unassigned peak variants to this region until there does not exist an unassigned peak variant within 500Kb of any of the peak variants in this region. This procedure is repeated until all peak variants are associated with a region. The fine-mapping region starts at 500Kb before the minimum variant position and ends at 500Kb after the maximum variant position. If any region extends beyond the start or end of a chromosome, we cap the region at the appropriate chromosome coordinates. As in Margoliash *et al*.^39^, we excluded the MHC region (hg38:chr6:28,510,120-33,480,577) as it is challenging to fine-map. The resulting regions are in hg38 coordinates. We used UCSC LiftOver^51^ to convert the regions to hg19 coordinates for fine-mapping in UK Biobank. The vast majority of trait-regions successfully lifted over from hg38 to hg19. Regions (2/145 for LDL, 5/284 for platelet count) that failed liftover were excluded from downstream analysis.

### Running PIPSORT in AoU and UKB

We ran PIPSORT to perform two-study fine-mapping separately in the UK Biobank (EUR vs. AFR) and *All of Us* datasets (NOT AFR vs. AFR). In each case, we first jointly pruned variants to exclude those not passing a p-value threshold of 0.0001 in either of the two cohorts considered. We used PLINK 2.0^42,43^ flags `--r-unphased square ref-based` to compute LD matrices separately for the two cohorts. We used the GWAS results to compute Z-scores for each variant by dividing the effect sizes by the standard error. The Z-scores and LD matrix for each study were passed to PIPSORT as input. For both datasets we ran PIPSORT twice on each trait-region, once with a sharing parameter of 0.25 and once with 0.75.

To determine the maximum number of causal variants to consider at a locus, we first ran a conditional regression procedure. For each trait-region in each of the two cohorts, we repeatedly ran association studies (up to a maximum of 15), each time adding the most significant variant as a covariate until none of the remaining variants had a P-value less than a certain threshold (5e-8 in EUR/NOT AFR and 1e-6 in AFR). We used the maximum number of variants added as covariates during the conditional regression procedure in either cohort to set the maximum number of causal variants to consider with PIPSORT. Due to computational limitations, we limit the maximum number of causal variants to 3. We further limit the maximum number of causal variants at 1 if either study has more than 1700 variants and at 2 if either study has more than 325 variants. For trait-regions in which either study has more than 3000 variants, we do not run PIPSORT as very large studies would exceed the runtime (24 hours) and memory (64 GB) limits we set per trait-region. This excluded 1 trait-region in AoU and 3 in UKB across all traits. We also do not run PIPSORT on loci where both studies do not have any significant variants or either study has less than 25 variants remaining after the joint variant processing described in this section. This excluded 129 AoU LDL, 171 AoU platelet count, 16 UKB LDL, and 26 UKB platelet count trait-regions.

### Running SuSiEx in AoU

We ran SuSiEx^14^ v1.1.2 to perform two-study fine-mapping in the AoU dataset (NOT AFR vs. AFR). We did not prune any variants from the studies. We used PLINK 2.0^42,43^ flags `--r-unphased square bin4 ref-based` to compute LD matrices separately for the two cohorts. The summary statistics and LD matrix for each study were passed to SuSiEx as input. We ran SuSiEx with `--n_sig=3 --level=0.8 --min_purity=0`.

We considered any locus for which PIPSORT identified an AS-V to also be identified by SuSiEx if at least one candidate AS-V variant returned by PIPSORT had an overall SuSiEx PIP>0.1 and the post-hoc ancestry-specific probability of the credible set in the ancestry where the variant is polymorphic was >0.9.

### Running MESuSiE in AoU

We ran MESuSiE^17^ to perform two-study fine-mapping in the AoU dataset (NOT AFR vs. AFR). We first pruned variants to include only those that appear in both studies. We used PLINK 2.0^42,43^ flags `--r-unphased square ref-based` to compute LD matrices separately for the two cohorts. The summary statistics and LD matrix for each study were passed to MESuSiE as input. We ran MESuSiE with `L=3` and `ancestry_weight=c(3,3,1)`. The ancestry weight of `c(3,3,1)` was used when running MESuSiE on real data as simulation experiments showed this most closely matches PIPSORT with a sharing parameter of 0.25.

### Fine-mapping in the PGC schizophrenia dataset

We ran PIPSORT, SuSiEx, and MESuSiE to perform two-study fine-mapping using the summary statistics provided by the PGC for GWAS performed separately in the EAS and EUR populations for schizophrenia^52^. We followed a similar process as described above to construct trait-regions. For peak identification, we required lead variants in either population to pass the standard p<5e-8 genome-wide significance threshold. As we did not have access to individual-level statistics, we used the 1000 genomes dataset^47^ to compute LD matrices and the number of peak variants added to a trait-region to set the maximum number of causal variants to consider. This required additional filtering of variants to restrict to those common to both the PGC schizophrenia GWAS and 1000 genomes. We applied this filtering separately in each population. 1 SCZ trait-region was excluded due to memory and runtime constraints, 4 did not pass our criteria for the number of (significant) variants.

### Identifying high confidence AS-Es

To obtain a list of high confidence AS-Es, we applied stricter thresholds, restricting to trait-regions (1) in which one population has a high probability (>0.3) of not having any causal variants, (2) where the sum of the probability difference between candidate AS-E variants being not shared vs. shared is greater than 0.2, and (3) where the sum of candidate AS-E variant PIPs is at least 0.5.

### Variant and gene annotation

We used the Ensembl REST API^53^ to obtain variant consequences annotated by Variant Effect Predictor (VEP)^54^ based on Ensembl release 113. We annotated variants with the most severe VEP consequence. Candidate *cis* regulatory regions identified by ENCODE^25^ were obtained from the UCSC Genome Browser^51^ ENCODE cCREs track. We used the web version of locus zoom^55^ (my.locuszoom.org) and uploaded summary statistics in build GRCh38 to obtain gene annotations at a trait-region for figures.

### Local ancestry inference and analysis

Local ancestry inference was performed with GNOMIX^56^. Full details of local ancestry inference are provided in Gonzalez Rivera *et al*. (manuscript under consideration).

For stratified regression analysis, we used 4 cohorts in AoU. In addition to the AoU NOT AFR and AFR cohorts already used for fine-mapping, we included AoU EUR and AoU ALL. For the candidate AS-E on which we performed stratified regression, we first identified the corresponding Gnomix local ancestry region. For each cohort, we stratified samples by those that are homozygous for non-European local ancestry, heterozygous for European local ancestry, and homozygous for European local ancestry at this region. We performed 12 association tests for each candidate AS-E, testing the 3 local ancestry groups in the 4 cohorts separately. This analysis was performed in AoU only.

To test for an interaction between a candidate AS-E and local ancestry in the GNOMIX region corresponding to the AS-E, we first computed a local ancestry covariate that was 0 for samples homozygous for non-European local ancestry, 1 for samples heterozygous for European local ancestry, and 2 for samples homozygous for European local ancestry at that region. Then we performed association testing with the candidate AS-E at that gnomix region by first regressing the phenotype on age, sex, and the first 10 PCs (and the statin-use covariate for LDL). The residuals from this regression were then regressed on the genotype of the candidate AS-E as well as an interaction term between the genotype and the local ancestry covariate for that region. This analysis was performed in AoU only.

## Data availability

This study used data from the *All of Us* Research Program’s Controlled Tier Dataset V7.1, available to authorized users on the Research Workbench. We used genetic and phenotype datasets provided by UK Biobank under application number 46122 (see https://www.ukbiobank.ac.uk/use-our-data/apply-for-access).

## Code availability

The PIPSORT tool is available on Github: https://github.com/cast-genomics/pipsort

AoU workflows for this project are available on Github: https://github.com/cast-genomics/cast-workflows

## Author contributions

T. M. developed PIPSORT, performed analyses, and wrote the manuscript. N. M. helped perform phenotype preprocessing and with pipeline setup in the AoU dataset. J. M. helped perform GWAS analyses in UKB. W. G. G. R. performed local ancestry inference and global ancestry analysis in AoU. T.A., K.A.F., and A.G. helped write the manuscript. M. G. supervised the study and wrote the manuscript.

## Acknowledgements

This research was supported in part by NIH/NHGRI grants R01HG010885 (M.G. and A.G), RM1HG011558, (M.G and K.A.F) and U01HG013442 (M.G). Our work used the UK Biobank Resource under application number 46122 and data provided by patients and collected by the NHS as part of their care and support. W.G.G.R was supported by the Alfred P. Sloan Foundation Minority Ph.D. (MPHD) Program [G-2022-10127] and NIH grant T15LM1127113. We gratefully acknowledge *All of Us* and UK Biobank participants for their contributions, without whom this research would not have been possible. We also thank the National Institutes of Health’s *All of Us* Research Program for making available the participant data examined in this study. We additionally thank Arya Massarat for many helpful discussions, and Matteo D’Antonio and Eric Mendenhall for helpful comments.

