## Supplementary Figures for "Characterization of shared and ancestry-specific signals driving complex traits using multi-ancestry fine-mapping"

#### Supplementary Fig. 1

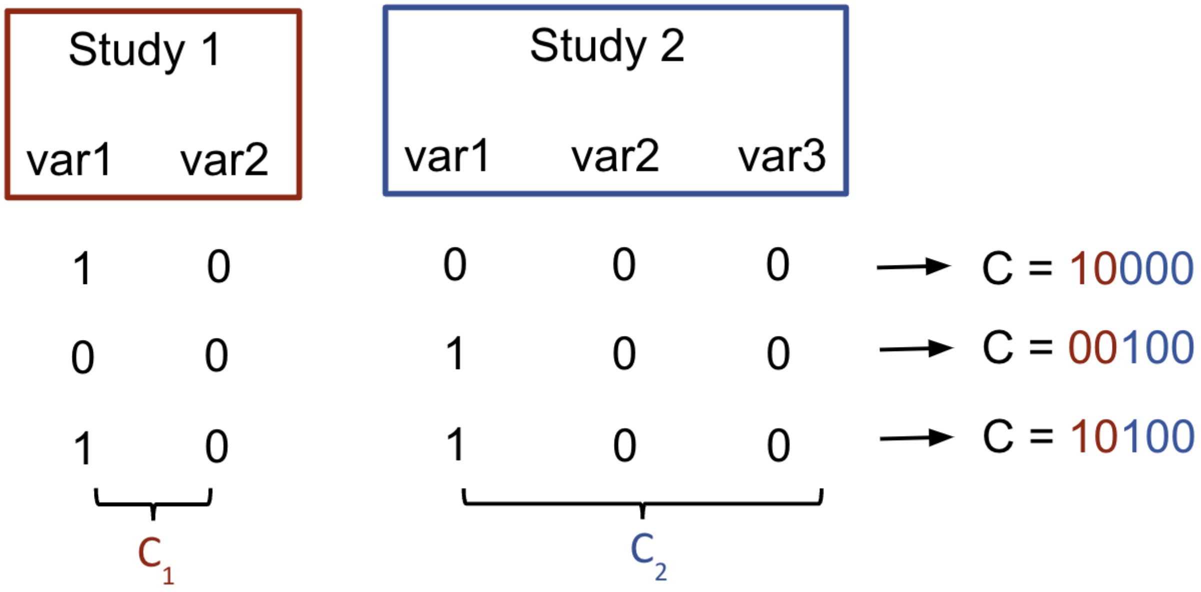


**Example PIPSORT configurations.** In this example, we have two studies. Study 1 consists of variants var1 and var2. Study 2 consists of var1, var2, and var3. For a particular configuration vector *C*, the first two numbers in red represent the causal setup for Study 1 and the last three numbers in blue represent Study 2. The figure shows the three configuration vectors that are generated when exploring var1 as the only causal variant in either study. If we explore a maximum of *k*=2 causal variants, the set of all configurations that we explore is {00000, 00001, 01000, 00010, 01010, 10000, 00100, 10100, 10101, 10001, 00101, 11110, 10110, 01110, 11010, 11100, 11000, 00110, 01100, 10010, 01011, 00011, 01001}. As another example, configuration 10101 models var1 as causal in both studies and var3 as causal in study 2. While there are three positions set to 1, the three positions correspond to two unique variants.

Supplementary Fig. 2

**a**

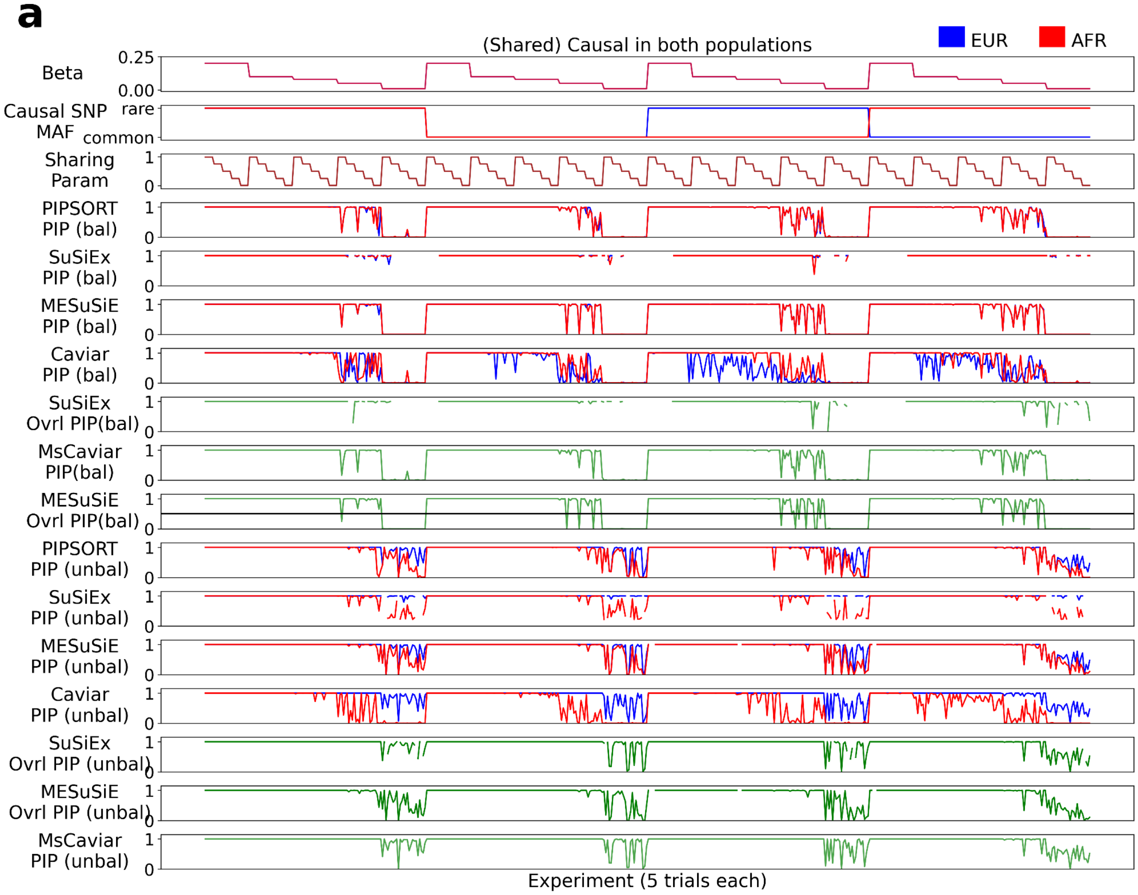


**b**

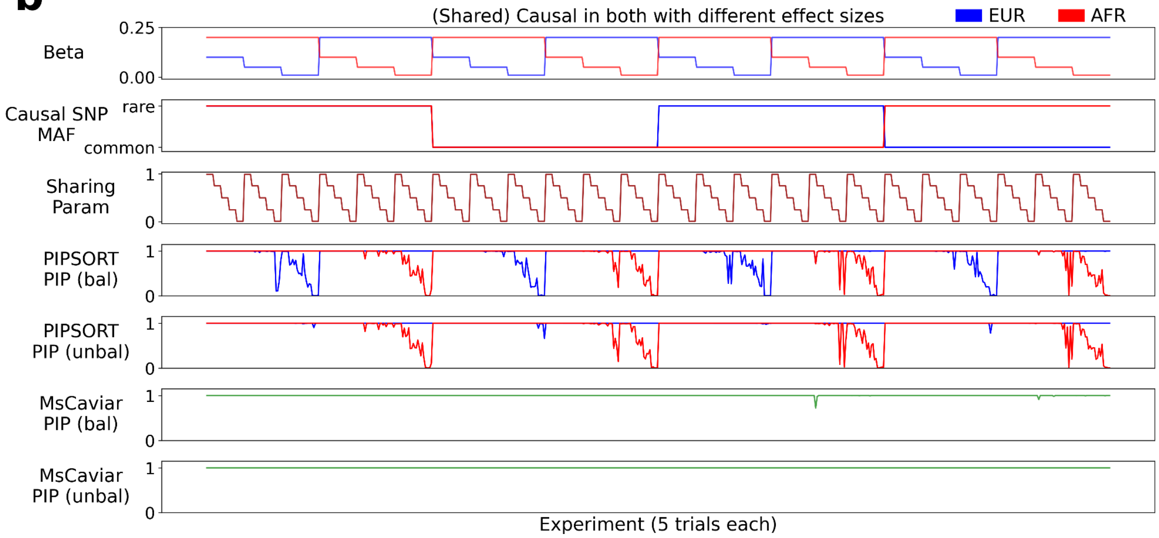

**Evaluating PIPSORT using simulated data with a single shared causal variant. a. Simulations with the same effect size in both populations.** Simulations are based on European and African cohorts from the UK Biobank (see **Methods**). The top top panels show simulation parameters (1st from top=effect size; 2nd=causal SNP minor allele frequency) for each simulation run along the x-axis. Red and blue lines denote simulation parameters for the African and European populations, respectively. The third panel shows the sharing parameter input to PIPSORT. The next 7 panels show fine-mapping results from PIPSORT and other methods with balanced population sizes, and the bottom 7 show fine-mapping with the full (unbalanced) population sizes available in the UK Biobank. PIPSORT PIP denotes population-specific PIPs returned by PIPSORT. SuSiEx PIP denotes the post-hoc population-specific probability computed for the credible set. MeSuSiE PIP denotes the population-specific PIP returned by MeSuSiE. SuSiEx Ovrl PIP denotes the PIP returned by SuSiEx prior to post-hoc estimation of population-specific PIPs. MeSuSiE Ovrl PIP denotes the probability that the variant is a shared signal across both populations. Caviar PIPs denote PIPs returned under single-cohort fine-mapping using CAVIAR. MsCaviar PIPs, shown in green, denote the single PIP returned from performing multi-ancestry fine-mapping with MsCAVIAR. **b. Simulations in which the effect size varies across populations.** Panels are the same as in **a.**, with the exception that CAVIAR, MeSuSiE, and SuSiEx were not included in the analysis.

### Supplementary Fig. 3

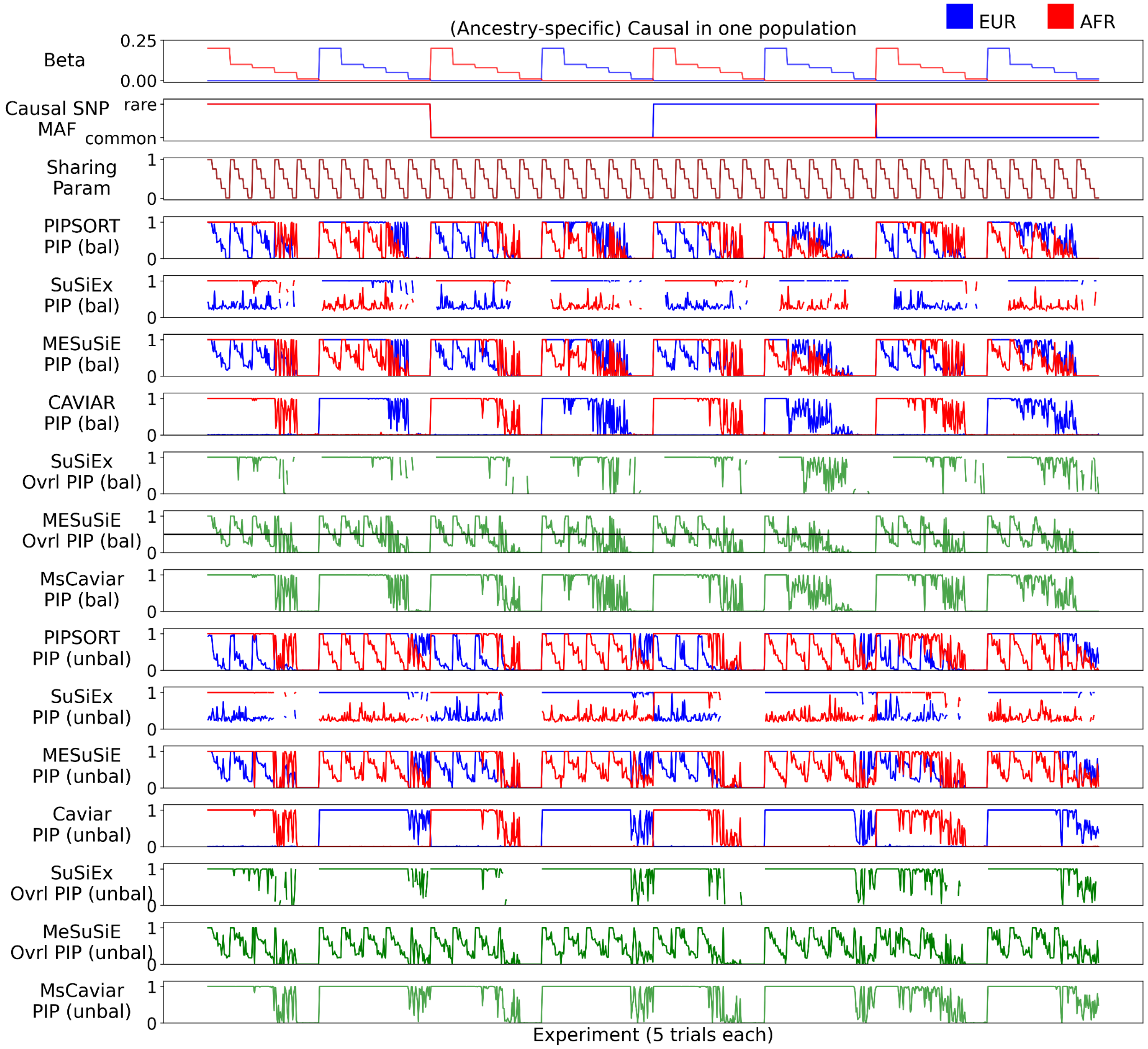


**Evaluating PIPSORT using simulated data with a single causal variant with an ancestry-specific effect.** Panels are the same as in **Supplementary Fig. 2**. Whereas **Supplementary Fig. 2** shows simulations with shared causal variants, this figure shows simulations where the effect sizes vary across ancestries.

Supplementary Fig. 4

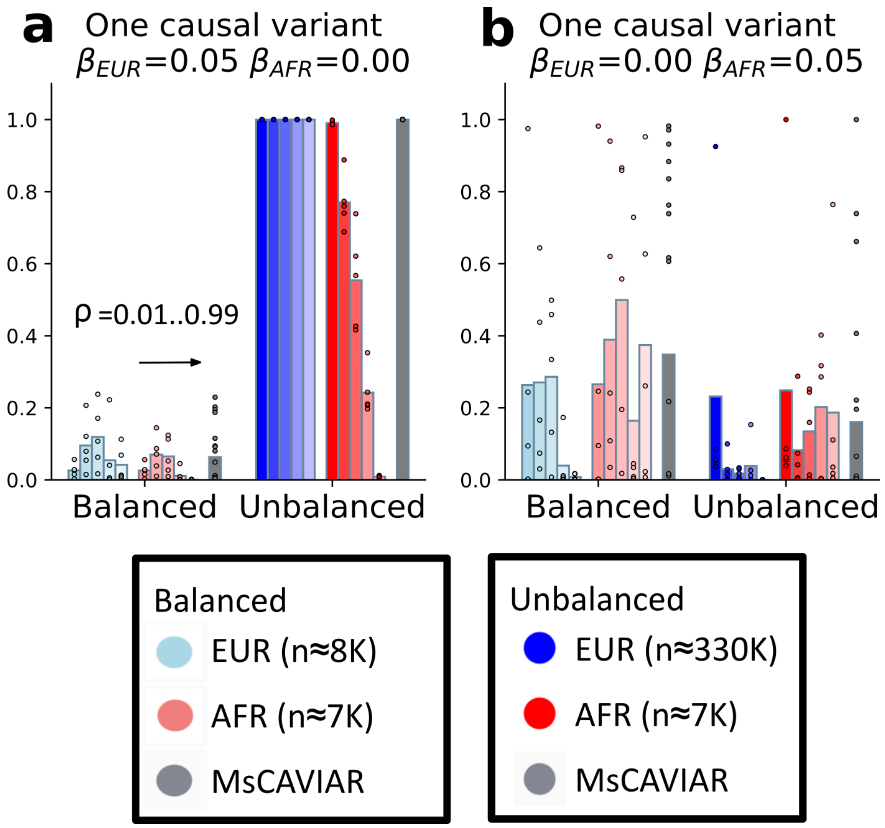


**Simulation results with a single causal variant.** Barplots show the PIPs for simulations in which either the causal variant is European-specific (**a**) or African-specific (**b**). In each panel, left bars show results for balanced cohort sizes and right bars show results for the full (unbalanced) cohort sizes. Bar shading from left to right indicates simulations with different values of the sharing parameter (0.01, 0.25, 0.5, 0.75, 0.99). PIPs denoted by each bar are averaged across 5 simulations. The underlying values for each simulation are shown as dots. For comparison, results for MsCAVIAR, which returns only a single overall PIP, are shown in gray. Bars for MsCAVIAR are averaged across 25 simulations.

### Supplementary Fig. 5

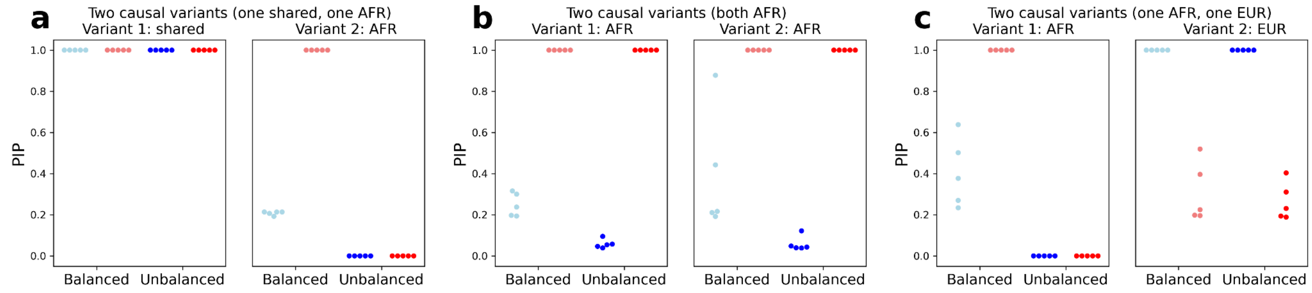


**Simulation results with two causal variants.** Plots show study-specific PIP values for scenarios in which: one variant is shared and the other is African-specific (**a**), both variants are African-specific (**b**), and one is African-specific and the other is European-specific (**c**). For each scenario, results from 5 independent simulations with sharing parameter 0.25 are shown and causal variants are simulated with an effect size of 0.2. For **a-c** light blue=European cohort downsampled 8000 samples; dark blue=full European cohort; light and dark red=full African cohort.

### Supplementary Fig. 6

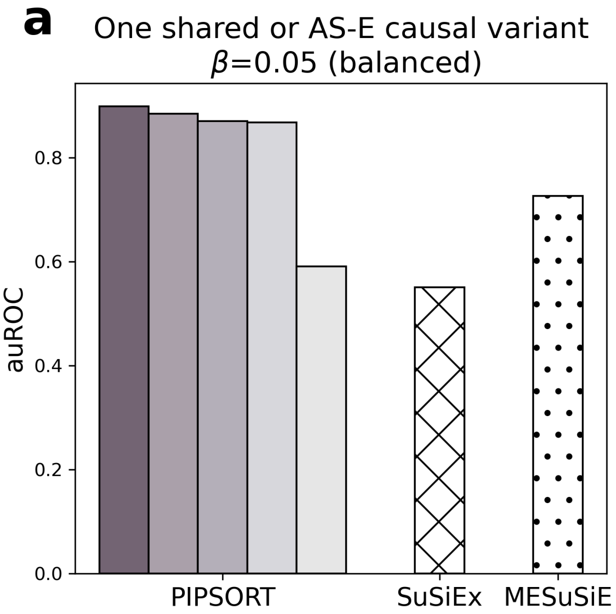


**auROC for predicting an AS-E vs. shared variant using PIPSORT, SuSiEx, and MeSuSiE.** This figure is the same as **Fig. 2d** except for the balanced simulation setting. Each bar depicts the area under the receiver operator curve (auROC) for using the difference in ancestry-specific PIPs to predict if a causal variant is an AS-E vs. shared signal. Each bar is computed from 300 simulations: 100 that simulate a EUR AS-E, 100 that simulate an AFR AS-E, and 100 that simulate a shared AS-E, all with an effect size of 0.05. For PIPSORT, bar shading from left to right indicates simulations with different values of the sharing parameter (0.01, 0.25, 0.5, 0.75, 0.99). For MESuSiE, we used ancestry weights w=(3,3,1), which indicates a 3:3:1 ratio of signals that are present in EUR-only, AFR-only, or both and performs similarly to setting the PIPSORT sharing parameter to 0.25.

### Supplementary Fig. 7

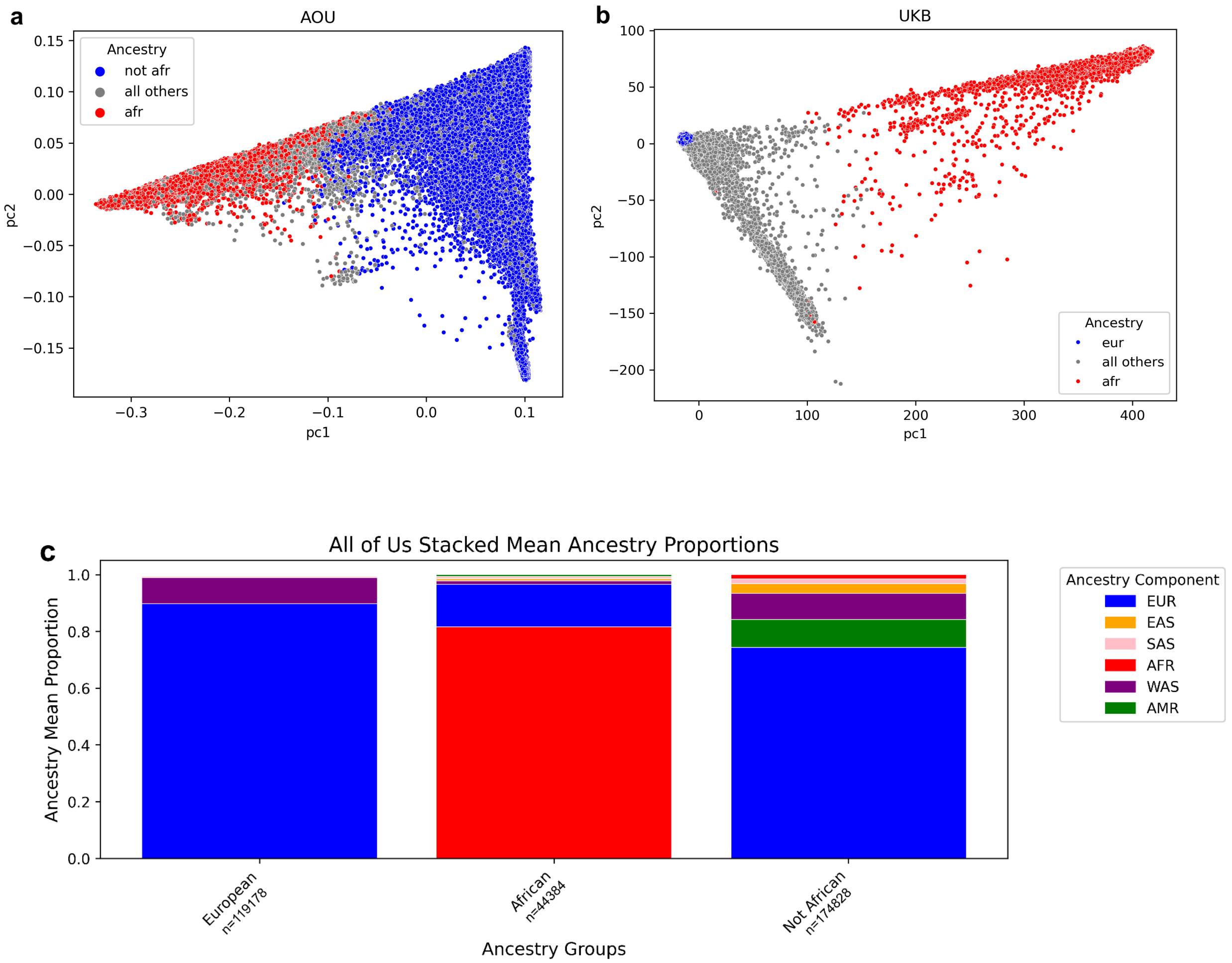


| | EUR | EAS | SAS | AFR | WAS | AMR |
|---|---|---|---|---|---|---|
| European | 0.898 | 0.001 | 0.005 | 0.002 | 0.091 | 0.002 |
| African | 0.15 | 0.008 | 0.007 | 0.815 | 0.013 | 0.006 |
| Not African | 0.743 | 0.034 | 0.017 | 0.015 | 0.093 | 0.098 |

**Global ancestry analysis of AoU and UKB cohorts used for fine-mapping.** The first two genetic principal components are plotted for AoU in **(a)** and for UKB in **(b).** The AOU NOT AFR and UKB EUR cohorts are shown in blue and the AOU AFR and UKB AFR cohorts in red. In AoU, samples corresponding to gray dots are not used for fine-mapping and are primarily samples that are filtered due to excessive genetic relatedness but also include a smaller set of samples that either have self-identified race as Black/African American or genetically inferred ancestry as AFR (but not both). In UKB, the gray dots are also not used for fine-mapping and are samples that have self-identified ancestry belonging to other groups. **c. Global ancestry proportions of the different cohorts in AoU.** The African (n=44,384) and not African (n=174,828) cohorts correspond to those used for fine-mapping (see **Methods**). The European cohort (n=119,178) is shown for comparison, and consists of samples that have self-identified race as White and genetically inferred ancestry as EUR as reported by AoU. Although the non-African cohort is a superset of the European cohort, it still primarily consists of European ancestry. The table below the figure shows the average percentages corresponding to each ancestry component in each cohort (computation details in **Methods**). For each cohort, global ancestry percentages are averaged across all samples. (EUR=European; EAS=East Asian; SAS=South Asian; AFR=African; WAS=West Asian; AMR=Native American).

Supplementary Fig. 8

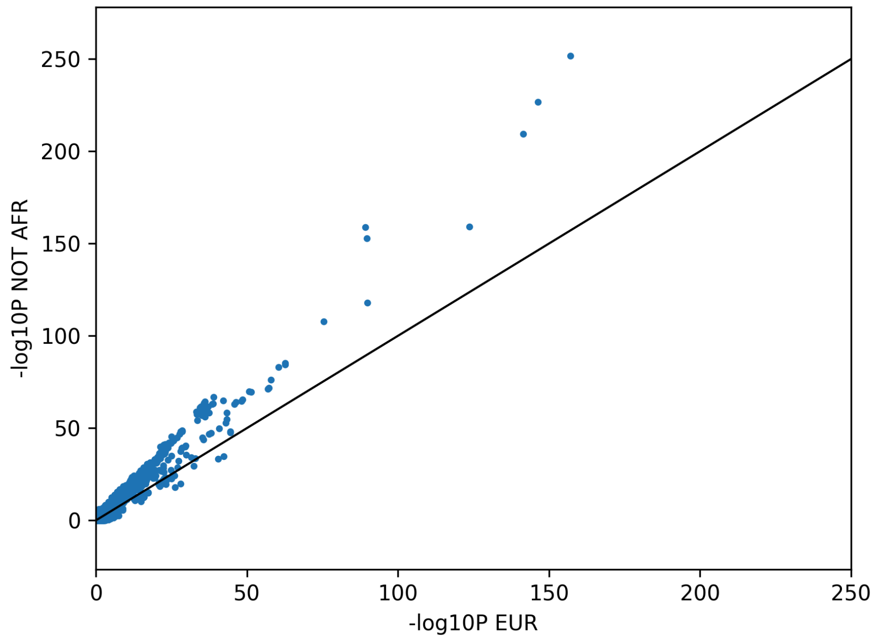


**A comparison of EUR and NOT AFR p-values for significant LDL GWAS variants.** 1503 variants are common across LDL cholesterol GWAS performed in AoU NOT AFR and AoU EUR (cohorts described in **Methods**) and achieve genome-wide significance in at least one cohort. For these variants, the -log10 p-value in NOT AFR is plotted against the -log10 p-value in EUR. The x=y line is shown for comparison.

Supplementary Fig. 9

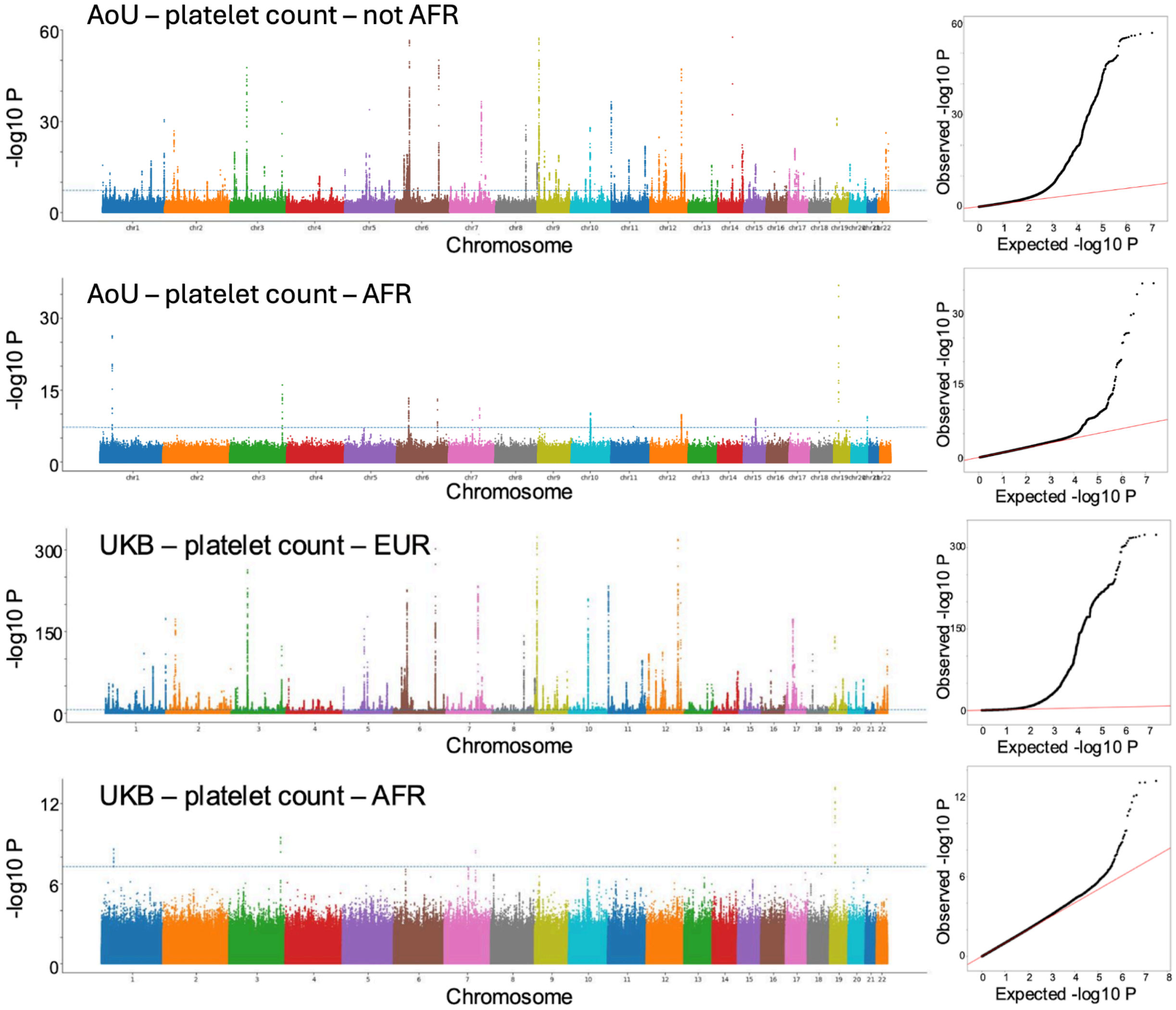


**GWAS for platelet count.** Left panels show Manhattan plots for GWASs for platelet count run separately in each of the 4 cohorts (AoU NOT AFR, AoU AFR, UKB EUR, UKB AFR). Right panels show quantile-quantile plots visualizing the distribution of p-values from each GWAS.

Supplementary Fig. 10

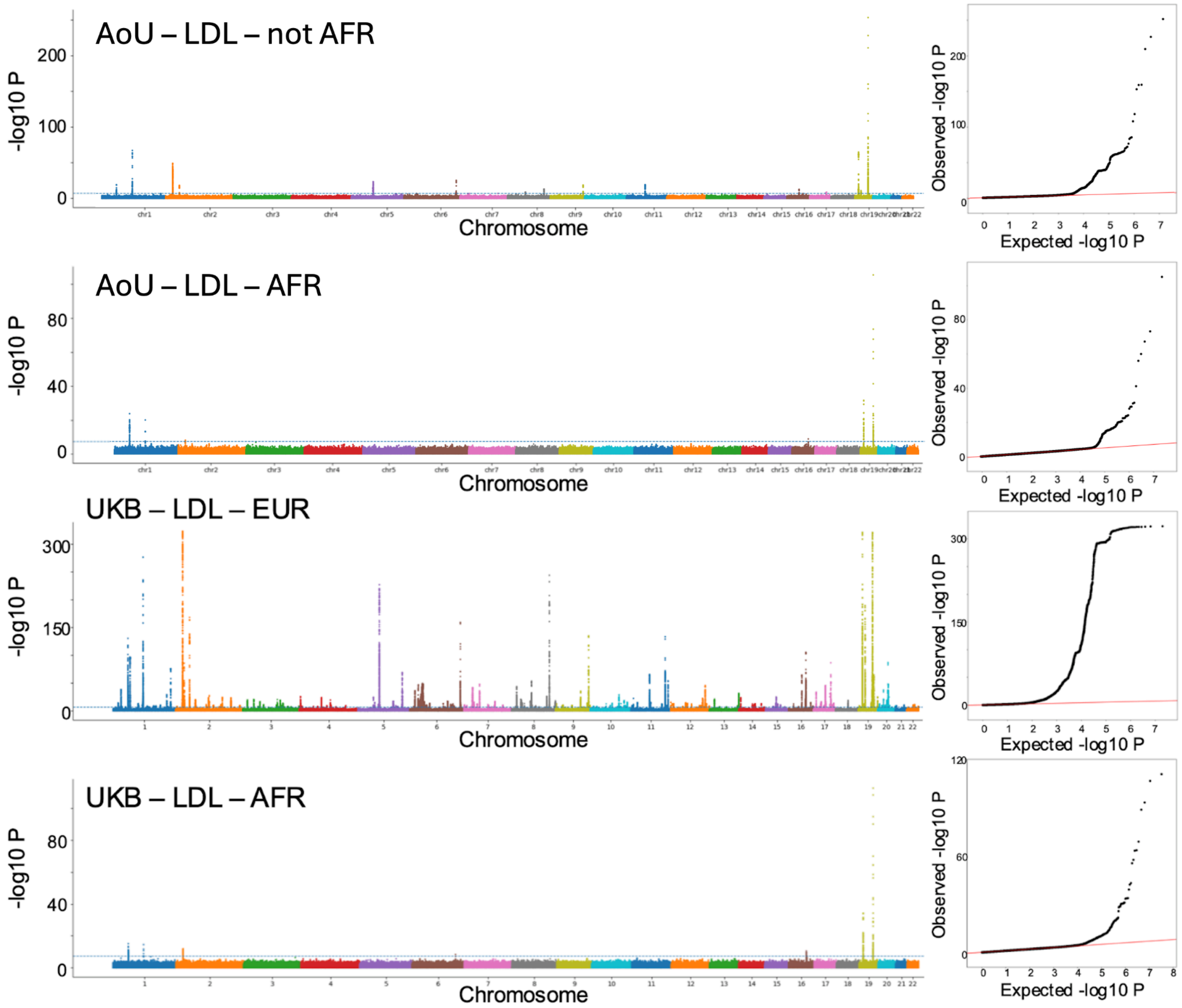


**GWAS for LDL cholesterol.** Left panels show Manhattan plots for GWASs for LDL cholesterol run separately in each of the 4 cohorts (AoU NOT AFR, AoU AFR, UKB EUR, UKB AFR). Right panels show quantile-quantile plots visualizing the distribution of p-values from each GWAS.

### Supplementary Fig. 11

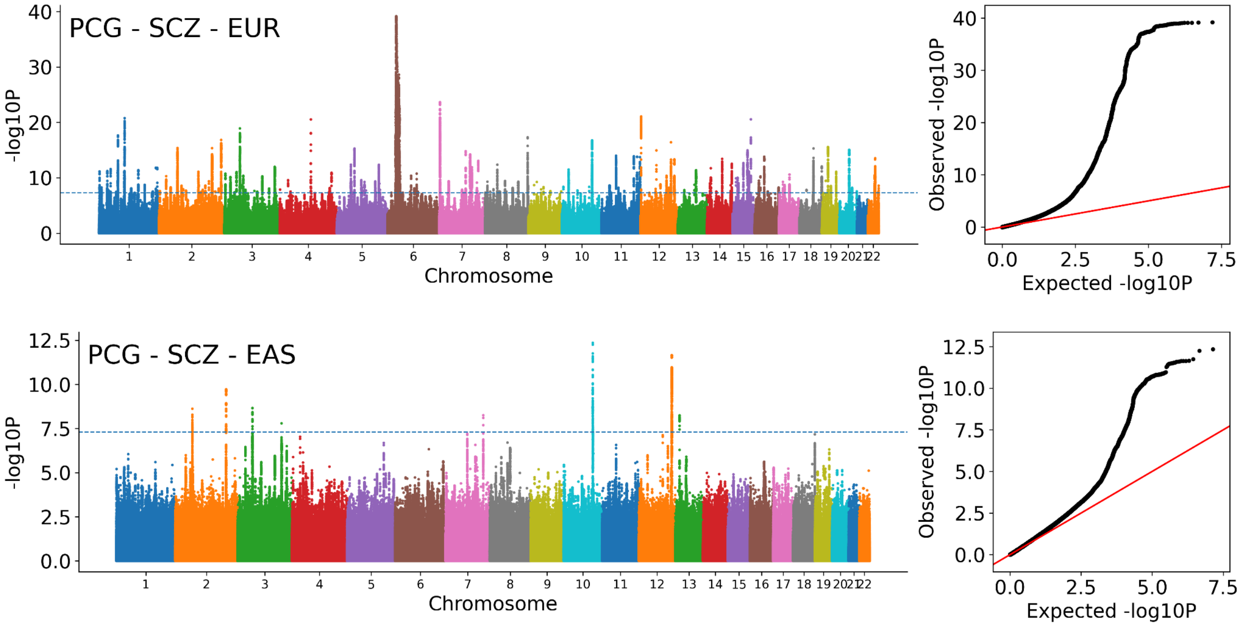


**GWAS for schizophrenia.** Left panels show Manhattan plots for GWASs from PGC for schizophrenia in EUR and EAS cohorts. Right panels show quantile-quantile plots visualizing the distribution of p-values from each GWAS.

### Supplementary Fig. 12

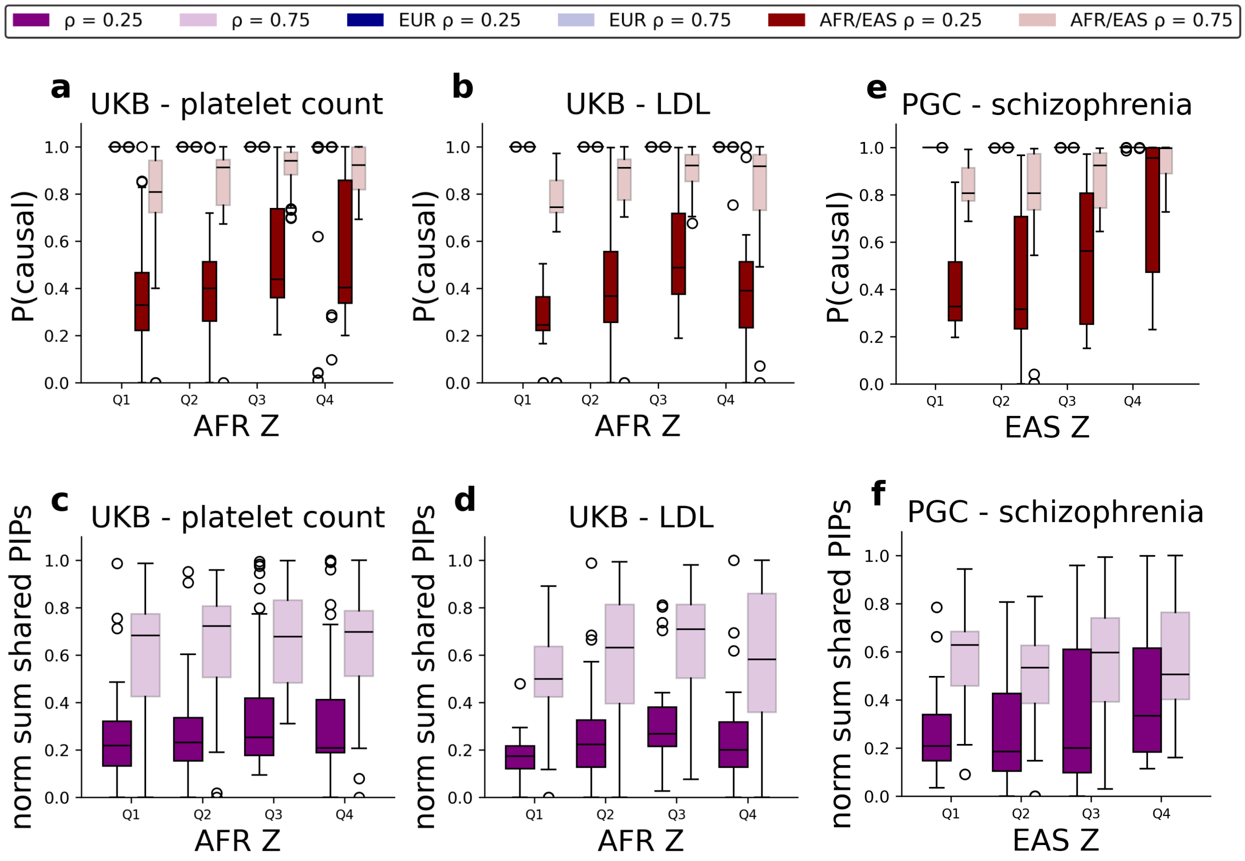


**Evidence of extensive signal sharing across populations in UKB (platelet count and LDL) and PGC SCZ. a-b. Probability of at least one causal variant at each trait-region in each population in UKB.** These are the same as **Supplementary Fig. 13a-b** but for UKB with UKB EUR in blue and UKB AFR in red. **c-d. Normalized sum of shared PIPs at each trait region.** These are the same as **Supplementary Fig. 13c-d** but for UKB. **e. Probability of at least one causal variant at each trait-region PGC.** These are the same as **Supplementary Fig. 13a-b** but for PGC with PGC EUR in blue and PGC EAS in red. **f. Normalized sum of shared PIPs at each trait region.** These are the same as **Supplementary Fig. 13c-d** but for PGC. Boxplot elements are as defined as in **Fig. 3**.

Supplementary Fig. 13

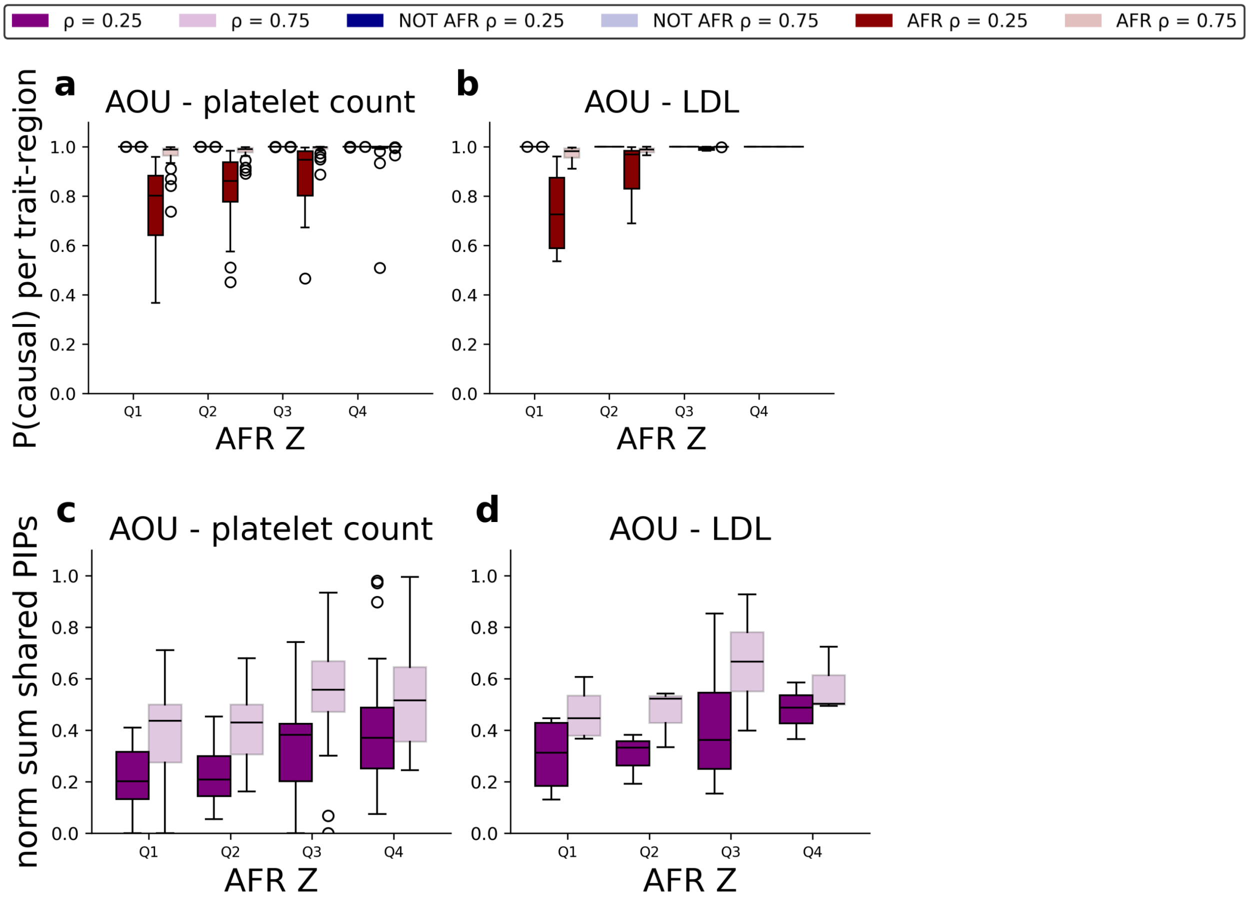


**Evidence of extensive signal sharing across African and non-African cohorts in AoU. a-b. Probability of at least one causal variant at each trait-region in each cohort.** Boxplots show the distribution of P(causal), computed as 1-P(no causal variant), for each cohort. The x-axis stratifies results by quartiles for the strongest absolute AFR z-score for a variant in the trait-region. Results are shown separately for each cohort (blue=NOT AFR, red=AFR) and for each value of the sharing parameter tested (dark=0.25, light=0.75). Results for **(a)** platelet count and **(b)** LDL. **c-d. Normalized sum of shared PIPs at each trait region.** Boxplots show the distributions of the normalized sum of shared PIPs, computed as the sum of shared PIPs over all variants returned by PIPSORT (those with at least one per-population PIP > 0.05) divided by the number of causal variants considered at each trait-region. The x-axis stratifies results by AFR association statistics as in **(a-b)**. For each quartile, the left boxplot with darker shading corresponds to ρ=0.25 and the right to ρ=0.75. **(c)** shows the results for platelet count and **(d)** for LDL. Boxplot elements are as defined as in **Fig. 3**. Similar plots without stratification are shown in **Fig. 3b-c**.

Supplementary Fig. 14

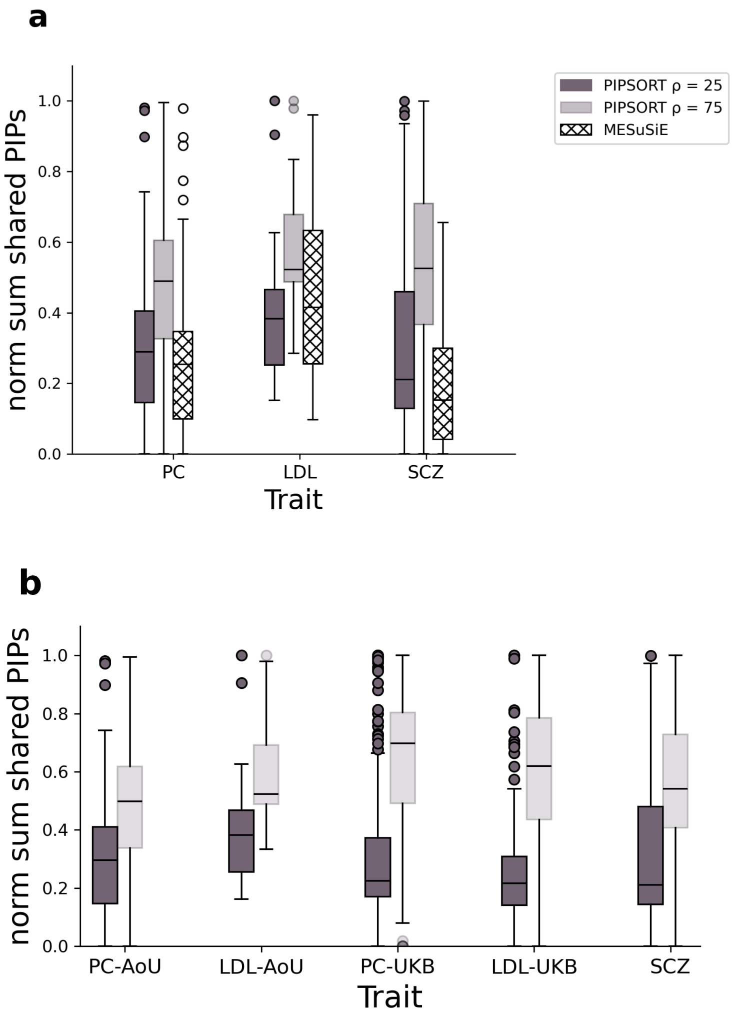


**Evidence of shared causal signals across AoU, UKB, and SCZ. a. Normalized sum of shared PIPs at each trait region for PIPSORT and MESuSiE in AoU and SCZ.** We summed the shared PIPs (probability that a variant is a shared causal signal) across all variants returned by PIPSORT and MESuSiE and normalized by the maximum number of causal variants considered at each trait-region. This metric can be interpreted as the average probability that each independent signal is shared. Boxplots show the distributions of the normalized sum of shared PIPs, computed as the sum of shared PIPs over all variants with at least one per-population PIP ≥ 0.1 for PIPSORT and with a shared PIP ≥ 0.1 for MESuSiE divided by the number of causal variants considered at each trait-region. For PIPSORT, the boxplots with darker shading correspond to ρ=0.25 and lighter shading to ρ=0.75. Boxplot elements are the same as in **Fig 3b. b. Normalized sum of shared PIPs at each trait region for PIPSORT in AoU, UKB, and SCZ.** This plot is the same as **(a)** but includes the data for UKB. As MESuSiE was not run in UKB, this plot contains data for PIPSORT only. No comparable metric is output by SuSiEx.

### Supplementary Fig. 15

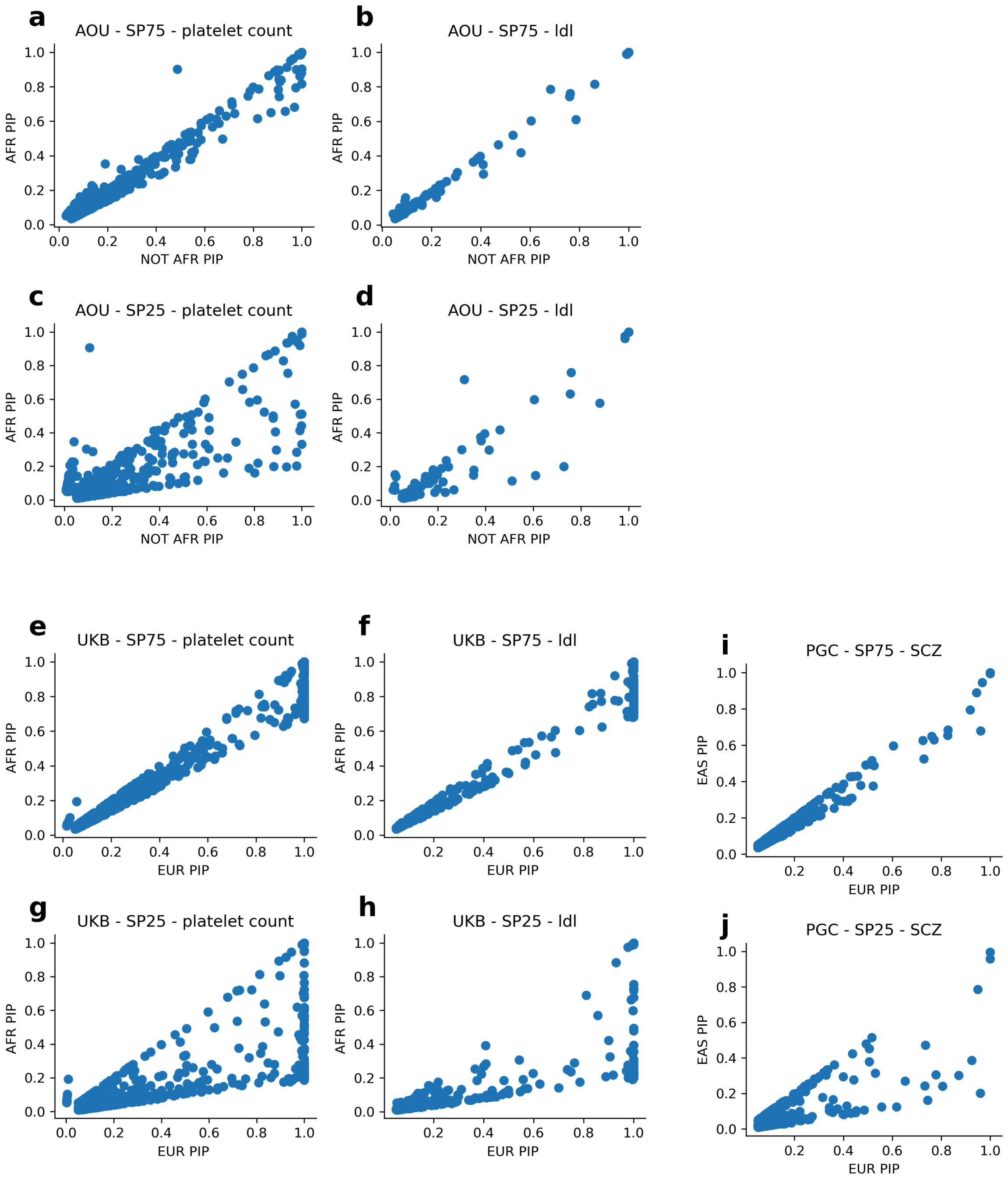


**Comparison of EUR/non-AFR PIPs (x-axis) vs. AFR/EAS PIPs (y-axis) across both population cohorts for all traits in AoU, UKB, and PGC. a-d** show results for AoU, **e-h** for UKB, and **i-j** for PGC. Within each dataset the top row of plots show results for ρ=0.75 and bottom row of plots show ρ=0.25. Plots include all variants for which PIPSORT returned a PIP of at least 0.05 in at least one population for each trait.

Supplementary Fig. 16

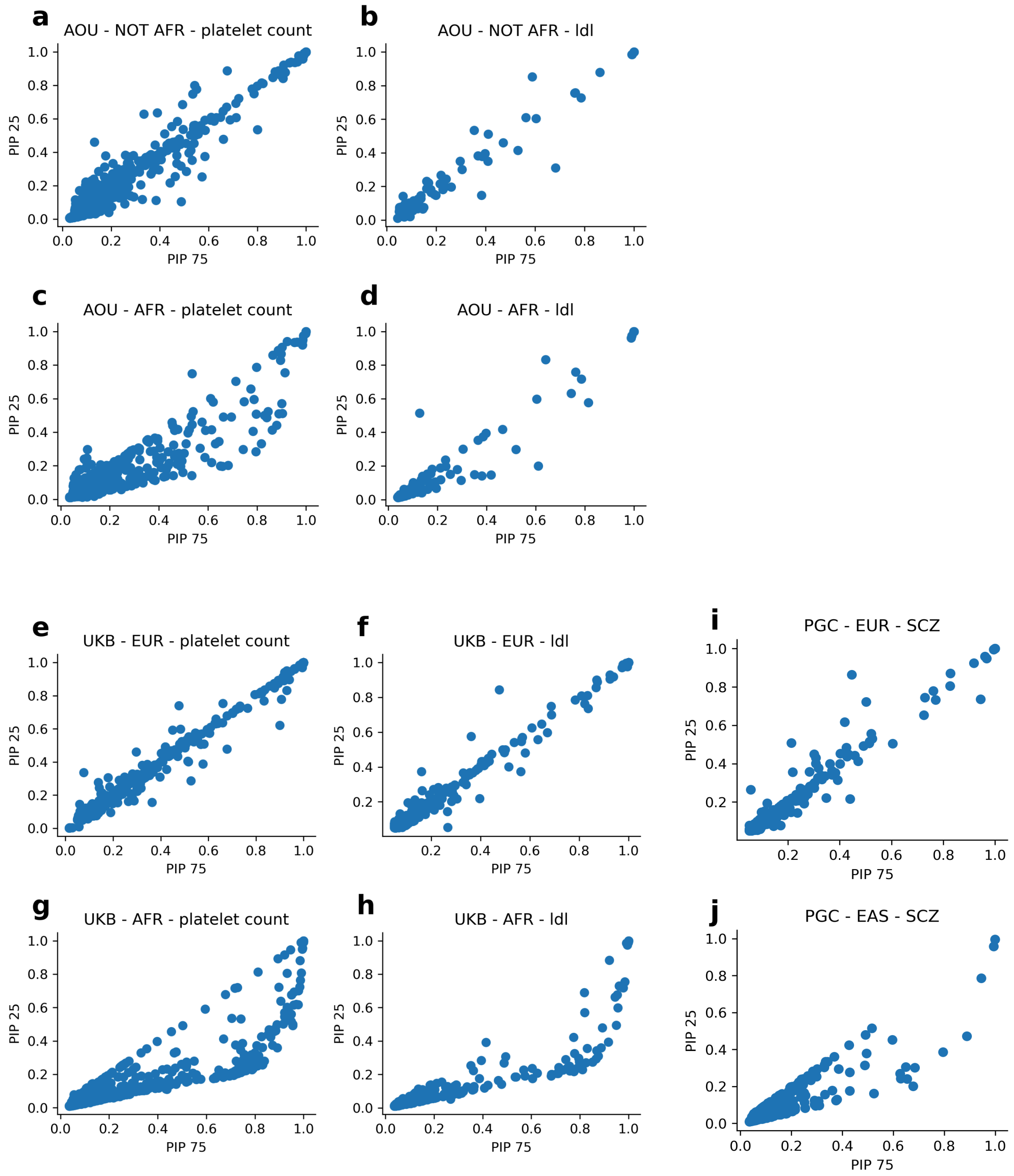


**Comparison of ancestry-specific variant-level PIPs with ρ=0.75 (x-axis) vs. ρ=0.25 (y-axis) across both population cohorts for all traits in AoU and UKB. a-d** show results for AoU and **e-h** show results for UKB. Plots include all variants for which PIPSORT returned a PIP of at least 0.05 in at least one population for each trait.

### Supplementary Fig. 17

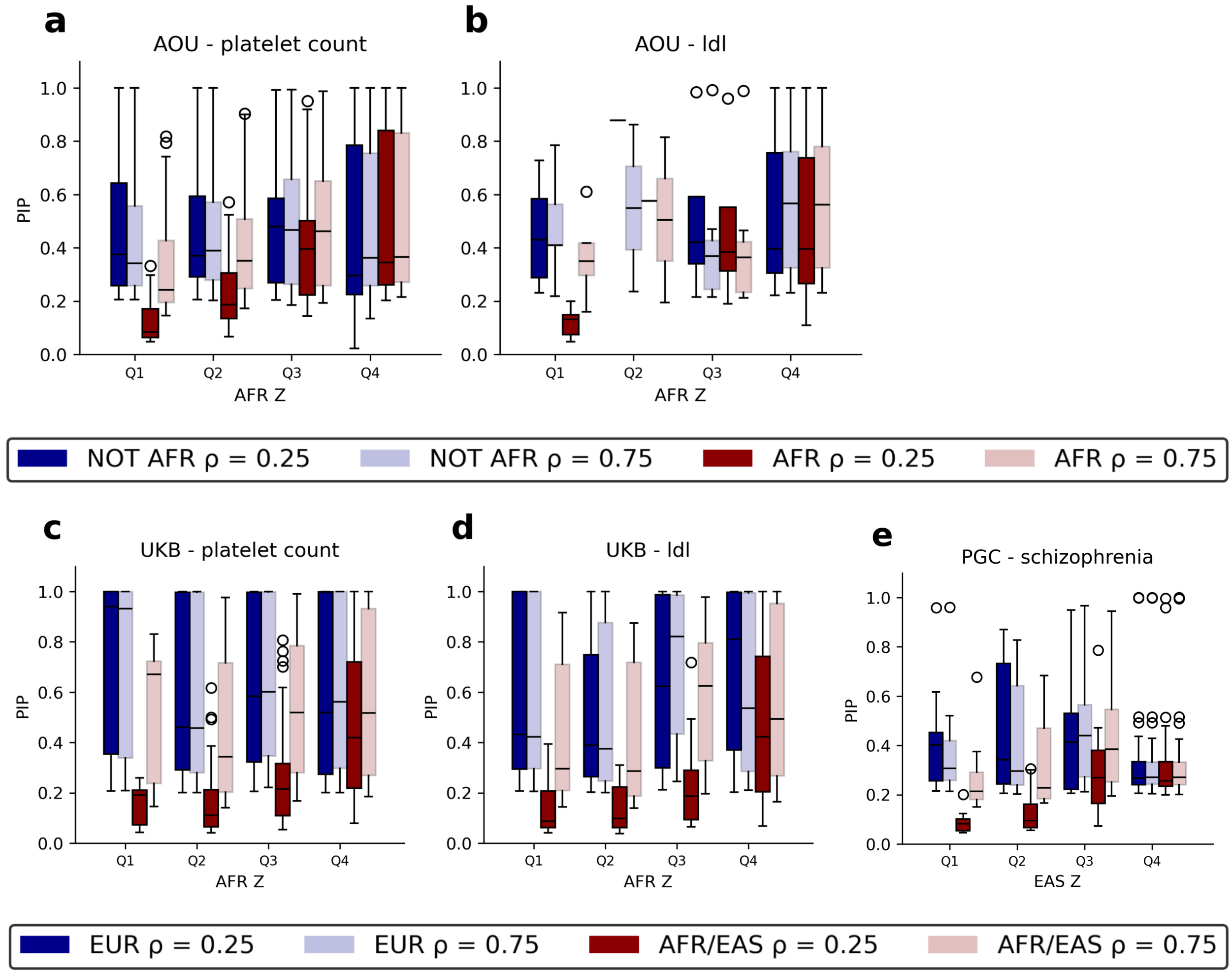


**PIP in each ancestry cohort across different values of ρ stratified by the significance of AFR association statistics in AoU, UKB, and PGC.** The y-axis shows ancestry-specific PIPs for all variants for which PIPSORT returned a PIP of at least 0.05 in at least one population for each trait. Blue=EUR; red=EUR/not AFR; dark=sharing parameter 0.25; light=sharing parameter 0.75. **a-b** show results for AoU, **c-d** for UKB and **e** for PGC. The x-axis stratifies results by quartiles for the strongest absolute AFR z-score for a variant in the trait-region. In all plots horizontal lines show median values, boxes span from the 25th percentile (Q1) to the 75th percentile (Q3), and whiskers extend to Q1–1.5*IQR (bottom) and Q3+1.5*IQR (top), where IQR gives the interquartile range (Q3-Q1). Outlier values are shown as individual data points.

### Supplementary Fig. 18

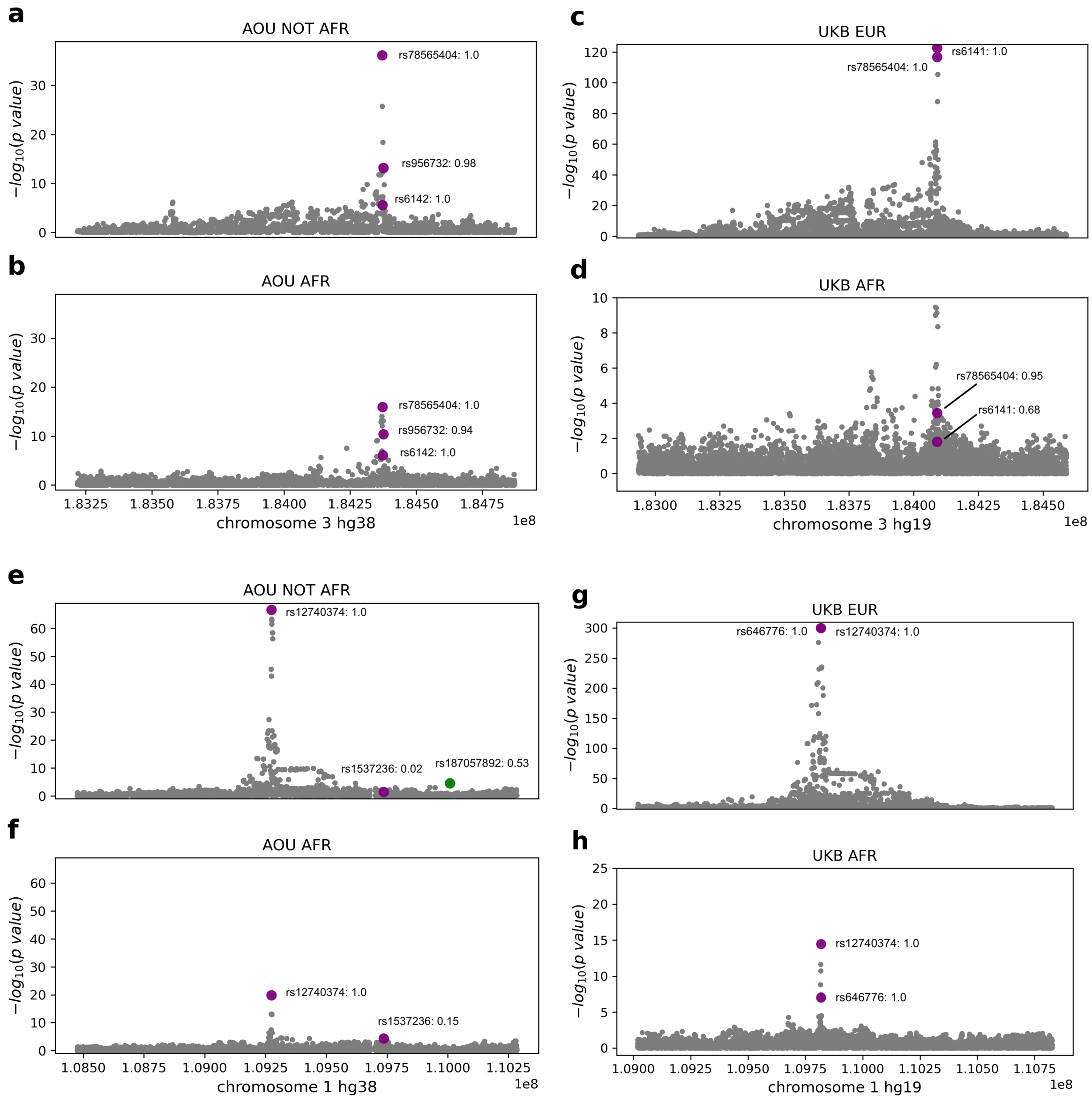


**Additional examples of high confidence shared signals common to both AoU and UKB.** Purple dots are variants that were fine-mapped in both cohorts. Green dots are rare in AFR and fine-mapped only in NOT AFR/EUR. **a-d. A Chromosome 3 trait-region for platelet count.** Manhattan plots are shown for AoU NOT AFR **(a)**, AoU AFR **(b)**, UKB EUR **(c)**, and UKB AFR **(d)**. The variant rs78565404, a 3 prime UTR variant in *THPO*, is commonly fine-mapped with PIP>=0.95 (with $\rho$=0.25) across all cohorts. **e-h. A Chromosome 1 trait-region for LDL.** Manhattan plots are shown for AoU NOT AFR **(e)**, AOU AFR **(f)**, UKB EUR **(g)**, and UKB AFR **(h)**. For simplicity, we annotate variants for which PIPSORT computed a PIP>0.1. The variant rs12740374, a 3 prime UTR variant in *CELSR2*, is commonly fine-mapped with PIP=1.0 (with $\rho$ =0.25) across all cohorts.

#### Supplementary Fig. 19

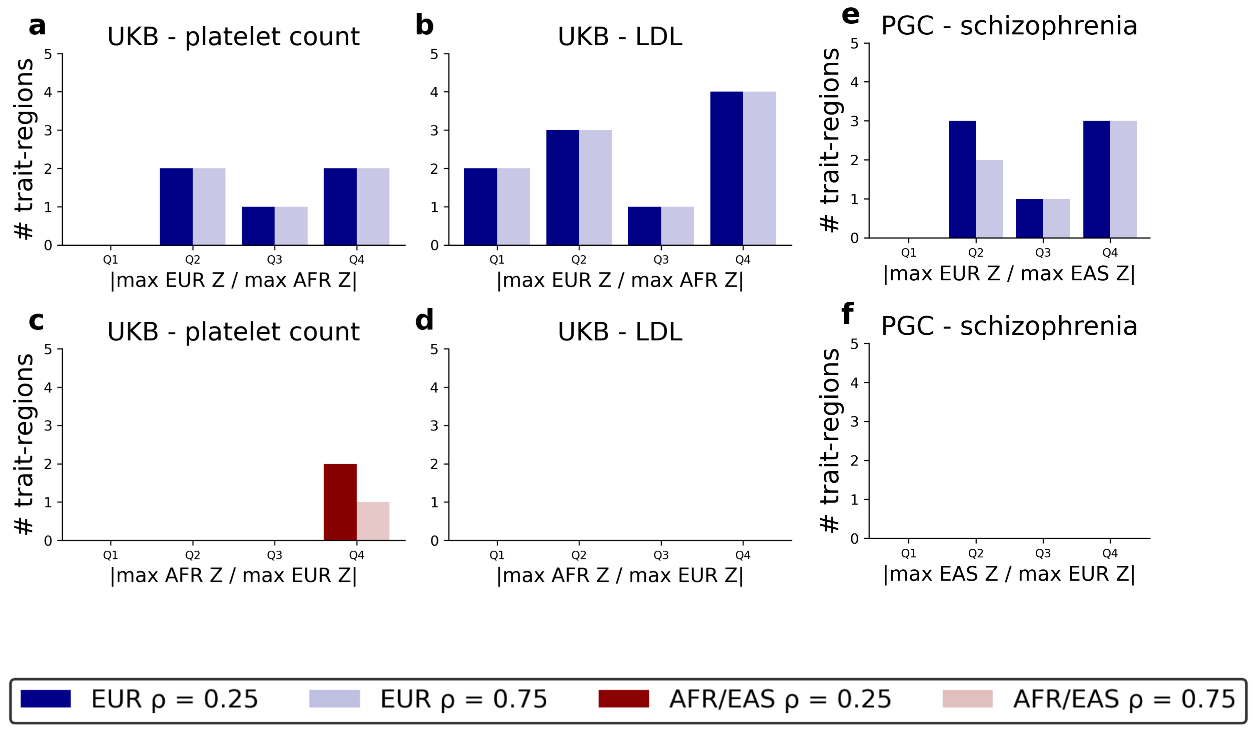


**Ancestry specific causal variants identified in UKB and PGC. a-b. The number of trait-regions in UKB with AS-Vs in the EUR cohort for different values of ρ stratified by the relative significance of EUR to AFR association statistics.** The height of the bar (y-axis) corresponds to the number of trait-regions with EUR AS-Vs. The x-axis stratifies the bars by quartiles for the absolute value of the ratio of the strongest EUR Z-score to the strongest AFR Z-score for a variant in the trait-region. For each quartile, the left bar with darker shading corresponds to ρ=0.25 and the right bar with lighter shading to ρ=0.75 **(a)** shows the results for platelet count and **(b)** for LDL. **c-d. The number of trait-regions in UKB with AS-Vs in the AFR cohort for different values of ρ stratified by the relative significance of AFR to EUR association statistics.** These plots are the same as those in **(a-c)** but counting the trait-regions with AFR AS-Vs vs. the relative significance of AFR to EUR association statistics. **(c)** shows the results for platelet count, and **(d)** for LDL. **e-f. The number of trait-regions in PGC with AS-Vs for schizophrenia.** These plots are the same as those in **(a-d)** but **(e)** counts the schizophrenia trait-regions with EUR AS-Vs vs. the relative significance of EUR to EAS association statistics and **(f)** shows that we did not detect any EAS AS-Vs in PGC. Similar plots for AoU are shown in **Fig. 4**.

### Supplementary Fig. 20

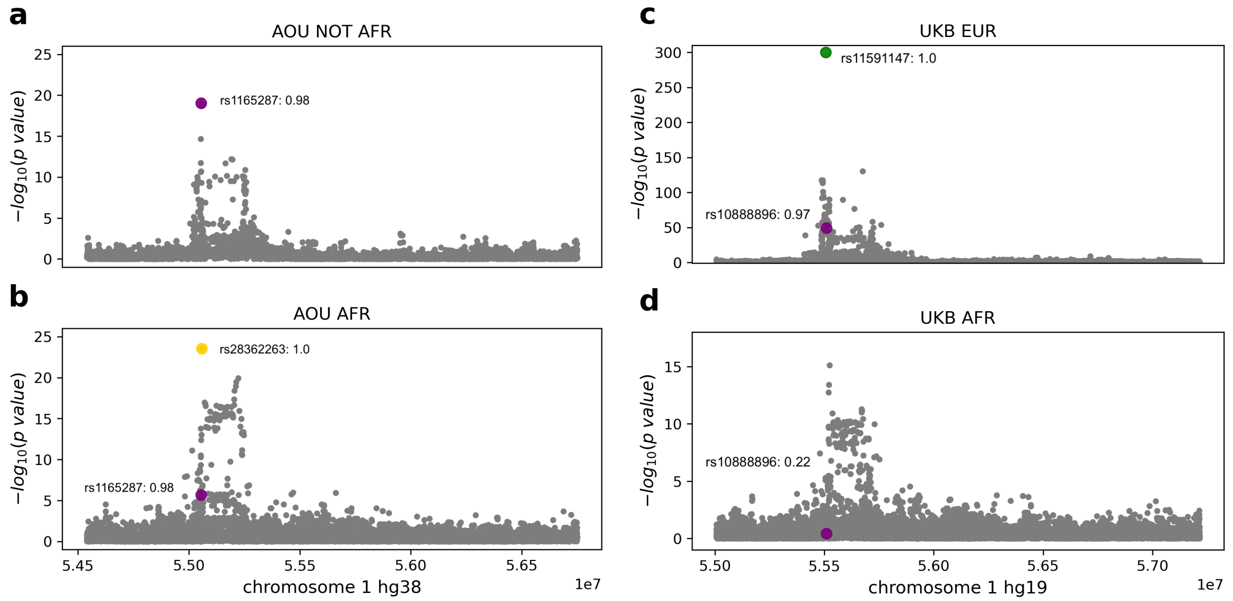


**Ancestry-specific variants in *PCSK9*.** Manhattan plots are shown for an LDL associated trait-region overlapping *PCSK9* for AoU NOT AFR **(a)**, AoU AFR **(b)**, UKB EUR **(c)**, and UKB AFR **(d)**. Across AoU and UKB, rs10888896 (purple), an intronic variant near PCSK9, is fine-mapped in all cohorts. In AoU, PIPSORT identifies an AFR AS-V, rs28362263 (yellow), a missense variant in *PCSK9,* which is common in AFR but rare in all other AoU ancestry cohorts, with a PIP of 1 in AFR. In UKB, rs28362263 is included as input to PIPSORT in UKB AFR but is assigned a near-zero PIP. The variant rs10888896 is assigned a strong PIP in UKB EUR but not in UKB AFR. In addition, PIPSORT identifies rs11591147 (green), another missense variant in *PCSK9*, as a EUR AS-V in UKB. The location of this variant on the Manhattan plot in **(c)** is approximate as the p-value was below the precision of the association testing framework. This variant is not fine-mapped in AoU as it is tri-allelic in AoU and was filtered by our pipeline, which considered only bi-allelic variants for analysis.

### Supplementary Fig. 21

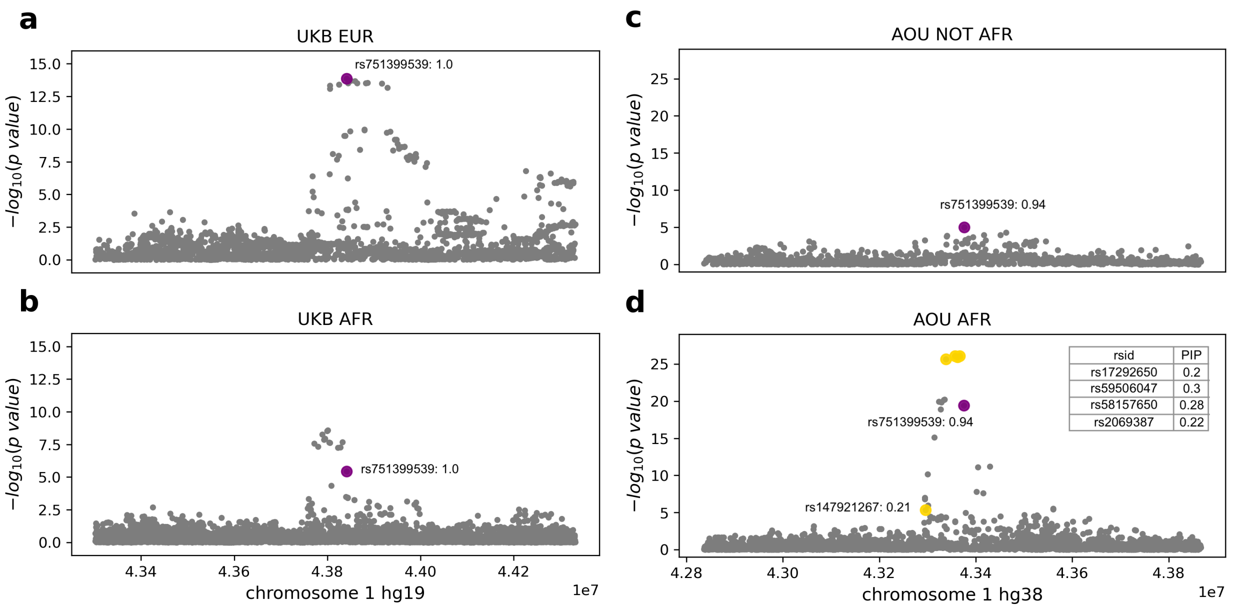


**Variant associations at the *MPL* trait-region for platelet count.** Manhattan plots are shown for a platelet count associated trait-region overlapping *MPL* for UKB EUR **(a)**, UKB AFR **(b)**, AoU NOT AFR **(c)**, and AoU AFR **(d)**. Panels **c-d** are duplicated from **Fig. 4g-h** for ease of comparison. We annotate variants with PIP≥0.1. One variant, rs75139539, is fine-mapped with a strong PIP of 1 in both the UKB EUR cohort **(a)** and UKB AFR cohort **(b)**. This variant is also fine-mapped in AoU with strong PIPs in both the AOU NOT AFR and AFR cohorts. All 4 AS-V variants (yellow) fine-mapped in AoU AFR correspond to points in the cluster of gray dots above the purple dot in **(b)**, but are fine-mapped with near-zero PIPs in UKB AFR.

### Supplementary Fig. 22

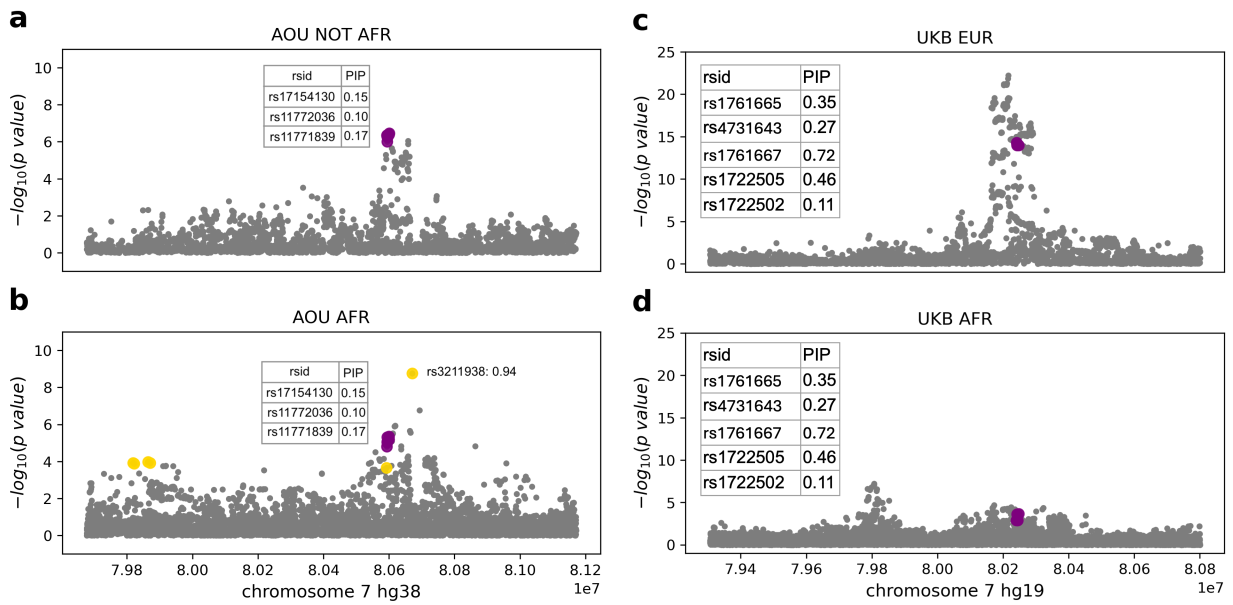


**An additional example high confidence AS-V.** Manhattan plots are shown for a platelet count associated trait-region overlapping *CD36* for AoU NOT AFR **(a)**, AoU AFR **(b)**, UKB EUR **(c)**, and UKB AFR **(d)**. For simplicity, we annotate variants with PIP>0.1 although additional variants output by PIPSORT are illustrated in the plots. Purple variants are ones that were fine-mapped in both cohorts and yellow are rare in NOT AFR/EUR and fine-mapped only in AFR. Across all 4 Manhattan plots, the overlapping purple dots are annotated in the inset tables. We do not observe any overlap in the fine-mapped variants (output by PIPSORT with PIP≥0.5) across AoU and UKB for this trait-region. In AoU, the variant rs3211938, a stop-gained variant in *CD36*, is assigned the strongest PIP of 0.94 in AoU AFR. It is not included as input to PIPSORT in AoU NOT AFR as it is very rare (MAF<0.001) in that cohort. It is not included as input to PIPSORT in UKB as its p-value did not pass our significance threshold (see **Methods**).

### Supplementary Fig. 23

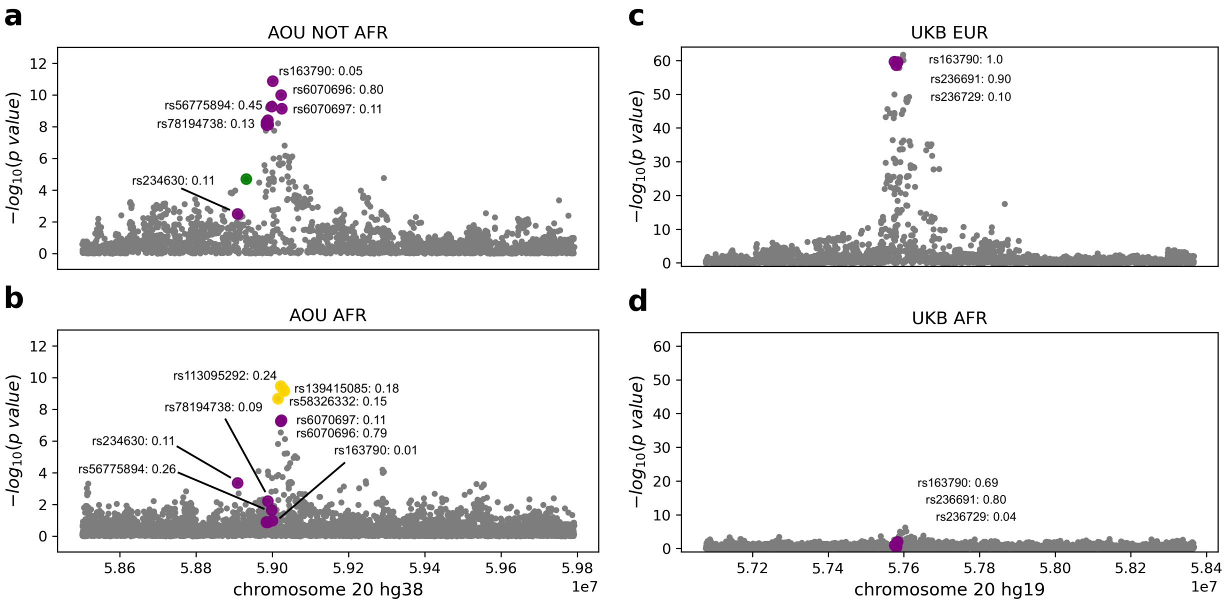


**An example high confidence AS-V for the AFR AoU cohort.** Manhattan plots are shown for a platelet count associated trait-region in Chromosome 20 overlapping *TUBB1* and *ATP5F1E* for AoU NOT AFR **(a)**, AoU AFR **(b)**, UKB EUR **(c)**, and UKB AFR **(d)**. For simplicity, we annotate variants with PIP>=0.1 (with the exception of rs163790) although additional variants output by PIPSORT are illustrated in the plots. Purple variants are ones that were fine-mapped in both cohorts, yellow are rare in NOT AFR and fine-mapped only in AFR, green are rare in AFR and fine-mapped only in NOT AFR. In AoU, three variants (yellow) are candidate AFR AS-Vs, two of which are intronic variants in *TUBB1* and *ATP5F1E*, and one of which (rs139415085) is a 3 prime UTR variant in *ATP5F1E*. The majority of the annotated variants are intronic variants. Variant rs151352 is also a 3 prime UTR variant in *ATP5F1E*. Variant rs6070697 is a missense variant in *TUBB1*. Commonly fine-mapped in AoU and UKB, although with much lower PIPs in AoU, variant rs163790 is an intronic variant in *CTSZ*. Variant rs139415085 is not included in the UKB GWAS. Variants rs113095292 and rs58326332 are in the UKB AFR GWAS with relatively significant p-values (2.1e-5 and 7.3e-6 respectively) given the cohort size, but receive near-zero PIPs.

### Supplementary Fig. 24

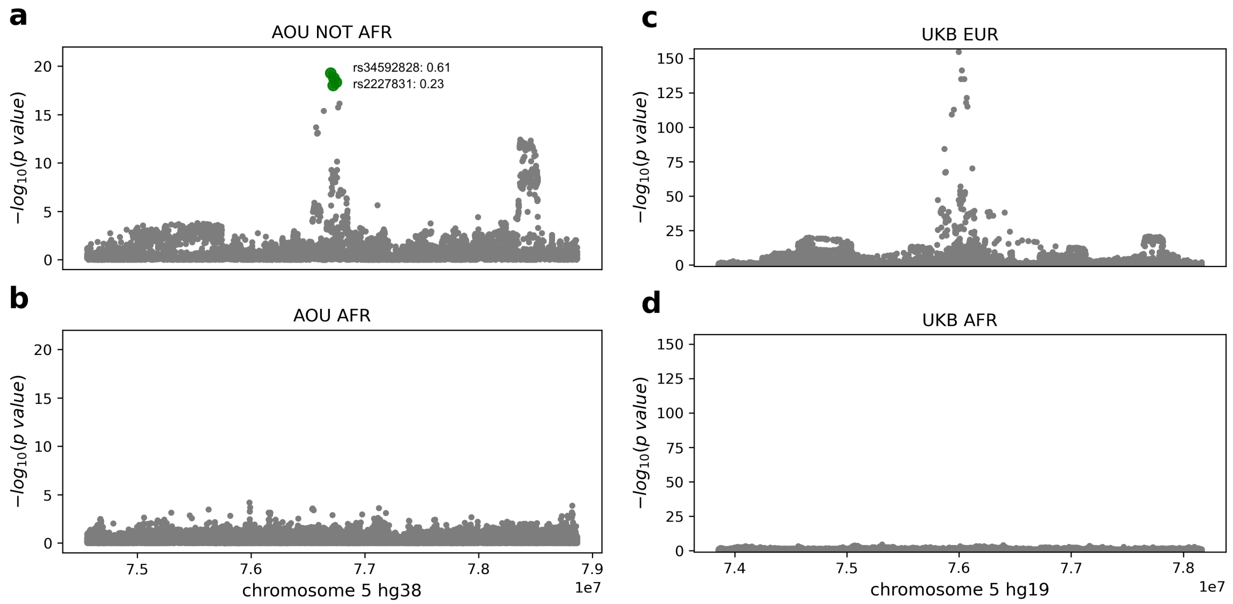


**An example high confidence AS-V for the NOT AFR AoU cohort.** Manhattan plots are shown for a platelet count associated trait-region overlapping *IQGAP2* for AoU NOT AFR **(a)**, AoU AFR **(b)**, UKB EUR **(c)**, and UKB AFR **(d)**. For simplicity, we annotate variants with PIP≥0.1 although additional variants output by PIPSORT are illustrated in the plots. Green variants are ones that are rare in AFR and fine-mapped only in NOT AFR. In AoU, the variant rs34592828, a missense variant in *IQGAP2*, is assigned the strongest PIP of 0.61 in AoU NOT AFR. It is very rare, and therefore not fine-mapped, in AoU AFR. PIPSORT did not run in the UKB cohort as there were too many variants in this region (see **Methods**). We still include the UKB Manhattan plots for comparison as well as to note that the variant rs34592828 is the most significant variant in **(c)** and corresponds to the gray dot at the very top of the UKB EUR Manhattan plot.

### Supplementary Fig. 25

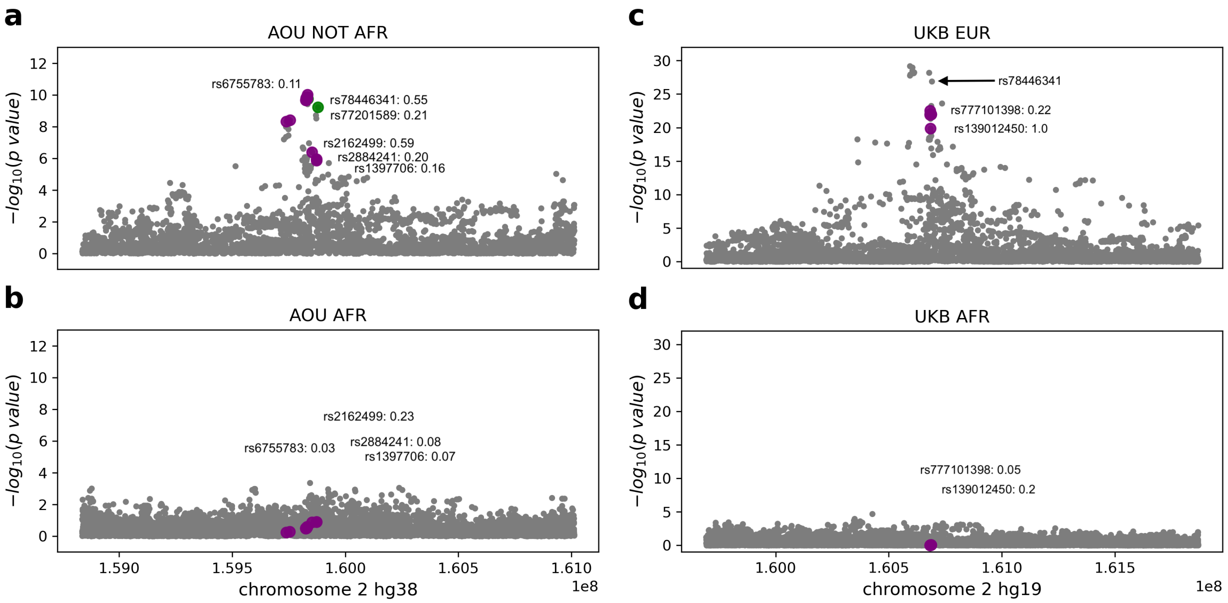


**An example high confidence AS-V for the NOT AFR AoU cohort.** Manhattan plots are shown for a platelet count associated trait-region in Chromosome 2 overlapping *LY75* for AoU NOT AFR **(a)**, AoU AFR **(b)**, UKB EUR **(c)**, and UKB AFR **(d)**. For simplicity, we annotate variants with PIP>=0.1 although additional variants output by PIPSORT are illustrated in the plots. Purple variants are ones that were fine-mapped in both cohorts and green are rare in AFR and fine-mapped only in NOT AFR. In AoU, the variant rs78446341, a missense variant in *LY75*, is assigned a strong PIP of 0.55 in AoU NOT AFR. It is very rare, and therefore not fine-mapped, in AoU AFR. This variant was passed as input to PIPSORT in UKB but was assigned a near-zero PIP. We highlight it in **(c)** and note that it has a stronger p-value in UKB EUR than the variants that were fine-mapped with more significant PIPs. The majority of the other variants illustrated are intronic variants in *LY75*. Variant rs1397706 fine-mapped in AoU is also a missense variant for *LY75*.

### Supplementary Fig. 26

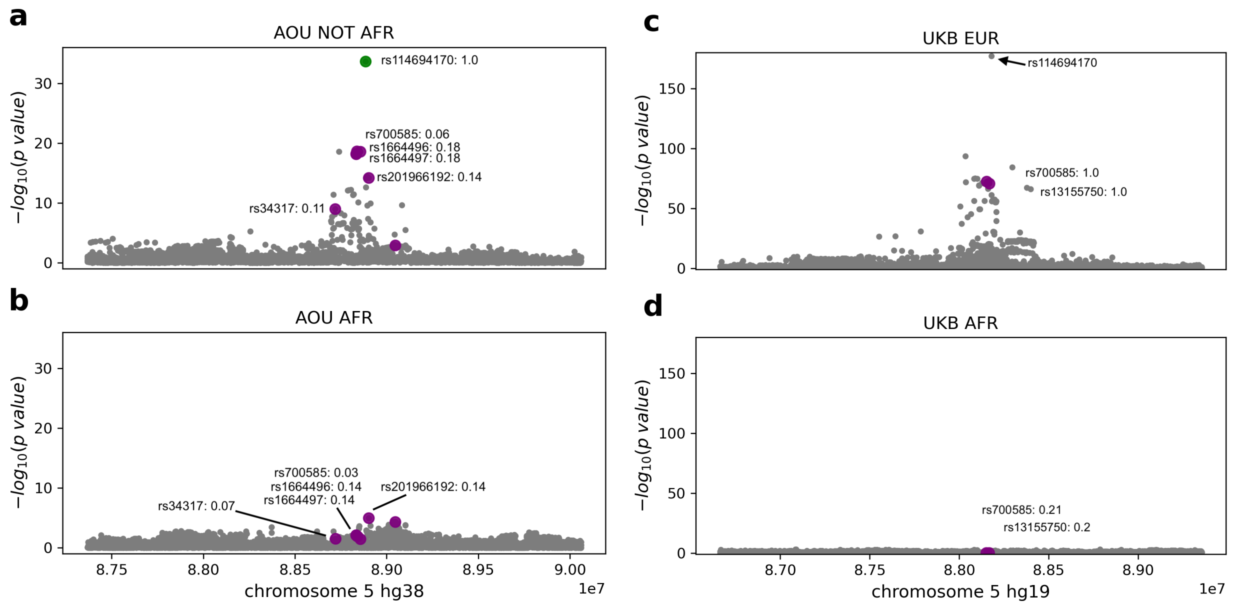


**An example high confidence AS-V for the NOT AFR AoU cohort.** Manhattan plots are shown for a platelet count associated trait-region in Chromosome 5 overlapping *MEF2C* for AoU NOT AFR **(a)**, AoU AFR **(b)**, UKB EUR **(c)**, and UKB AFR **(d)**. For simplicity, we annotate variants with PIP≥0.1 although additional variants output by PIPSORT are illustrated in the plots. Purple variants are ones that were fine-mapped in both cohorts and green are rare in AFR and fine-mapped only in NOT AFR. In AoU, the variant rs114694170, a non-coding exon variant for *MEF2C*, is assigned the strongest PIP of 1.0 in AoU NOT AFR. It is very rare, and therefore not fine-mapped, in AoU AFR. This variant was passed as input to PIPSORT in UKB but was assigned a near-zero PIP. We highlight it in **(c)** and note that it has a stronger p-value in UKB EUR than the variants that were fine-mapped with more significant PIPs. All other variants annotated in the figures are intronic variants in *MEF2C*.

### Supplementary Fig. 27

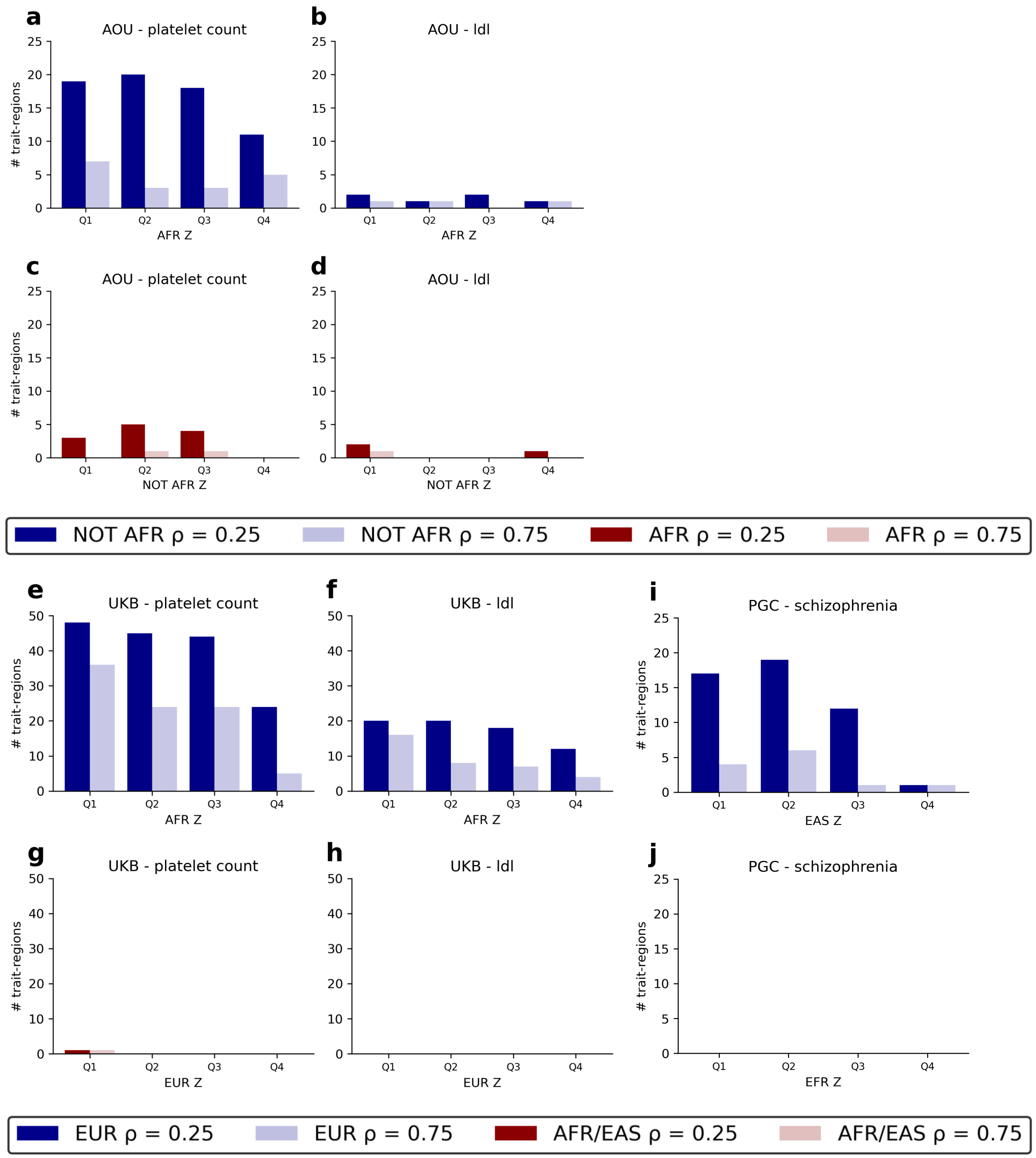


**Ancestry specific causal effects identified in AOU, UKB, and PGC.** All the plots shown here count the number of trait-regions in one dataset with AS-Es in one cohort for different values of ρ stratified by the significance of association statistics in the other cohort. The height of the bar (y-axis) corresponds to the number of trait-regions with AS-Es in one cohort while the x-axis stratifies the bars by quartiles for the strongest Z-score for a variant in the trait-region in the other cohort. For each quartile, the left bar with darker shading corresponds to ρ=0.25 and the right bar with lighter shading to ρ=0.75. Results are provided for AOU in **(a-d)**, UKB in **(e-h)**, and PGC in **(i-j)**. **(a-b)** show AOU NOT AFR AS-Es stratified by the significance of AFR association statistics for platelet count **(a)** and LDL **(b)**. Likewise, **(c-d)** show AOU AFR AS-Es stratified by the significance of NOT AFR association statistics for both traits. **(e-f)** show UKB EUR AS-Es stratified by the significance of AFR association statistics. **(g-h)** show UKB AFR AS-Es stratified by the significance of EUR association statistics. Lastly, **(i)** shows PGC EUR AS-Es stratified by the significance of EAS association statistics for schizophrenia and **(j)** shows that we did not detect any EAS AS-Es in PGC for schizophrenia.

#### Supplementary Fig. 28

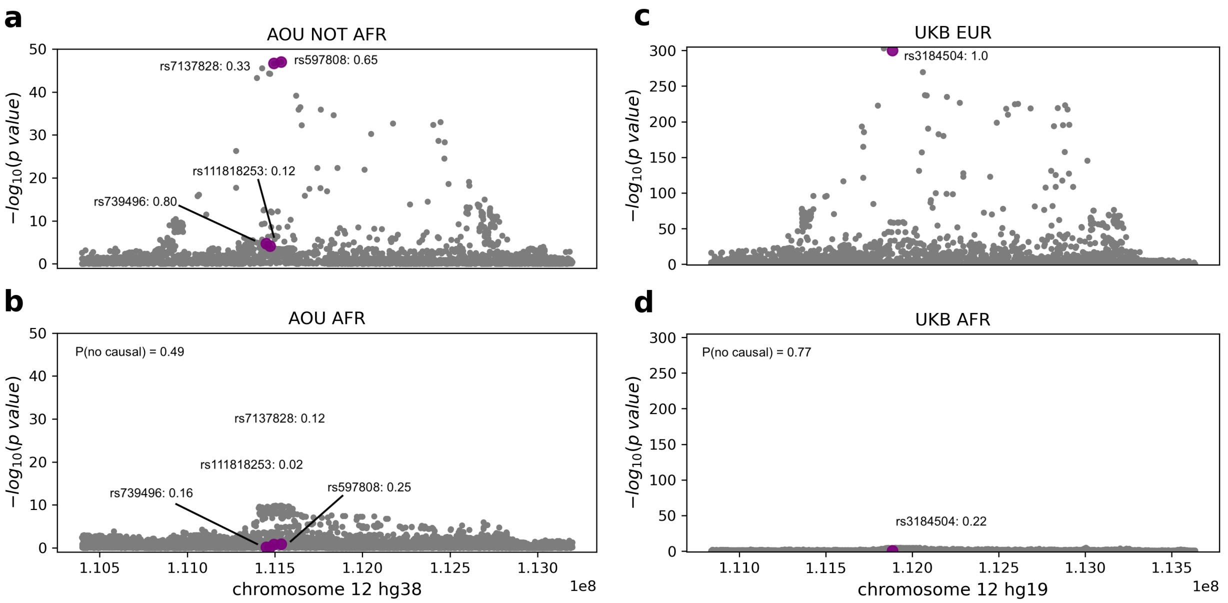


**An example high confidence NOT AFR/EUR AS-E in AoU and UKB.** Manhattan plots are shown for a platelet count associated Chromosome 12 trait-region overlapping *SH2B3* and *ATXN2* for AoU NOT AFR **(a)**, AoU AFR **(b)**, UKB EUR **(c)**, and UKB AFR **(d)**. Purple variants are ones that were fine-mapped in both cohorts. We use ρ=0.25. Variant rs739496 is assigned the highest PIP of 0.80 in AoU NOT AFR and is a 3 prime UTR variant in *SH2B3*. Variant rs3184504, a missense variant for *SH2B3*, is assigned the highest PIP of 1.0 in UKB EUR, but is filtered from our AoU analyses as it is tri-allelic. These variants are assigned much lower PIPs of 0.16 and 0.22 in AoU AFR and UKB AFR respectively. Further, the probability that the African cohort does not have any causal variants at this trait-region is 0.49 in AoU and 0.77 in UKB. The replication of this trend in AoU, and not just in UKB, increases our confidence that this region contains a true candidate NOT AFR/EUR AS-E as opposed to the difference in PIPs stemming from a lack of statistical power.

#### Supplementary Fig. 29

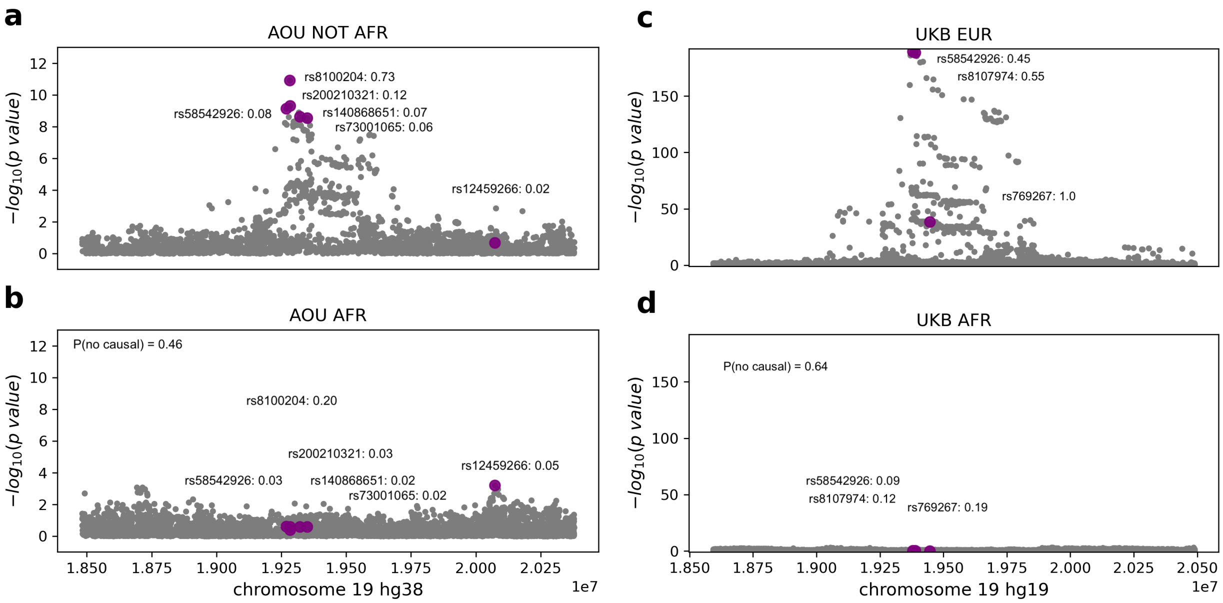


**An example high confidence NOT AFR/EUR AS-E in AoU and UKB.** Manhattan plots are shown for an LDL associated Chromosome 19 trait-region for AoU NOT AFR **(a)**, AoU AFR **(b)**, UKB EUR **(c)**, and UKB AFR **(d)**. Purple variants are ones that were fine-mapped in both cohorts. We use $\rho$=0.25. In AoU, variant rs8100204, an intronic variant for *SUGP1*, is assigned the highest PIP in this trait-region of 0.73 for AoU NOT AFR and a much lower PIP of 0.20 in AoU AFR. In UKB, we observed 3 variants assigned strong PIPs in UKB EUR and relatively lower PIPs in UKB AFR. Variant rs769267 is a synonymous variant in *MAU2*, rs8107974 is an intronic variant in *SUGP1*, and rs58542926 is a stop gain variant in *TM6SF2*. Variant rs58542926 is also fine-mapped in AoU but with much lower PIPs in both AoU cohorts. Further, the probability that the African cohort does not have any causal variants at this trait-region is 0.46 in AoU and 0.64 in UKB. The replication of this trend in AoU, and not just in UKB, increases our confidence that this region contains a true candidate NOT AFR/EUR AS-E as opposed to the difference in PIPs stemming from a lack of statistical power.

#### Supplementary Fig. 30

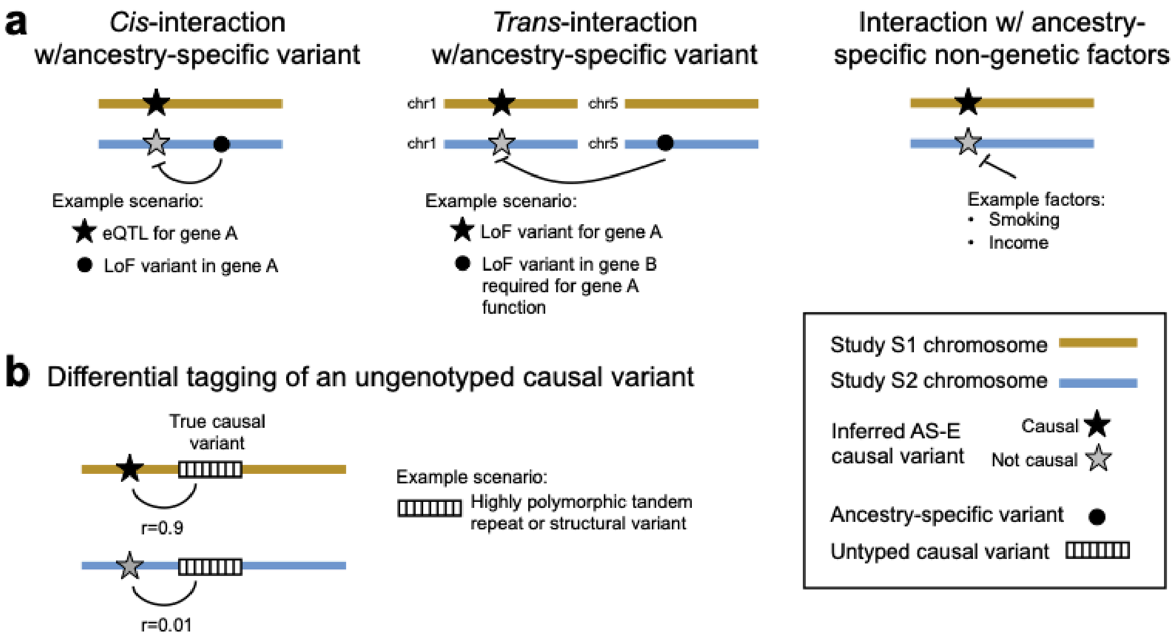


**a. Example scenarios by which true ancestry-specific variant effects could occur.** These include (left) *cis* interactions between a causal variant and a nearby variant on the same haplotype. For example, an expression quantitative trait locus (eQTL) for a gene might have no effect if the gene is already impacted by a loss of function (LoF) variant; (middle) *trans* interactions in which the effect of a causal variant is modified by a distal variant. For example, if functioning copies of two genes is required for a certain function, if one of them is non-functional then an LoF variant in the second gene might have no effect; (right) a causal variant might interact with ancestry-specific non-genetic factors, causing it to have an effect in one study but not another. For all plots, black stars indicate causal variants, gray stars indicate non-causal variants, and black circles indicate ancestry-specific variants. **b. Example scenario by which a variant can appear to have an ancestry-specific effect.** If the true causal variant is not observed, but strongly tagged by another variant that shows different LD with the causal variant in different ancestries, that variant might appear to be causal in one ancestry but not another.

### Supplementary Fig. 31

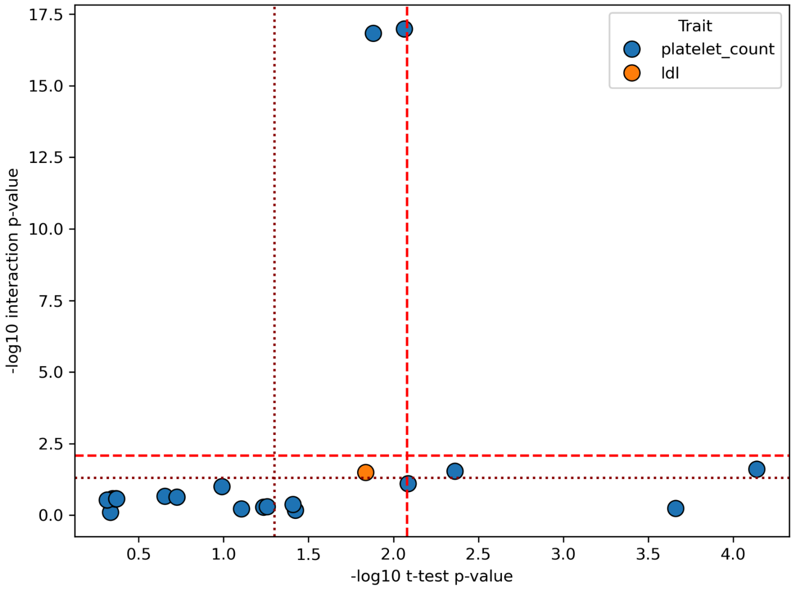


**Significance of the interaction term versus significance of the t-test.** We ran association testing on a candidate AS-E with an interaction term between the candidate AS-E and local ancestry. The -log10 p-value for the interaction term is plotted along the y-axis. We ran association testing on the candidate AS-E in individuals homozygous for European ancestry and non-European ancestry separately and performed a Welch t-test on the effect sizes. The -log10 p-value from the t-test is plotted along the x-axis. The threshold for nominal significance (P<0.05) is shown in the dark red dotted line (lower, left) and with a multiple hypothesis correction in the red dashed line (upper, right).

### Supplementary Fig. 32

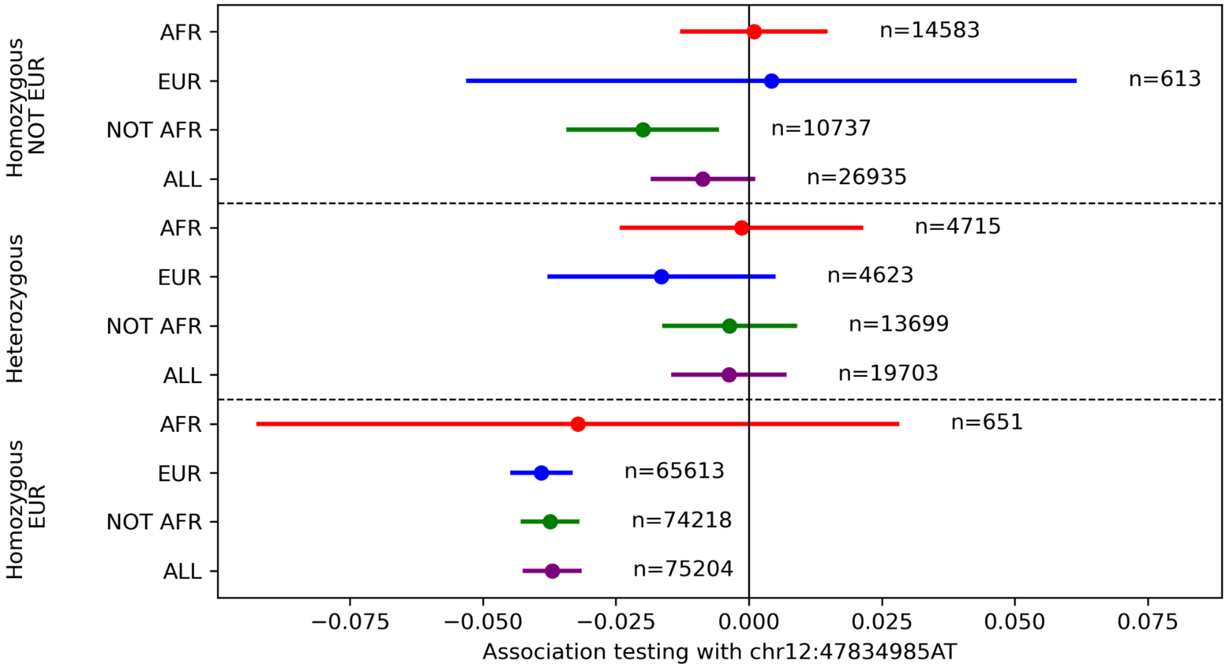


**Stratified regression plot for local ancestry candidate AS-E. (a) Stratified regression for a chromosome 12 trait-region associated with platelet count.** PIPSORT identified 3 candidate AS-Es at this region. Variant rs11608702 (hg38:12:47834985:A:C) is an intronic variant for *LINC02354*. The stratified regression plot for rs11608702 illustrates an example AS-E that shows evidence of being driven by local ancestry rather than global ancestry but for which the test for interaction between local ancestry and variant effect was not significant (P=0.08).

Supplementary Fig. 33

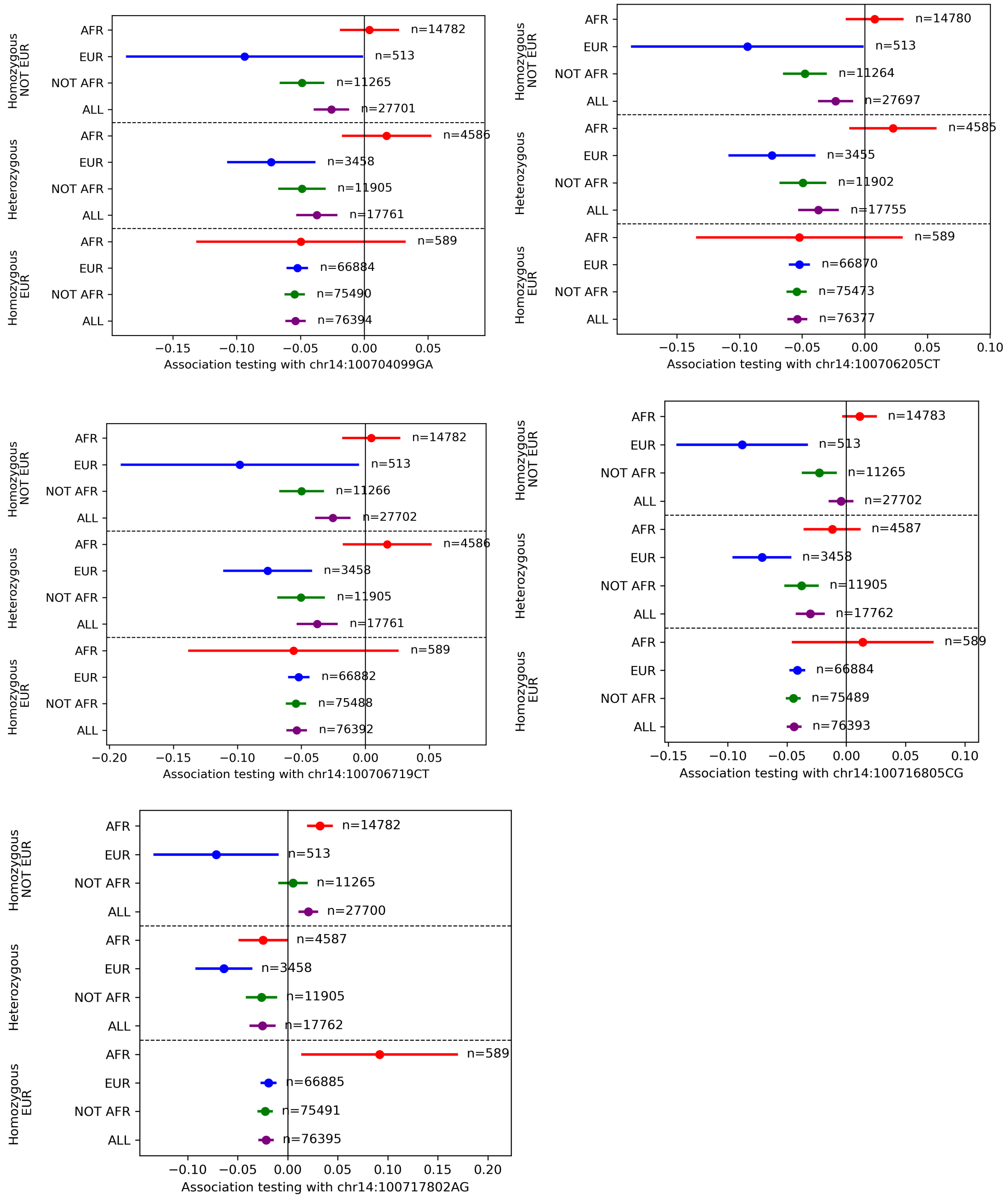


**Additional stratified regression plots for Chromosome 14 AS-E example in Fig. 5.** Stratified regression plots for the other 5 AoU high confidence AS-E variants at the Chromosome 14 trait-region for platelet count in **Fig. 5h-i** are provided here.

### Supplementary Tables

#### Supplementary Table 1

| | MsCaviar | SuSiEx | MESuSiE | MultiSuSiE | PIPSORT |
|---|---|---|---|---|---|
| Allows for heterogeneity in effect sizes | ✓ | ✓ | ✓ | ✓ | ✓ |
| Can directly model a variant present in both studies as causal only in one | × | × | ✓ | × | ✓ |
| Does not assume all causal signals are shared (AS-E, AS-V) | × | × | ✓ | × | ✓ |
| Does not require studies to have the same set of variants (AS-V) | × | ✓* | × | ✓ | ✓ |
| Directly computes ancestry-specific PIPs | × | × | ✓ | × | ✓ |

**Overview of features of existing multi-ancestry fine-mapping tools vs. PIPSORT.** *Although SuSiEx does not explicitly look for AS-Vs and computes only a single (not ancestry-specific) PIP, it does allow for variants that are missing in one study and not another, in which case the study for which it is missing does not contribute to the PIP computation.
